# Barriers to cascade screening in people at risk of Thoracic Aortic Disease: a mixed methods evaluation from the DECIDE-TAD initiative

**DOI:** 10.1101/2023.12.02.23299279

**Authors:** R. G. Abbasciano, J. Dionne, J. Miksza, S. Oczkowski, J. Barwell, N. Shannon, R. Grant, P. Clift, R. Proietti, E. Hope, U. Ahern, K. Hewytt, L. Ghosh, R. Kaur, M. Lewis, A. Cotton, L. Skinner, H. Saadia, G. McManus, N. Qureshi, H. Aujla, S. Page, R. Sayers, M. Bown, J. Maltby, G. Krasopoulos, D. Cameron, A. Oo, J. Elefteriades, G. Owens, G.J. Murphy

**Affiliations:** Department of Cardiovascular Sciences, University of Leicester, Leicester, UK; Department of Health Research Methods, Evidence, and Impact, McMaster University, Hamilton, Canada; Leicestershire Clinical Genetics Service, University Hospitals of Leicester NHS Trust, Leicester, UK; Department of Clinical Genetics, Nottingham University Hospitals, Nottingham, UK; Department of Cardiology, Queen Elizabeth Hospital Birmingham, Birmingham, UK; Liverpool Centre for Cardiovascular Science, University of Liverpool and Liverpool Heart and Chest Hospital, Liverpool, UK; Department of Cardiothoracic Surgery, University Hospital of Southampton, Southampton, UK; Department of Cardiothoracic Surgery, Liverpool Heart and Chest Hospital, Liverpool, UK; Department of Cardiothoracic Surgery at University Hospitals Birmingham NHS Trust, Birmingham, UK; Department of Cardiothoracic Surgery, Barts Heart Centre, St Bartholomew’s Hospital, London, UK; Department of Cardiothoracic Surgery, John Radcliffe Hospital, Oxford, UK; Aortic Dissection Awareness UK & Ireland, UK; NIHR School of Primary Care Research, University of Nottingham, Nottingham, UK; College of Medicine, Biological Sciences, and Psychology, University of Leicester, Leicester, UK; Department of Cardiothoracic Surgery, Yale University School of Medicine, New Haven, Connecticut, US; Department of Cardiothoracic Surgery, The Johns Hopkins Hospital, Baltimore, Maryland, US; Leicester Clinical Trials Unit, University of Leicester, Leicester, UK

## Abstract

**Background:** Cascade genetic and imaging screening for relatives of people with non-syndromic thoracic aortic diseases (NS-TAD) is recommended by guidelines. However, the availability and uptake of cascade screening is low. The aim of this study was to use applied health research methods to identify barriers to screening, and strategies to overcome these.

**Methods:** A cohort study using routinely collected health data evaluated barriers to imaging, genetic testing, and treatment for people with NS-TAD. Delphi consensus exercises and workshops evaluated the screening process and patient experience. Focus groups considered strategies to overcome individual and institutional barriers to uptake. A consensus exercise evaluated the evidence to support cascade screening.

**Results:** A cohort study of 33,793 patients with a TAD diagnosis between 2013 and 2018 demonstrated barriers to treatment and imaging surveillance in females, non-whites, and people from-low socioeconomic backgrounds. A survey of aortic dissection survivors and relatives in England reported that 33/70 (47%) of aortic dissection survivors who responded had undergone genetic testing, including 10/22 (45%) with a positive family history of TAD. In first- and second-degree relatives 66/150 (44%) and 32/155 (21%) of respondents were offered imaging or cascade genetic testing respectively. Only 19/70 (27%) probands and 20/155 (13%) relatives who responded reported that they were involved in shared decisions about their care. Barriers to the uptake of cascade screening included limited awareness of the disease and genetic aetiology, poor health literacy, concerns about cost-effectiveness of screening with low detection rates, requirements for life-long surveillance, and the management of uncertain test results. The consensus exercise demonstrated that the certainty of the evidence to guide cascade screening was Low or Very Low.

**Conclusions:** Barriers to the implementation of cascade screening in people at high-risk for TAD occur at multiple levels suggesting that a complex intervention is required to improve equity of access.

## Introduction

Thoracic aortic disease (TAD) causes over 2000 deaths per year in the UK and almost 10,000 in the US.(1, 2) The prevalence of the disease is increasing.(3) TAD has a long latent phase characterised by asymptomatic aneurysm formation followed by presentation with an acute aortic syndrome, most commonly an acute aortic dissection, which has >70% mortality. (4) Over 20% of TAD is caused by a single genetic mutation. Up to 30% of first- and second-degree relatives of people with TAD will also have recognised mutations or aortic aneurysms. (5)

Cascade screening, where relatives undergo genetic tests and imaging significantly reduces mortality through detection of latent disease, effective secondary prevention, and early treatment(6). It is recommended by treatment guidelines if the proband is aged < 60 years at presentation, or if there is a family history.(4, 7) However, the access to, and uptake of cascade screening is highly variable, particularly in those with Non-Syndromic TAD.(8)

The aim of this study was to define the organisational and individual barriers to cascade screening in people at risk of NS-TAD, and to consider the scope and design of an intervention to overcome these.

## Methods

### Analyses Plan

The project included: **1.** A cohort study to determine if regional factors and common causes of health inequality influenced access to care, imaging surveillance, and genetic testing for TAD. **2.** A survey of aortic dissection survivors and their families, focus groups, and focused interviews to determine the screening process and patient experience, and identify strategies to overcome individual and institutional barriers to uptake. **3.** A Delphi consensus exercise to identify key research questions in cascade screening formulated as Population, Intervention, Comparator, Outcome (PICO). Systematic reviews addressed each PICO, followed by Grading of Recommendations, Assessment, Development, and Evaluation (GRADE) (9) to assess the certainty of the evidence to underpin best practice.

The cohort study was conducted and reported according to the Strengthening the Reporting of Observational Studies in Epidemiology (STROBE) statement.(10) The qualitative research initiative was conducted and reported according to the Standards for Reporting Qualitative Research (SQRQ) checklist.(11) The systematic reviews were conducted and reported according to the PRISMA checklist.(12) The treatment guidelines were reported according to the AGREE recommendations (13) as described in the **Supplementary Material**.

### Ethics

The project was co-led by aortic dissection survivors and relatives in a partnership with Aortic Dissection Awareness in the UK and Ireland. There was no industry involvement in any aspect of this work. The work was conducted according to the Declaration of Helsinki. Ethics approval was obtained from the University of Leicester for the qualitative research initiatives, and the cohort study. The project was funded by the National Institute for Health and Care Research (NIHR203302). The views expressed in this article are those of the authors and not necessarily those of the NIHR.

### Cohort Study

The aim of the cohort study was to establish whether common causes of health inequality; defined as sex, ethnicity, social deprivation, geography or age contributed to unwarranted variations in treatment and survival, as well as access to imaging surveillance and genetic testing.

#### Cohort

The analysis cohort was identified from the HES admitted patient care (APC) data in England from 01/04/2013 to 31/04/2018 with linked Office for National Statistics (ONS) mortality data. The index date for each patient was defined as the first episode within the time period with a TAD diagnosis or procedure code. To identify a cohort with newly diagnosed TAD, subjects with TAD diagnosis in prior 5 years, prior diagnosis of endocarditis, LVAD or heart transplant, inconsistent timeline or residence outside England were excluded. Comorbidities were identified within the two years prior to index date. The data was linked to HES diagnostic imaging (DID) and outpatient datasets to obtain information on diagnostic scans and genetic screening appointments respectively.

#### Exposures and Outcomes

The primary outcome of interest was all-cause mortality at 1-year post diagnosis. Secondary outcomes were: **1.** A recorded intervention on the thoracic aorta within 6 months of diagnosis. We have previously demonstrated that unwarranted variation in this outcome results in unwarranted variation in mortality (14) **2.** An aortic imaging surveillance scan (CT, MRI or echocardiography) within 6 months of discharge, as per treatment guidelines (4, 7), or **3.** Review by a clinical geneticist with 6 months of diagnosis.

Exposures of interest were **1.** Region, defined by postcode. **2.** Common causes of health inequalities, including age, sex, social deprivation quintile and ethnicity.

Predefined HES ICD 10 diagnostic codes and OPCS4 procedure codes for the exposures and outcomes are listed in the **Supplementary Material**.

#### Analyses

Cox proportional hazards models were fitted for time to death within 1-year with adjustment for exposures of interest and comorbidities. A competing risks model was also fitted to account for the competing risk of death, with receiving a TAD procedure within 6 months as the outcome variable adjusted for the same variables. A logistic model was fitted to calculate the predicted number of elective TAD procedures and imaging procedures by postcode area. The expected and predicted number were used to calculate a standardised ratio and plotted on a funnel plot.

### Qualitative Research and Stakeholder Surveys

The qualitative research used a methodological framework which included the Unified Theory of Acceptance and Use of Technology (UTAUT and UTAUT2), the Theory of Planned Behaviour, and Health Literacy models. (15, 16). Health Literacy models (17) explored the ability to access, understand, evaluate, and use health information, crucial for making informed health decisions, particularly in technology use.

#### Survey

A survey was undertaken among members of Aortic Dissection Awareness UK & Ireland to identify variation in practice and public perceptions of cascade screening. The survey was piloted before dissemination. The survey ran from April 2022 to November 2022. It included a total of 60 questions and was built on the digital platform Survey Monkey (SurveyMonkey Inc.; San Mateo, California, USA; www.surveymonkey.com). Results were reported descriptively.

#### Focus Groups

People with TAD or family members participated in two rounds of focus groups conducted from October 2022 to May 2023. The first round explored peoples’ opinions on all aspects of design and use of a DST to support cascade screening and expanded on the answers from the survey. The second round concentrated on barriers to the use of a DST, particularly around the use of technology. Clinicians involved in the management of TAD were invited to participate in focused interviews that explored the same questions as the focus groups. A series of questions considered strategies to overcome barriers. Data processing prior and during the analysis included transcription with full anonymization and de-identification of excerpts, and general information about the group’s compositions were recorded. The full spectrum of questions and detailed analysis are provided in the **Supplementary Material**.

#### Qualitative synthesis

Analyses of the qualitative data aimed to provide an understanding of the barriers and facilitators influencing the adoption of genetic screening technologies and DSTs, guiding the development of more effective and user-centric tools. This analysis employed a thematic analysis approach (18) which involves the identification, analysis, and reporting of patterns or themes within data. The domains are described in more detail in the **Supplementary Material**.

### Evidence Synthesis and GRADE

#### PICO Consensus

The aims of the evidence synthesis were to determine the certainty of the evidence to underpin cascade screening in TAD. A longlist of PICOs was drafted by the project steering group based on the experience gathered through a previous Delphi exercise (10) and multiple patients and public involvement (PPI) sessions, in two rounds of a digital modified Delphi consultation, between May and June 2022. The Aortic Dissection Awareness & Ireland membership was invited digitally to submit proposals for potential PICOs. The steering group curated the selection of the final list based on thematic areas and requests for prioritisation by the patients.

#### Searches

Search strategies for the PICO questions were developed, based upon published systematic reviews, where available (**Supplementary Material**). Searches were updated to August 2022. The searches were conducted on MEDLINE, EMBASE, and PubMed and search results were subsequently uploaded into Rayyan for screening.(19) Two reviewers (RGA, GJM) screened the titles and abstracts for potentially relevant studies, which then underwent full-text review. Four researchers (RGA, SO, JCD, RG) screened full texts. References of included studies and clinical trial databases were hand-searched for relevant studies. Disagreements about relevance and inclusion during this phase were resolved by involving a third member of the WG (GJM or GO).

#### Data Extraction

Details of included studies were extracted into a standardized, pilot-tested spreadsheet. These included the year and language of publication, participant demographic features (confirmed syndromic condition of the proband, type of underlying syndromic condition, type of participants involved whether First-Degree Relatives or Second-Degree Relatives), interventions and comparators (type of test performed, including imaging modality, type of genetic test), along with indications for treatment in the included study (eg. size thresholds, genetic testing, imaging testing, Z scores or other BSA indices).

#### Quantitative Synthesis

Cochrane methodology was used for data synthesis and analysis.(20) For randomized controlled trials (RCTs), the risk of bias was assessed using the criteria outlined in the Cochrane Handbook for Systematic Reviews of Interventions.(20) Each domain’s risk was expressed as high, low or unclear and the risk of bias judgements across different studies were summarised in a ‘Risk of bias’ table’ and taken into account when considering treatment effects. The Newcastle-Ottawa scale (21) was used for observational studies.

#### Grading of Recommendations, Assessment, Development, and Evaluation

Each PICO question was reviewed by a multi-stakeholder group that considered an evidence summary, details of study designs and pooled effect estimates for each outcome, as well a rating of the overall certainty of evidence for each outcome. The GRADE approach was used to assess the certainty of evidence for each outcome as “high”, “moderate”, “low”, or “very low”.

## Results

### Cohort Study

The analysis cohort included 33,793 participants with a new ICD10 or OPCS 4 code related to a TAD diagnosis between 01/04/2013 and 31/03/2018 (**Figure 1)**.

**Figure 1.**
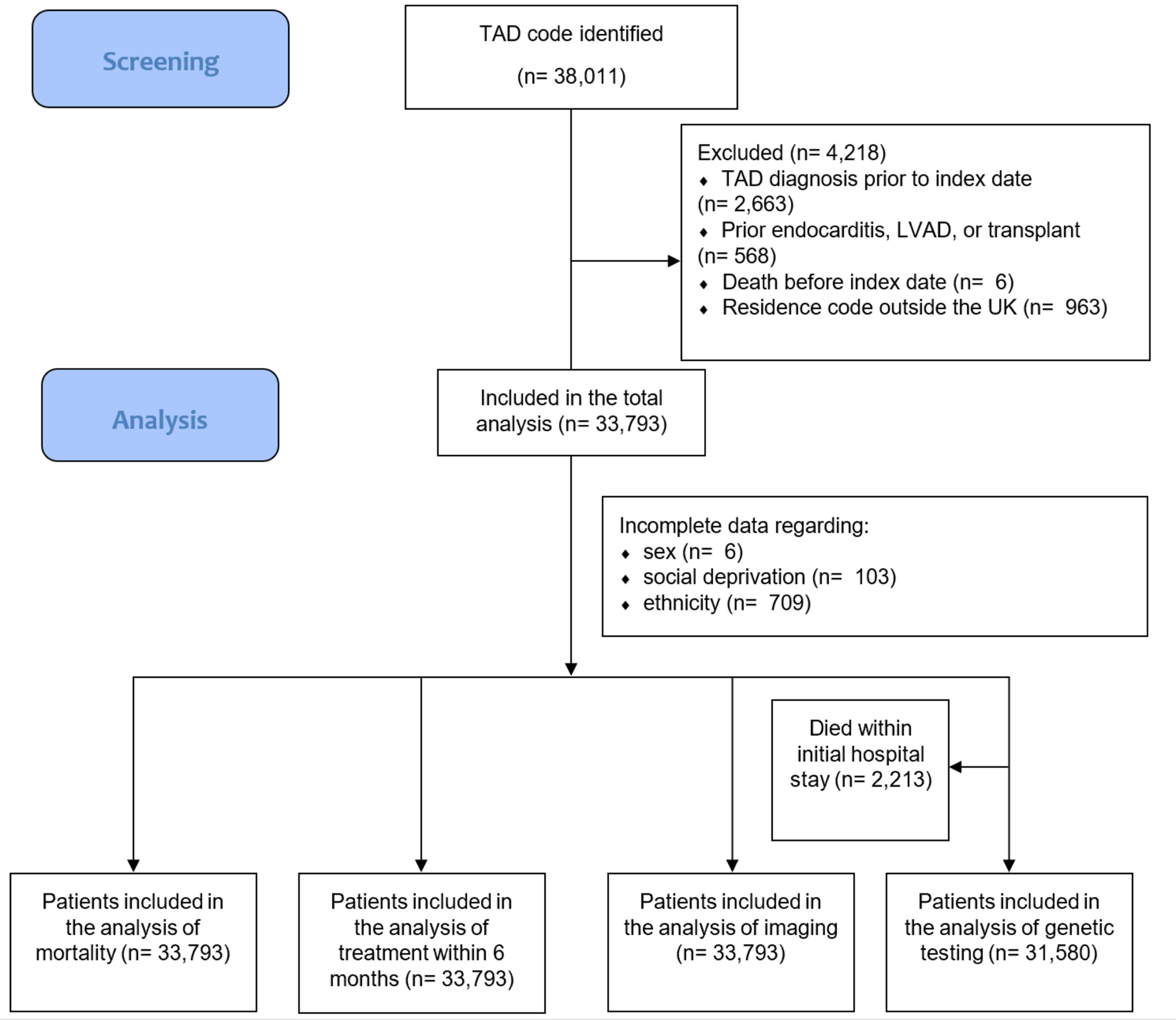
STROBE Flow Diagram for the HES/ONS cohort study The flow diagram reports the number of participants’ data screened and analysed in the cohort longitudinal study. *LVAD – Left Ventricular Assistance Device; TAD – Thoracic Aortic Disease*.

Male sex constituted 20,834 participants (61.7%), the median age was 73 years (IQR 63.0 – 81.0), 30,732 (93.3%) were white, followed by Asian (996, 3.0%), Black (658, 2.0%) and other ethnic groups (555, 1.7%), with 5919 (17.6%) from post codes with the highest levels of deprivation (**Table 1**).

**Table 1.**
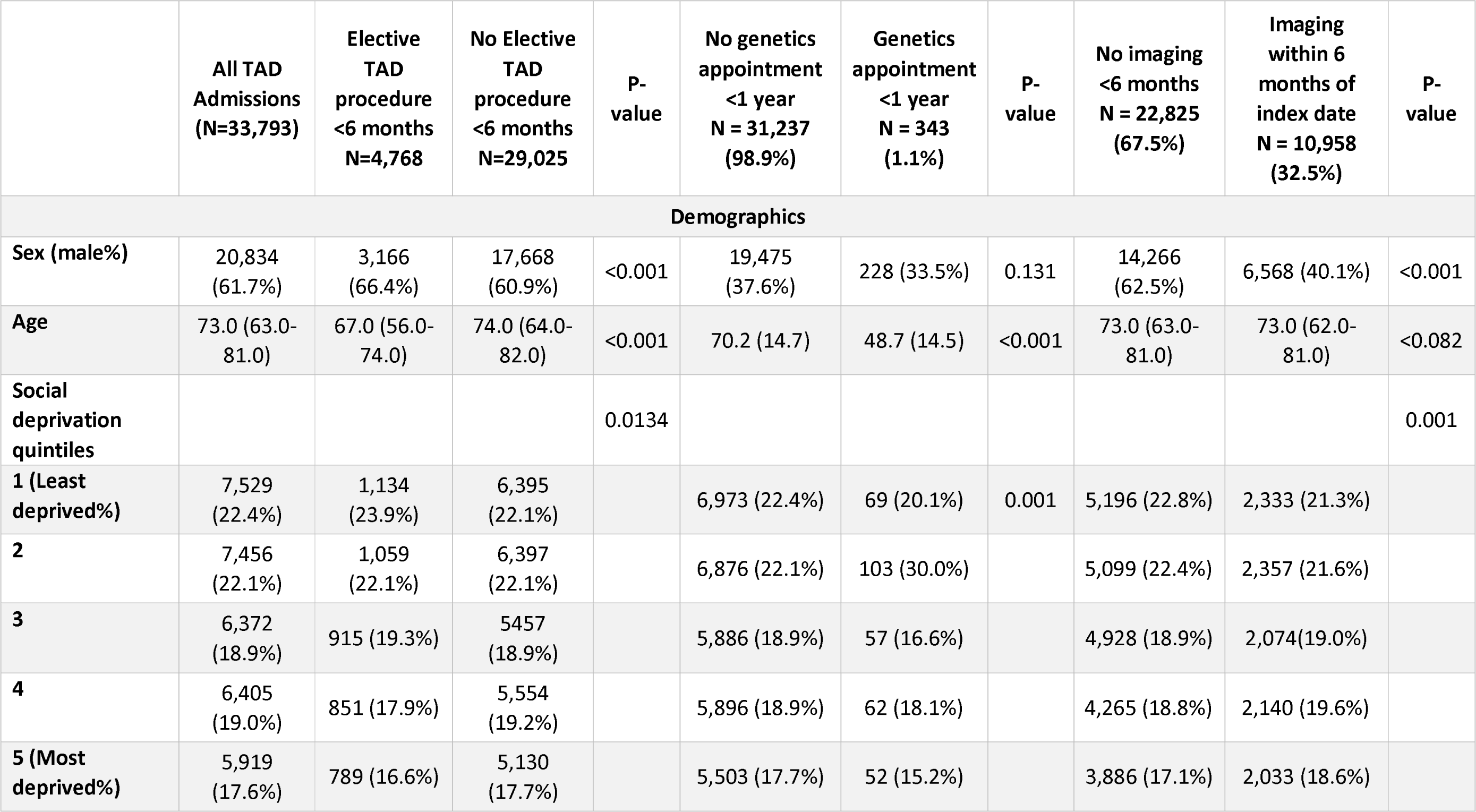
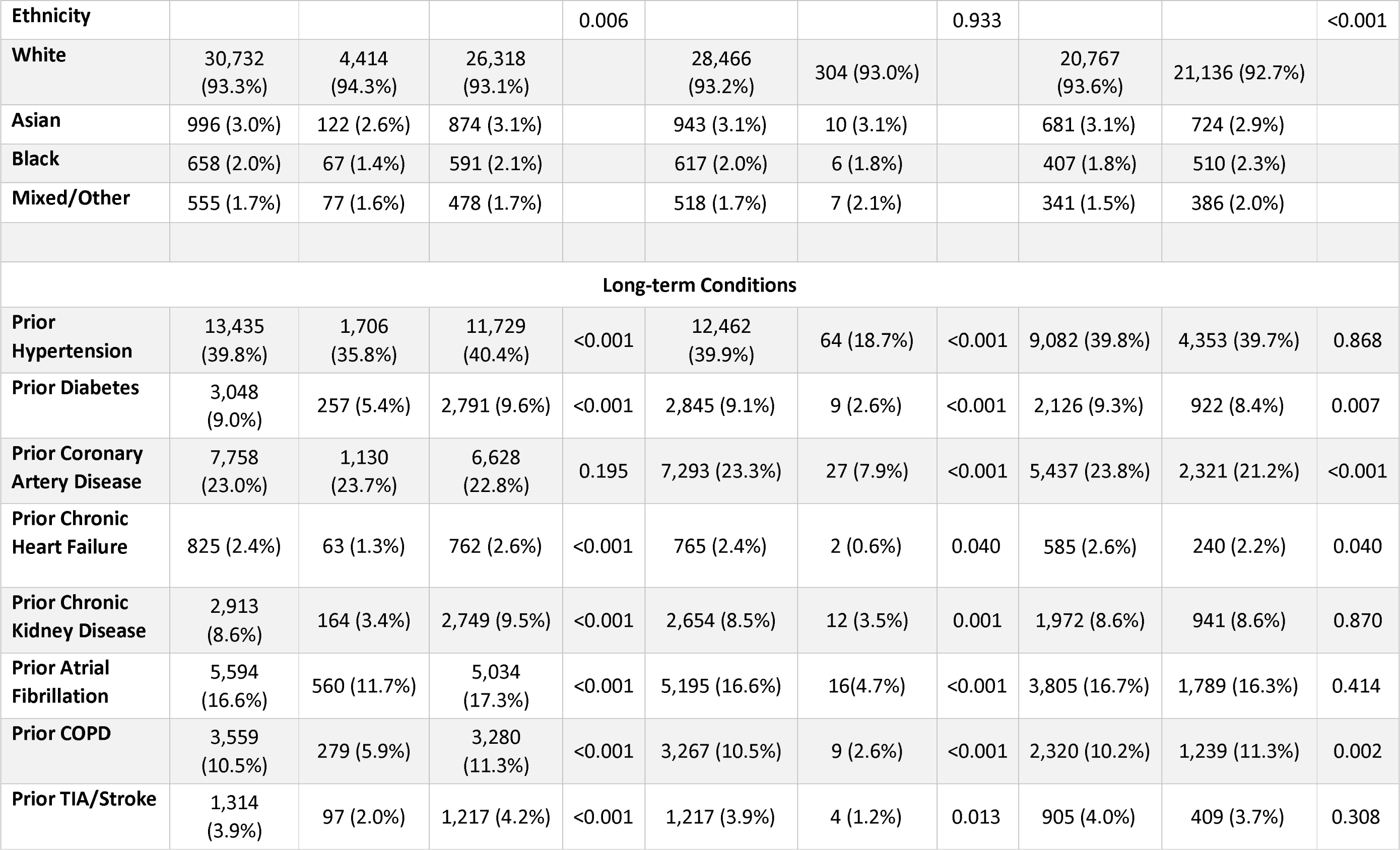
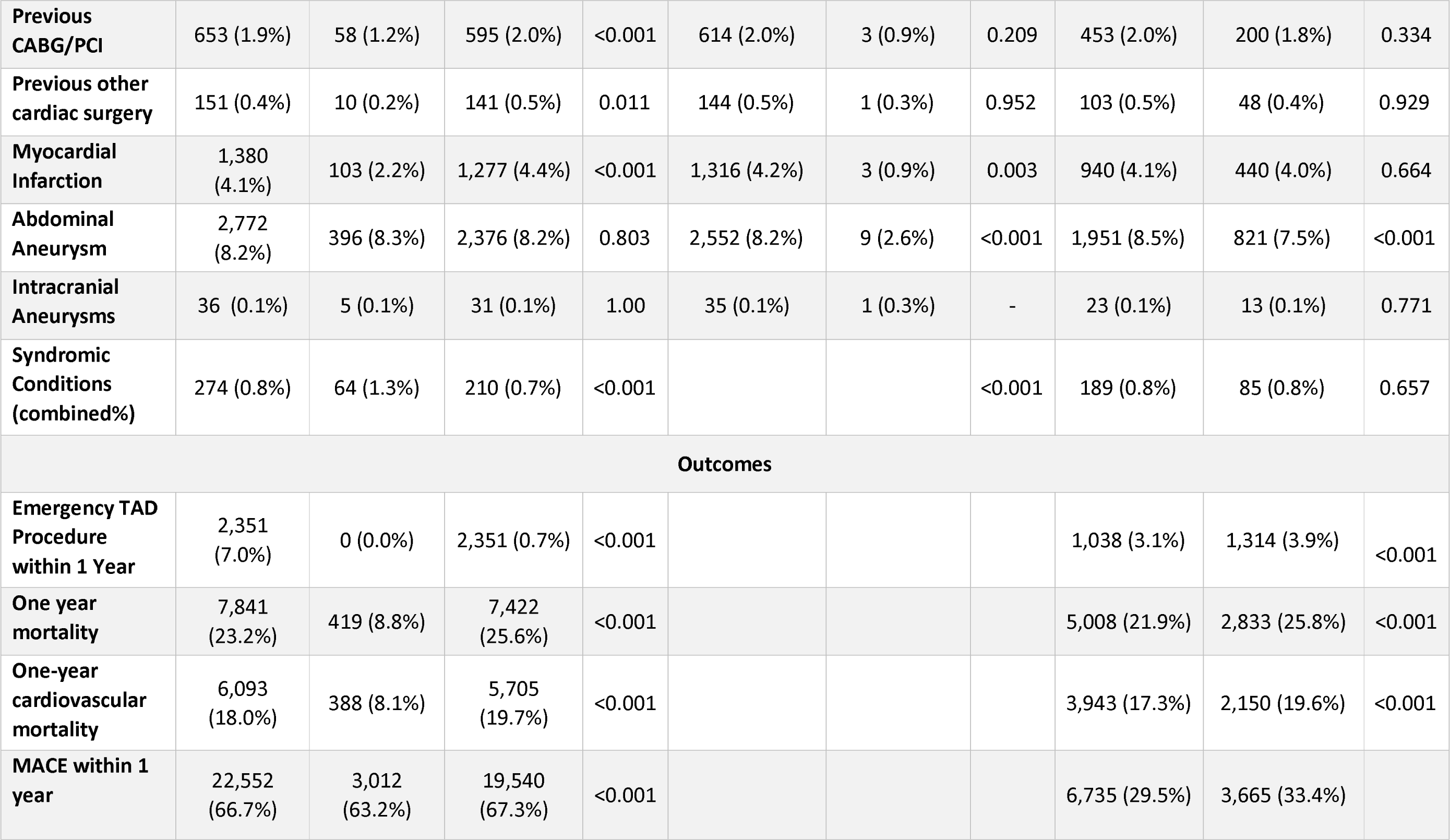
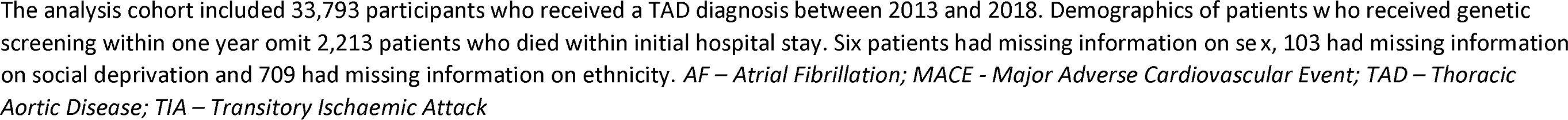
Baseline Status, Exposures and Outcomes of Interest in the Analysis Cohort.

Elective TAD procedures were performed within six months in 4768 patients (14.1%). The five-year survival after a first diagnosis of TAD was 45%. The percentage of elective TAD procedures performed within 6 months of diagnosis varied from 4.1% to 27.8% across postcode areas, with a median of 14.7%. The median proportion of people who underwent surveillance imaging was 34%, ranging from 12% to 60% between post codes. A genetic appointment within one year was received by 343 patients (1.1%).

After adjusting for patient-level characteristics, there were persistent associations between common causes of health inequality, time to treatment, and survival. Specifically, females, people from areas with high levels of deprivation and non-whites and lower treatment rates and higher mortality rates (**Figure 2**). These groups also experienced lower rates of imaging surveillance however these differences were no longer statistically significant after adjustment for baseline differences. Funnel plots for imaging surveillance within 6 months of discharge showed that 24.3% of postcode areas respectively were outside the 99.8% confidence intervals, indicating variation unexplained by patient level characteristics (**Figure 3**). There were too few people who attended a genomic medicine appointment for regression analyses.

**Figure 2.**
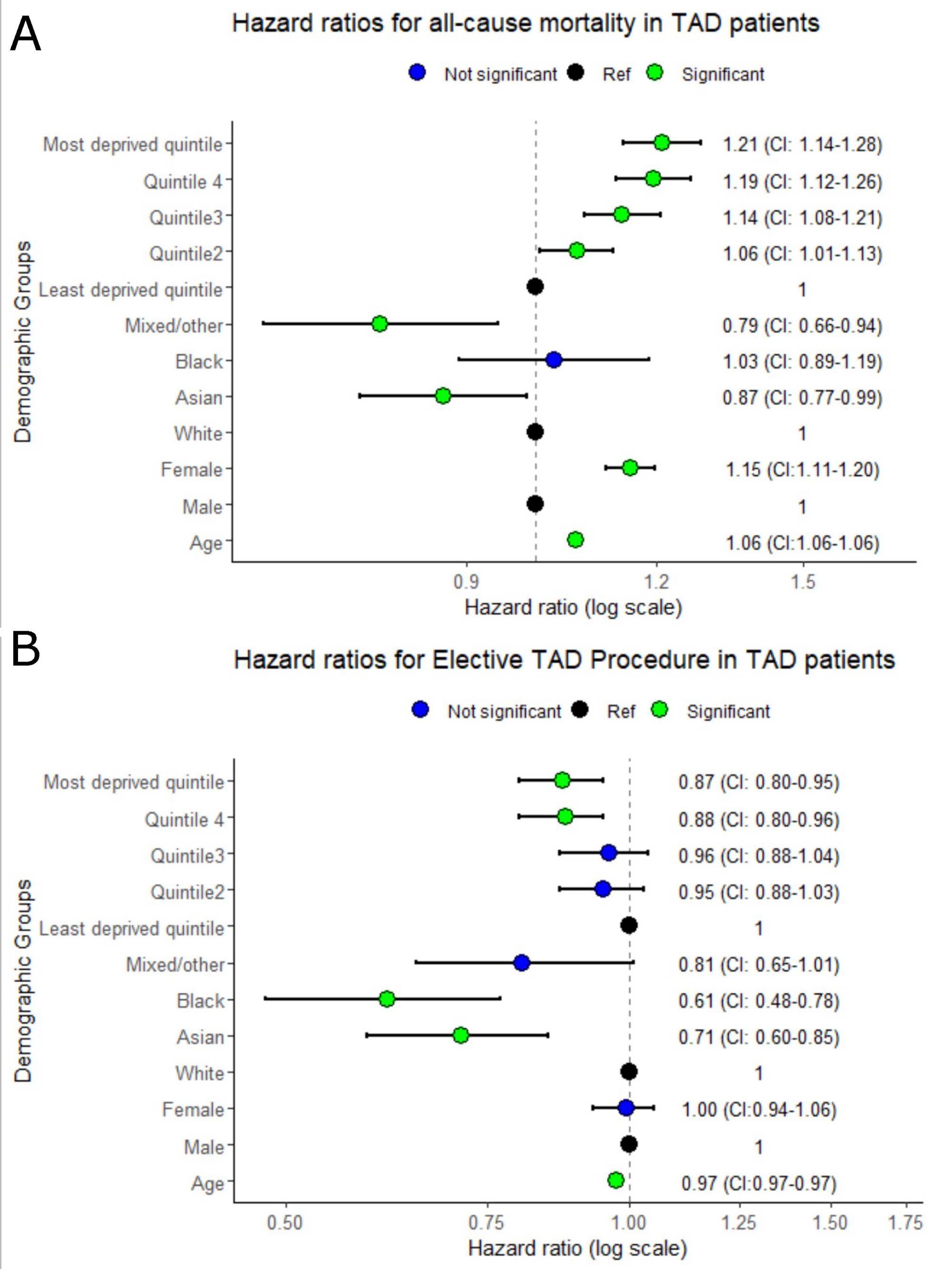
Forest plots with hazard ratios for survival and receiving TAD procedures. The forest plots report the hazard ratios (expressed in logarithmic scale) and confidence intervals for mortality (A) and receiving TAD procedures (B) stratified on social deprivation (in quintiles), ethnicity, sex, and age. Green dots represent significant values, while blue dots stand for non-significant. The black dots are used to mark the exposures considered as reference.

**Figure 3.**
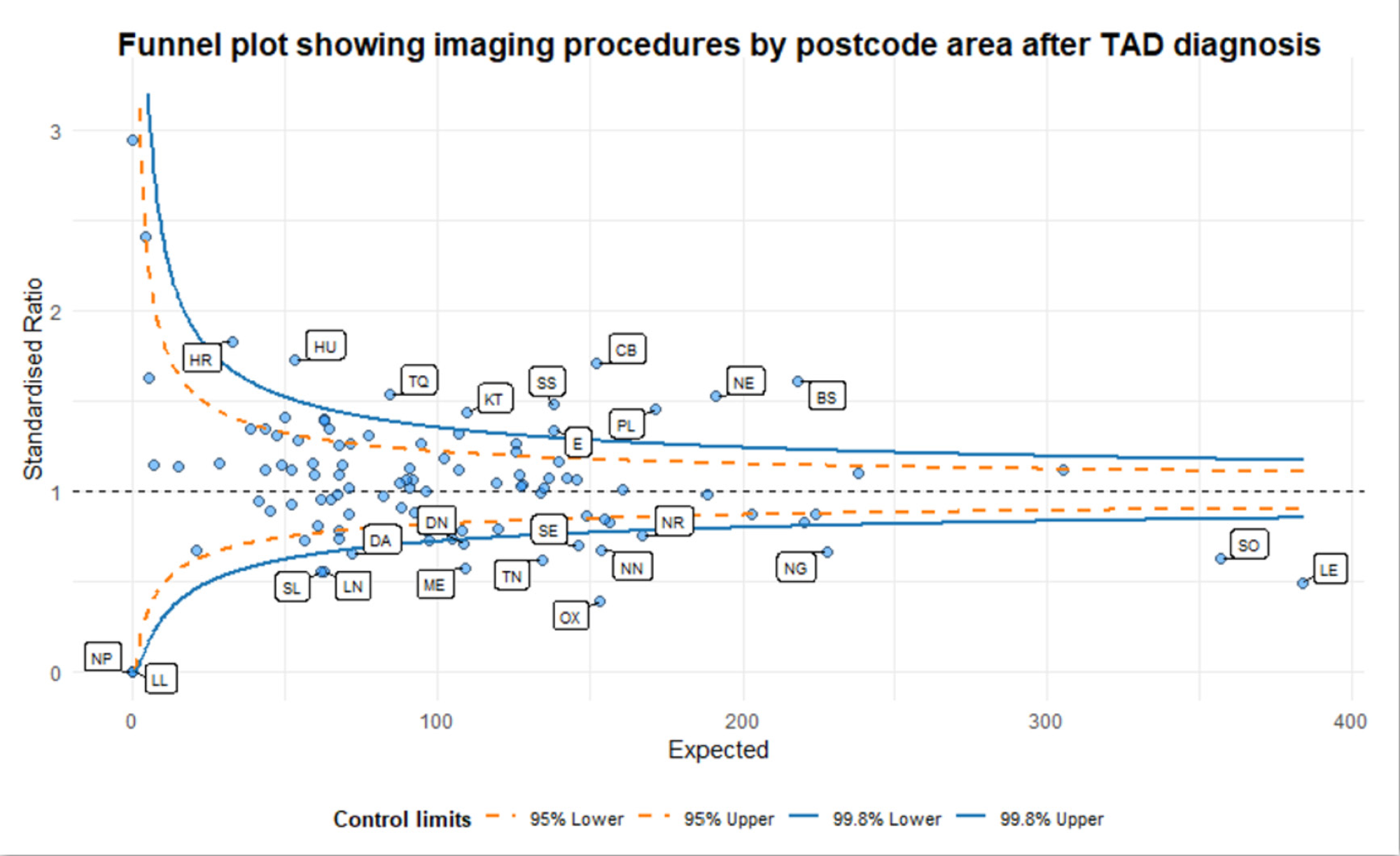
Geographical variation, funnel plot for probability of receiving imaging tests within 6 months of first presentation with TAD Funnel plot showing the standardised ratio of observed/expected case of imaging tests within 6 months from the index diagnosis of TAD for the UK postcode regions analysed, 24.3% of postcode areas were outside the 99.8% confidence intervals, indicating variation unexplained by patient level characteristics. *TAD – Thoracic Aortic Disease*

### Qualitative Research and Stakeholder Surveys

#### Survey

The survey received 71 responses from aortic dissection survivors and 171 responses from relatives (**Figure 4**), for a total of 242 responses. The complete survey results are presented in the **eTable 1**. Only 7 (4.5%) respondents were from non-white backgrounds.

**Figure 4.**
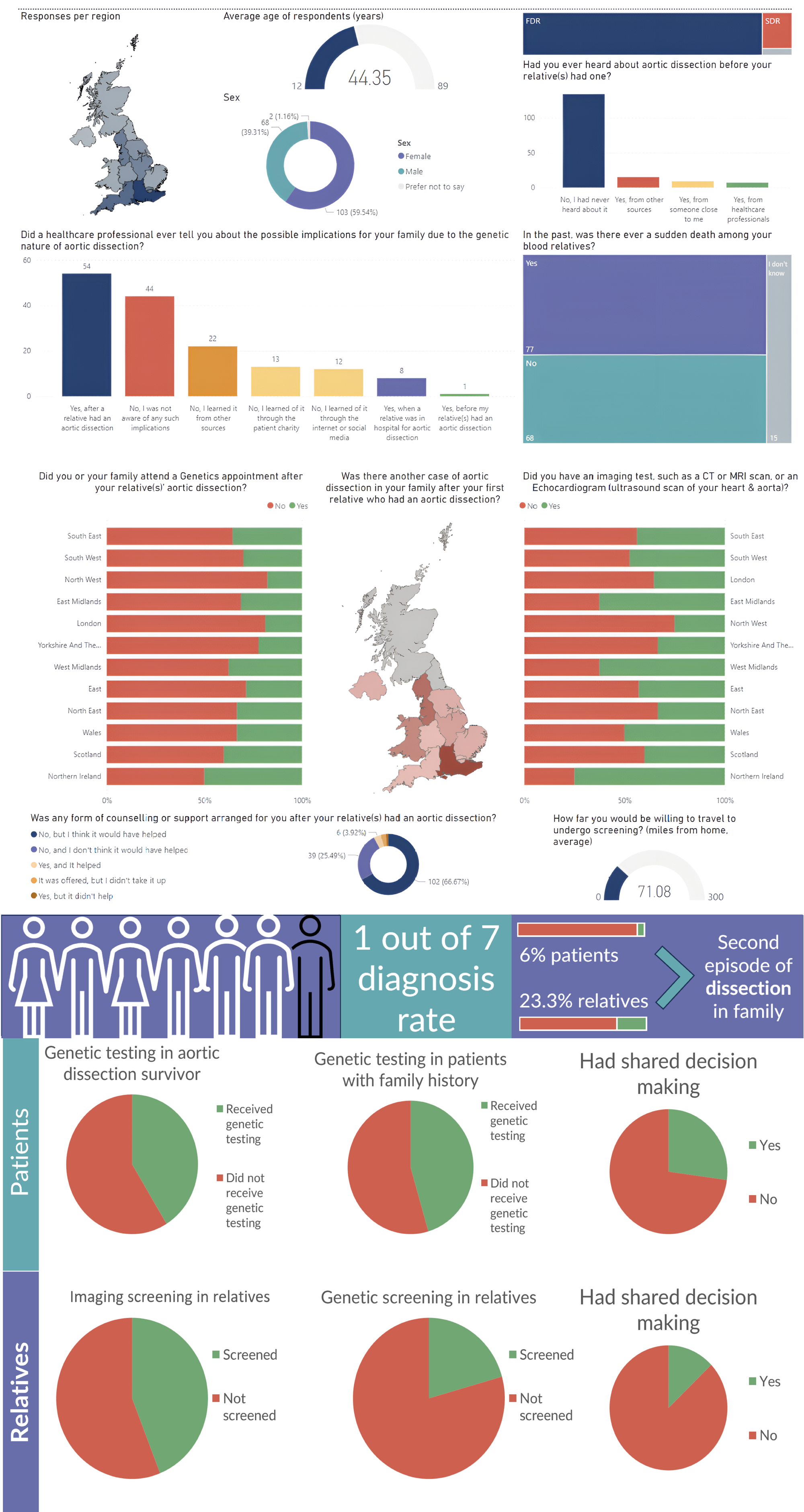
Visual summary of the main findings obtained from the survey of aortic dissection relatives. The survey ran for 7 months, from April 2022 to November 2022 and was completed digitally by 171 respondents. *CT – Computed Tomography; FDR – First Degree Relative; MRI – Magnetic Resonance Imaging; SDR – Second Degree Relative*

The survey demonstrated that 33/70 (47%) of aortic dissection survivors who responded had undergone genetic testing, including 10/22 (45%) with a positive family history of TAD. In first- and second-degree relatives, 66/150 (44%) and 32/155 (21%) of respondents were offered imaging or cascade genetic testing respectively.

Thirty-three out of 69 (48%) of dissection survivors with a positive family history of TAD had received genetic testing. One out of 7 of those tested were diagnosed with a genetic mutation in their family. Sixty-six out of 150 (44%) and 32/155 (21%) of the first- and second-degree relatives of TAD sufferers who responded were offered imaging or cascade genetic testing respectively. Among these, 32/149 (21%) were found to have an undiagnosed aneurysm, and 20/147 (13.6%) had a positive genetic test. Four out of 70 (6%) dissection survivors, and 35/150 (23.3%) relatives who responded had a second family member who went on to have an aortic dissection.

Only 19/71 (27%) probands and 20/155 (13%) relatives in our survey reported that they were involved in shared decisions about their care.

#### Focus Groups and Interviews

Nineteen participants took part in focus groups with 19 participating in the first round and 9 returning for the second round. Of those (n=12) who provided demographic information 5/12 (42 %) were female, 10/12 were of white ethnicity (83%) and 1 (8%) each from Asian and Caribbean backgrounds. The participants had mostly already received genetic screening themselves, although some were at the earlier stages of considering it. Four clinicians took part in the clinician interviews.

Detailed results from the focus groups are presented in the **Supplementary Material** and **eTable 2**. In summary:

***Barriers*** at the level of the individual included limited awareness of the disease and genetic aetiology among clinicians and the public, poor health literacy, and concerns about the cost-effectiveness of screening if detection rates were low in unselected cohorts, the requirements for life-long surveillance, and the management of uncertain test results.

Organisational barriers included clinicians’ concerns about financial constraints, alignment with existing care pathways, potential increased workload. Areas of uncertainty included funding and cost-effectiveness. Attitudes towards adoption varied, and clear implementation plans and providing equitable access to health information were emphasised as necessities.

***Facilitators*** for a Decision Support Tool that would enable shared decision making are shown in **Table 2**. The thematic analysis suggests the development of a DST should incorporate 4 main strategies from a user perspective.

- Improving Usability: Making the DST more user-friendly, with clear and comprehensible information.
- Multilingual and Accessible Content: Ensuring the DST caters to a diverse range of users, including those with varying levels of health literacy.
- Incorporating Feedback Mechanisms: Utilizing user feedback for ongoing improvement.
- Providing Holistic Resources: Offering resources that address broader social and personal concerns related to genetic testing.

**Table 2.** Individual and Organisational Facilitators in the Design of a Decision Support Tool for Cascade Screening in People at Risk of TAD.

| **Domain** | **Facilitators** |
| --- | --- |
| *Usability Improvement* | Address concerns about the clarity and complexity of the DST and genetic testing process. Ensure information is presented in a clear, understandable, and user-friendly manner. Introduce more guidance within the tool to navigate through complex processes, such as genetic testing. |
| *Multilingual and Accessible Content* | Provide multilingual support and other accessibility options to cater to a diverse user base with varying digital literacy and abilities. This could include voice guidance, subtitles, or text-to-speech options. |
| *Incorporate Feedback Mechanisms* | Regularly seek feedback from users to ensure the tool remains user-friendly, effective, and relevant. Feedback mechanisms could include surveys, focus groups, or user testing sessions. |
| *Provide Holistic Resources* | Apart from technical guidance, provide resources addressing social and personal concerns related to genetic testing and TAD, such as potential impacts on employment and insurance. This will help users make more informed decisions. |
| *Inclusion of Health Professionals* | Involve healthcare professionals in the process. Provide them with adequate training on TAD and genetic testing so that they can effectively support and guide patients in using the DST. Healthcare professional endorsement can increase trust and acceptance of the DST among patients. |
| *Implement a Clear and Coordinated Pathway* | Facilitate a clear pathway for genetic testing and TAD treatment. This includes providing comprehensive information about the steps involved in genetic testing and TAD treatment and integrating the DST within existing healthcare infrastructure for seamless patient care. |
| *Data Security Measures* | Address data security concerns by implementing robust data protection measures and being transparent about data collection and usage practices. This will reassure users that their personal information is secure and used responsibly. |

List of facilitators identified at an individual and organisational level identified through the qualitative synthesis of the interviews and focus group activities. TAD – Thoracic Aortic Disease

From an organisational perspective, the DST should be enhanced through organisational strategies such as involving healthcare professionals, creating clear pathways for genetic testing and treatment, and ensuring data security.

### Evidence Synthesis and GRADE

#### Consensus Exercise

The longlist of 14 research questions was reduced to 12 PICOs for prioritisation by the consensus exercise (**Figure 5**). Searches (**eFigure 1**) identified no results for 5 PICOS, so only 7 were subjected to quantitative and qualitative synthesis.

**Figure 5.**
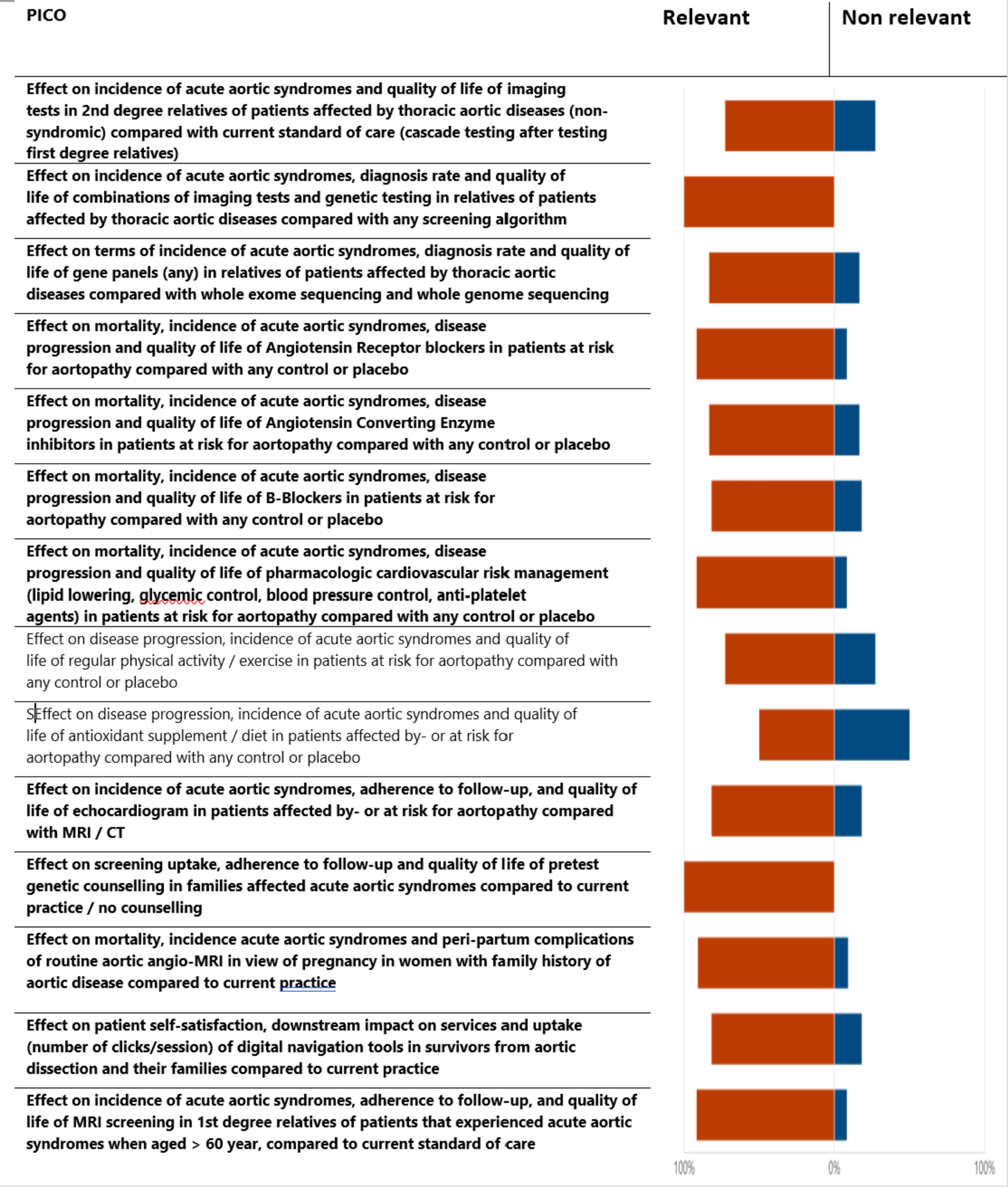
PICOs long list and shortlisting process A longlist of 14 PICOs was formulated based on responses from the first round of a Delphi process, around potential population, interventions, comparisons and outcomes. In an online survey open to patients and clinicians, respondents were asked to select the PICOs that were deemed as relevant research questions. A percentage above 75% as relevant (orange) and below 30% as non-relevant was used as a cut-off to include the PICO in the evidence synthesis exercise. Twelve PICOs (in **bold**) made the final list.

#### Quantitative Syntheses

For PICOs evaluating different screening protocols, all the studies identified in searches were observational. The result of the risk of bias assessments are shown in **eTable 3**. The quantitative synthesis favoured routine imaging tests in first-(test positivity 26%, 95% CI 16% to 4%) and second-degree relatives (test positivity 24%, 95%CI 44% to 11%) (**eTable 4, eFigure 2** and **3**). Genetic testing returned positive results in 21% of cases (95%CI 16% to 26%) when using gene panels and 30% of cases (95%CI 23% to 39%) when using whole exome sequencing (**eFigure 4** and **5**)

For PICOs relating to the secondary prevention of established aortopathy 4 of the 14 included studies were at low risk of bias **(eTables 6 and 7)**. However, applicability was a major limitation, as most trials were conducted in people with syndromic TAD. A summary of the main results is shown in **Table 3** with detailed results shown in **eFigures 8-14**. Quantitative synthesis of RCTs of ARB versus control demonstrate reductions in mortality, but not the frequency of acute aortic syndrome or the need for surgery. RCTs of Beta blockers versus control did not demonstrate superiority for these outcomes, however ARB demonstrated no superiority over Beta Blockers in head-to-head trials. The certainty of these findings was downgraded to Low or Very Low due to imprecision, and applicability concerns. There was no evidence to recommend secondary prevention interventions targeting risk factors for atherosclerotic disease in aortopathy.

**Table 3.**
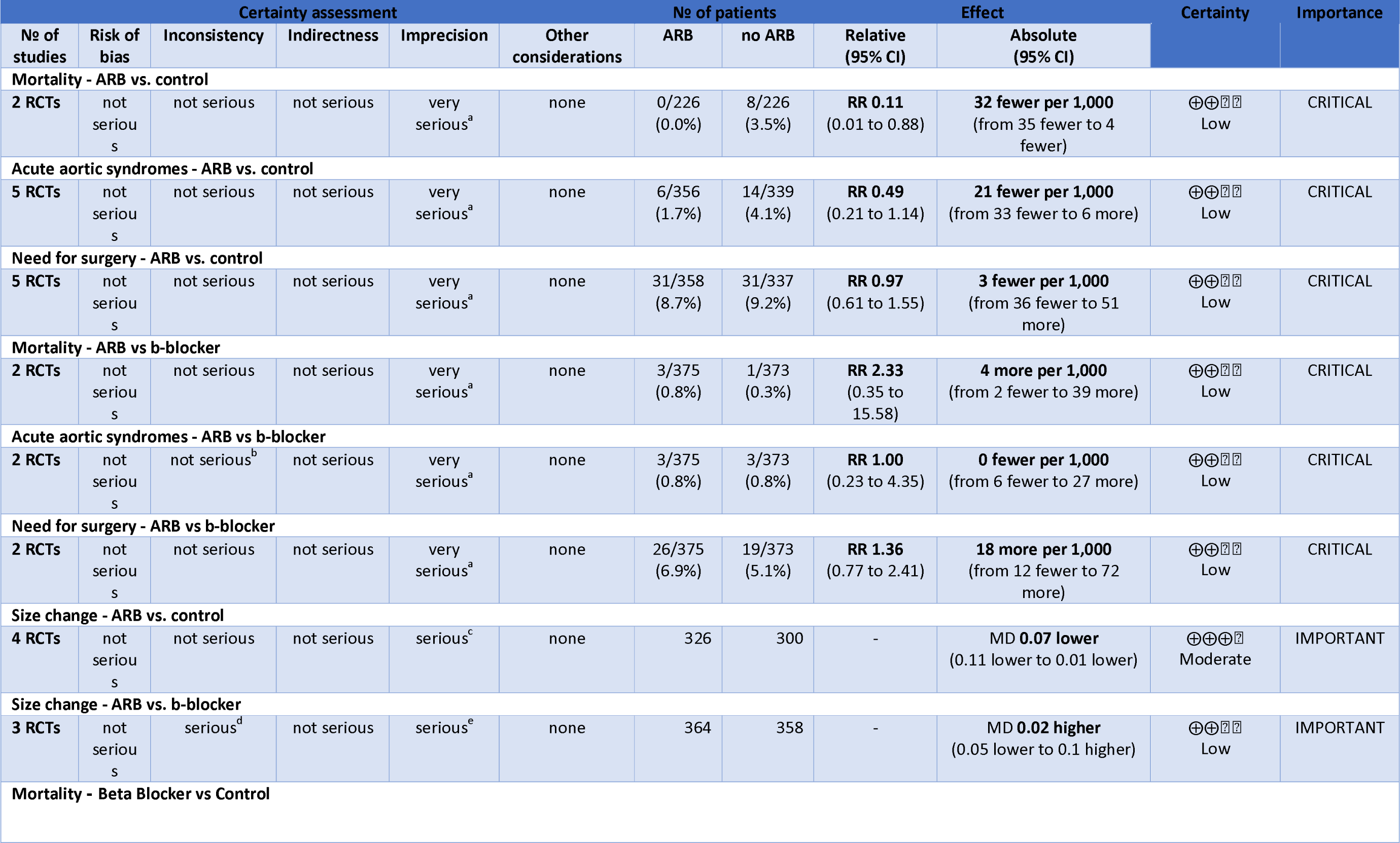
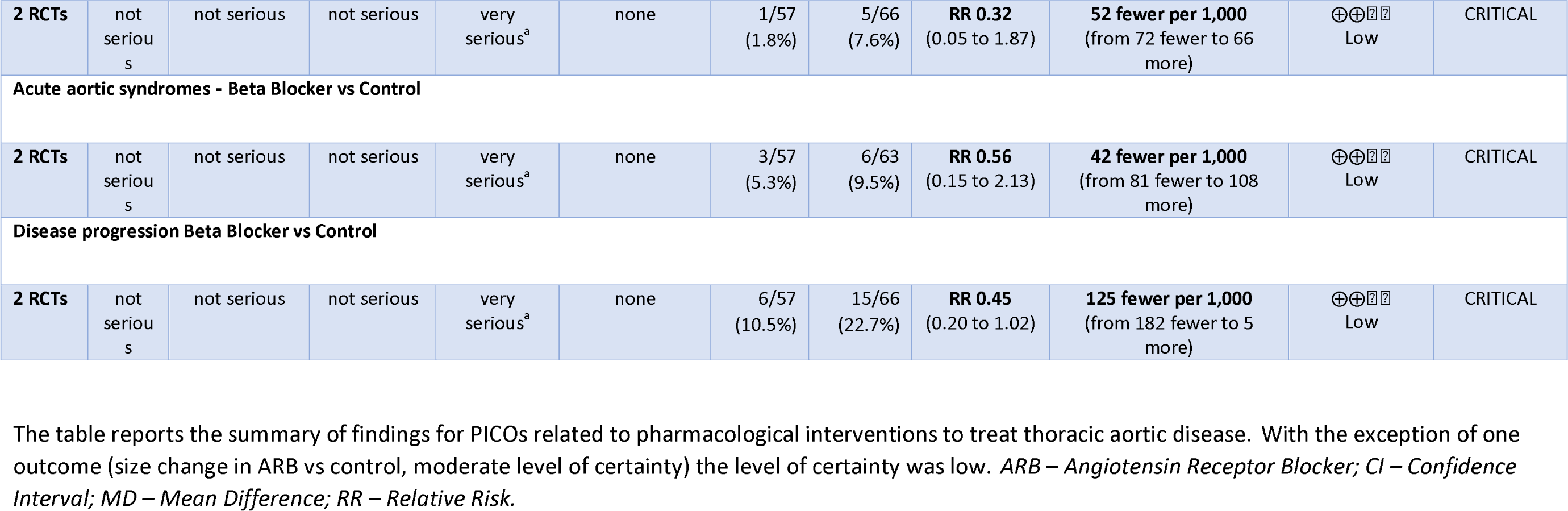
Summary of finding table for the GRADE assessment of the trials evaluating pharmacological interventions for thoracic aortic diseases.

#### Grading of Recommendations, Assessment, Development, and Evaluation

For the GRADE assessments the Evidence-to-Decision frameworks are provided in **eTables 4-13**. A summary of the recommendations is shown in **Table 4**. The expanded rationales are presented in the **Supplementary Material**. Briefly, conditional recommendations with low or very low certainty evidence were made for screening in first-*and* second-degree relatives, the routine use of combined genetic and imaging cascade screening, whole exome sequencing over gene panels, and the application of Decision Support Tools to support shared decision making about screening in people at risk. A strong recommendation was made for routine imaging of all first-degree relatives with NS-TAD. For secondary prevention, conditional recommendations with low or very low certainty evidence were made for ARBs and Beta Blockers in non-syndromic TAD. Research recommendations were made for the comparison of MRI versus transthoracic echocardiography for cascade screening, and the management of non-syndromic TAD in pregnancy.

**Table 4.** Screening, Diagnostic and Therapeutic Strategies in Relatives of Patients with Aortic Diseases.

| Question | Recommendation | Justification |
| --- | --- | --- |
| **<u>Screening</u>** |  |  |
| ***Should imaging tests in first-degree relatives (FDRs) vs. imaging tests in second-degree relatives (SDRs) be used for screening for aortopathy?*** | We suggest the use of imaging screening in FDRs and SDRs (conditional recommendation, very low certainty of evidence). | Patients and relatives prefer to know their risk status and feel reassured by negative tests. The panel recommends screening for first- and second-degree relatives but acknowledges the limitations of the evidence and the uncertainties of clinical outcomes. |
| ***Should imaging and genetic testing vs. genetic testing alone be used for screening of aortopathy?*** | We suggest the use of imaging and genetic screening for aortopathy (conditional recommendation, very low certainty of evidence). | Clinicians in the panel described how the major limitation in the available evidence is the lack of follow up data. Patient members of the panel felt that the presence of an additional secondary test is a valuable form of reassurance, despite the risk of finding variants of unclear significance. |
| ***Should whole exome sequencing vs. gene panels be used for screening relatives of patients affected by thoracic aortic diseases?*** | We suggest using whole exome sequencing over gene panels in screening family members of individuals with non-syndrome thoracic aortic disease (conditional recommendation, low certainty). | Whole exome sequencing reduces the selection bias of gene panels and it can find more cases of non-syndromic TAD, but it can also lead to overdiagnosis and false reassurance. The panel recognises how patients' preferences and values should be taken in consideration. |
| ***Should imaging vs. usual care be used for screening first-degree and/or second-degree relatives of patients with non-syndromic thoracic aortic disease?*** | We recommend extensive adoption of imaging screening in first degree relatives of patients with non-syndromic thoracic aortic disease. (strong recommendation, low certainty of evidence) | The high diagnosis rate in first degree relatives -- with over 1 in 4 relatives having an aortic aneurysm provides an opportunity to avoid a catastrophic event and carries minimal patient harm. |
| **<u>Surveillance and diagnosis modalities</u>** |  |  |
| ***Should decision support tools (DSTs) vs. usual care be used for*** | We suggest using DSTs for survivors of acute aortic syndromes and their families to help guide screening and | DSTs are designed to help patients make informed choices that align with their goals and improve their care |
| ***survivors of acute aortic syndromes and their families?*** | treatment decisions. (conditional recommendation, low certainty evidence) | outcomes. However, most of the existing decision aids assessed were focused on patients with abdominal aortic aneurysms. The quality and accessibility of the tools may also affect their impact. |
| ***Should echocardiogram vs. MRI be used in patients affected by- or at risk for aortopathy?*** | We could not make a recommendation for the use of echo or MRI for screening aortopathy given the lack of evidence comparing the two modalities (no recommendation, very low certainty of evidence). The panel highlighted that patients with type B dissection would be missed on screening with echo compared to MRI screening. | The panel highlighted during review of the evidence that there are potential inequities that exist between these screening modalities. Data is lacking on cost of resources required and cost effectiveness of using one modality compared to another. |
| ***Should pre-test genetic counselling vs. current practice / no counselling be used in families affected by acute aortic syndromes?*** | No studies were identified to inform this question examining these interventions. | The impact of pre-test genetic counselling for patients and families affected by acute aortic syndromes is a research priority. |
| ***Should routine aortic angio-MRI in view of pregnancy vs no routine aortic angio-MRI be used in women with a family history of aortic disease?*** | We could not formulate a recommendation on the specifics of prevention during pregnancy in women with a family history of aortic disease, due to the lack of studies. This topic constitutes a research priority, both for the burden of the disease and for the extent of the evidence gap. | In the absence of research conducted in patients with non-syndromic aortopathy, guidance for the management of pregnancy in aortopathic patients with syndromic conditions (in particular Marfan's syndrome) (26, 27) can be used as an indirect source of recommendations. |
| **<u>Pharmacological management</u>** |  |  |
| ***Should angiotensin receptor blockers (ARB) vs. no angiotensin receptor blockers be used for patients at risk of aortopathy?*** | We suggest using an ARB in patients with Marfan's syndrome, whether or not they are already on a B-blocker. (conditional recommendation, low certainty evidence) | Patients with aortopathy, especially Marfan syndrome, may benefit from ARBs to slow down aneurysm growth and reduce mortality, but the evidence is weak. ARBs may also cause side effects, and while lower doses may help, but their effectiveness is unknown. |
| ***Should beta-blockers vs. no beta-blockers be used for patients at risk of aortopathy?*** | We suggest using B-blockers in patients at risk for aortopathy, specifically those with Marfan's syndrome. (conditional recommendation, low certainty evidence) | B-blockers may slow aortic growth and have few side effects. However, this recommendation is weak, because the evidence is limited, mostly from Marfan syndrome |
|  |  | patients. |
| ***Does treatment of established cardiovascular risk factors improve outcomes in aortopathy*** | 1. We cannot make a recommendation for the use of antiplatelet medication in this patient population.<br>2. We suggest screening for diabetes and having glycemic control in those with aortic diseases to prevent complications as per general cardiovascular risk prevention (Conditional recommendation, very low certainty of evidence).<br>3. We cannot make a recommendation at this time for the use of lipid control in this population. (Very low certainty of evidence). | 1. There is no evidence for the use of antiplatelet therapy in this patient population. Use of antiplatelet therapy should be used based on other cardiovascular risk factors and recommendations in prior guidelines for the use of antiplatelet therapy in patients with known atherosclerotic disease.<br>2. The unexpected finding that diabetes may actually have a protective effect against aortic dissection has attracted considerable attention and controversy among researchers, with inconsistent conclusions, but currently the impact of antidiabetic treatments on TAD development and progression remains uncertain<br>3. Lipid control could be considered in this patient population based on individual factors, but data is lacking on the effect of lipid control in this population on primary aortic dissection prevention. |
| ***Should Angiotensin Converting Enzyme inhibitors (ACEi) vs. no ACEi be used for patients at risk of aortopathy?*** | We made no recommendation on the use of ACEi in thoracic aortopathy due to the lack of evidence. | The WG suggested to consider the use of ARB instead given there is more evidence for this, as described above, at least in patients with Marfan's. |

The table lists the PICO's discussed in the evidence synthesis exercise and the relative recommendations produced by the panel, along with a summary of the justification for each statement. The extended discussion according to the GUIDE evidence to decision framework is available in the Digital Appendix.
ACEi – Angiotensin Converting Enzyme Inhibitors; ARB – Angiotensin Receptor Blocker

## Discussion

### ­Main findings

A mixed methods evaluation including evidence synthesis, qualitative research, and cohort analyses identified multilevel barriers and facilitators to equitable cascade screening for TAD. Barriers included health inequalities associated with sex, ethnicity, deprivation, and geography, low certainty evidence to determine the indications for, or mode of screening, as well as secondary prevention in latent TAD. Organisational-individual barriers included limited awareness and health literacy, as well as concerns about costs effectiveness and the management of uncertain test results.

We speculated that a DST would overcome many of these barriers. Facilitators of an effective DST tool for individuals identified by the research included improving usability, multi-lingual and accessible content, the incorporation of feedback mechanisms, providing holistic approaches that address broader social and personal concerns related to genetic testing. Organisational facilitators included professional engagement, defining clear care pathways and data security. Our searches did not identify any previous or current RCT evaluating the effectiveness of a DST in TAD.

### Clinical importance

The analysis demonstrates that common causes of health inequality; female sex, ethnicity, deprivation, increased age, are prevalent in the care of people with TAD. Strategies to improve the effectiveness of cascade screening must therefore overcome barriers to recruitment and participation from these groups.

The results of the GRADE exercise are in broad agreement with other evidence-based guidelines, where the certainty of the evidence to support cascade screening, as well as effective secondary prevention in NS-TAD was Low or Very Low. For example, the 2022 AHA. ACC guidelines on the management of TAD (22) made a strong recommendation for cascade screening in the families of people with familial TAD, or people who present under 60 years of age. However, these were based on non-randomised data, most often screening studies in people with known genetic mutations, (23) that inflate detection rates. Overall, the evidence synthesis demonstrated an unmet need for RCTs to define the appropriate indications for and mode of cascade screening in the target population.

The qualitative research demonstrated an unmet need for people with TAD and their relatives to be involved in decisions about cascade screening. DSTs increase patient participation in decision making and result in patients choosing options that match with their values.(24) They are most useful where options have benefits and harms that people value differently, as is the case in cascade screening for genetic diseases, and where the certainty of the evidence to support decisions is low. DSTs that anticipate diverse contexts, target populations, and values, can also address health inequalities and regional variations in care. Evidence that DSTs improve clinical outcomes is less certain, (24) however here we suggest that patient empowerment, information, and value-based choices on cascade screening enabled by a Decision Support Tool should increase uptake of screening and early detection of latent TAD, facilitating improved secondary prevention and potentially reducing mortality. The thematic work identified a framework for the development of a DST designed to overcome individual and organisational barriers to implementation. Ultimately the only way to test the validity of this approach is the evaluation of any resulting DST in a randomised trial.

### Strength and Limitations

The work presented was co-produced by clinicians, researchers, and service users including aortic dissection survivors and their relatives. It addresses a top research priority for patients, specifically prevention through improved access to cascade screening. (25) The mixed methods approach places the research in context and, importantly identifies the importance of choice and values-based decision making for clinical progress. Finally, the research is novel in that it has applied behavioural psychology frameworks to the question of cascade screening, providing a new framework for DST development that is likely to improve effectiveness.

Limitations include the inability to directly measure cascade screening in the HES data, so that negative associations between underserved groups and cascade screening can only be inferred rather than demonstrated. In mitigation, the cohort size and longitudinal scope of the analyses provide new insights into regional inequity and the intersectionality of underserved characteristics, treatments, surveillance and outcomes. The strength of representation of underserved groups in the qualitive research was mixed with good representation of females, but less representation from non-white groups, pointing towards an area for improvement for any future research.

## Conclusion

A DST integrated in the existing healthcare infrastructure might mitigate or address the current barriers to access screening in TAD, by helping patients to make informed and value-based decisions about cascade screening, which could also lead to significant health benefits such as higher rates of early diagnosis of preclinical TAD, better secondary prevention and quality of life, and cost-effectiveness in an NHS context.

## Supporting information

Supplemental Methods, Results and Appendices

## Data Availability

All data produced in the present study are available upon reasonable request to the authors

## Notes

### Competing Interest Statement

The authors have declared no competing interest.

### Author Declarations

Ethics approval was obtained from the University of Leicester for the qualitative research initiatives, and the cohort study

