## Supplemental Methods, Results and Appendices for "Barriers to cascade screening in people at risk of Thoracic Aortic Disease: a mixed methods evaluation from the DECIDE-TAD initiative"

### Supplementary Methods

#### Methodological Framework for the Interview and Focus groups

Focus groups were conducted from October 2022 to May 2023 with two rounds of focus groups. The first rounds were to explore the participants opinions around all aspects of design and use of the DST and were designed to expand on the answers given in the patient and relatives surveys. The second round of focus groups concentrated on participants opinions around barriers to the use of the DST particularly around the use of technology. Clinicians were also approached and invited to participate in an interview based on their direct link to the DECIDE-TAD WG or their recognized professional role in the management of TAD. During these interviews the same topics were explored as with the patient and family focus groups. Surveys were created for the clinicians and the patients and family members on their opinions on overcoming the barriers identified and all participants were invited to complete them. Data processing prior and during the analysis included transcription with full anonymization and de-identification of excerpts, and general information about the group’s compositions were recorded.

This work was conducted within a methodological framework which included the Unified Theory of Acceptance and Use of Technology (UTAUT and UTAUT2), the Theory of Planned Behaviour, and Health Literacy models. The UTAUT and UTAUT2 explore frameworks explaining technology adoption through factors like performance and effort expectancy, social influence, facilitating conditions, hedonic motivation, price value, and habit, focusing on users' perceptions and interactions with technology. The Theory of Planned Behaviour explores behaviour as driven by intention, attitude, subjective norms, and perceived control, influencing decision-making processes. Finally, Health Literacy models explored the ability to access, understand, evaluate, and use health information, crucial for making informed health decisions, particularly in technology use.

Therefore, questions around barriers and facilitators centred on participants' and clinicians’ perceptions of technology usage, decision-making influences, and health literacy in relation to a Decision Support Tool (DST) for thoracic aortic dissection (TAD). The full spectrum of questions and detailed analysis are provided in the supplementary material. These questions explored critical aspects such as the importance of health outcomes, comfort with technology, social influences, data security, accessibility, digital literacy, and the role of healthcare professionals in adopting the DST.

**Analysis Strategy**

This analysis aimed to provide an understanding of the barriers and facilitators influencing the adoption of genetic screening technologies and DSTs, guiding the development of more effective and user-centric tools. This analysis employed a thematic analysis approach (23) which involves the identification, analysis, and reporting of patterns or themes within data. Thematic analysis is particularly suited for examining the perspectives of different research participants, highlighting similarities and differences, and generating unanticipated insights. To guide the interpretation of qualitative data we considered the aforementioned theoretical contexts and therefore the analysis specifically looked for:

- Perceptions and Usability of Technology: How participants viewed the ease of use and usefulness of genetic screening technologies and DST.
- Behavioural Intentions: Attitudes towards genetic screening, including perceived benefits and risks.
- Social Influences: The impact of societal norms and opinions of others on participants' decisions.
- Control and Accessibility: Participants' perceptions of their ability to access and effectively use genetic screening technologies.
- Health Literacy Levels: How well participants understood genetic screening information and its implications.

#### Structure and expanded Analysis of the Focus Group Exercise

#### Patients and relatives’ Focus Groups

**Questions**

1. how important is it for you that using the Decision Support Tool (DST) could potentially improve your health outcomes and protect your family from thoracic aortic dissection (TAD) risks?
2. Given your comfort with using technology and the complexity of the genetic testing process, how much would improvements in usability of the DST (such as multilingual support, provision of leaflets, and downloadable information) influence your likelihood to use it
3. How much would concerns about impact on employment and insurance or personal preferences of others in your social circle impact your decision to use the DST?
4. how critical is it for you that there is adequate GP knowledge about TAD and genetic testing, and robust data protection measures, to support your decision to use the DST?
5. how influential is the potential sense of relief or satisfaction derived from protecting your family's health and the usefulness of resources provided by the DST on your decision to use the tool?
6. Considering potential cost savings to the healthcare system from early identification and monitoring of TAD risks and availability of necessary technology within the hospital, how would you rate the importance of these factors when deciding to use the DST
7. how likely are you to incorporate the consistent use of the DST into your regular health management routines
8. how important is it to you that a Decision Support Tool (DST) are viewed positively and seen as a way to protect family members and improve understanding of thoracic aortic dissection (TAD)?
9. Given that the DST provides benefits such as having all information in one place, easy access to resources, and anytime accessibility, how would you rate its importance?
10. How significant is addressing concerns around digital literacy, language barriers, data privacy, and clarity of information for the implementation of the DST?
11. how much importance do you put on the influence of family members, GPs, and societal perceptions when considering the use of a DST?
12. Considering possible resistance from family members towards DST use due to potential impacts on insurance and employment, how important is addressing this resistance to you?
13. Considering the challenges in using the DST, how would you rate the importance of improving control over these processes, for example, by offering multi-language support and downloadable information?
14. How important is the anytime accessibility and multi-format availability of the DST to you?
15. Considering the necessity for secure data handling and transparent data collection purposes, how significant is the role of data security in your decision to use the DST?
16. How would you rate the importance of having healthcare professionals ensure users' understanding around using the DST
17. To what extent would you prioritise overcoming challenges in accessing accurate and relevant information when considering the use of a Decision Support Tool
18. How important is it for you that the DST can cater to individuals with different digital literacy levels, language preferences, and disabilities?
19. Considering your previous experiences of independently researching and evaluating health information, how important is it for the DST to provide clear and easily understandable information to minimise misinterpretation?
20. Given that data privacy concerns and a lack of understanding can compromise decision-making, how crucial is it for the DST to ensure transparent data collection and usage practices?
21. To what extent would you prioritise the role of healthcare providers in improving communication and decision-making support when considering the introduction of the DST?
22. How important is it for the DST to promote health and serve as a preventive measure by helping users understand their personal risk and prevent adverse outcomes?
23. How critical is it for you to address the lack of standardised care and coordination across specialties in the healthcare system through the use of the DST?
24. When considering the introduction of the DST, how important is it to you to have the tool integrated within the existing healthcare infrastructure to ensure its accessibility and usability?

**Questions for priorities**

1.	Acceptance and Training:

- On a scale of 1-10, how important do you think the potential of DST to improve patient care and streamline consultations is?
- How crucial would you rate the training for effective use of the DST in your professional practice on a scale of 1-10?
- On a scale of 1-10, how concerned are you about learning and using the DST?

2.	Organisational Support:

- How would you rate the importance of department heads, senior clinicians, and hospital management in the DST's adoption on a scale of 1-10?
- On a scale of 1-10, how significantly would uniformity in DST usage across the organisation affect your work?

3.	Environment and Barriers:

- How would you rate the potential financial constraints and barriers impacting your use of the DST on a scale of 1-10?
- On a scale of 1-10, how much do you think the compatibility with existing systems would affect your interaction with the DST?
- Given the improved technology acceptance post-COVID, how would you rate its impact on your comfort level with the DST on a scale of 1-10?

4.	Financial Implications:

- On a scale of 1-10, how do you think the funding for the DST would impact your work or department financially?
- How significant would you rate the potential cost savings from DST for the NHS on a scale of 1-10?

5.	Attitudes Towards Adoption:

- On a scale of 1-10, how much do you think your attitude towards technology affects your adoption of DST?
- How concerned are you on a scale of 1-10 about the effort required to learn new systems?
- How much would you rate the potential for mistakes and misinformation as a concern impacting the enthusiasm about DST's benefits on a scale of 1-10?

6.	Implementation Plans:

- How crucial would you rate the need for a clear plan for DST's implementation on a scale of 1-10?
- On a scale of 1-10, how crucial is identifying stakeholders and gaining management support for DST implementation?
- How essential would you rate incorporating DST into treatment pathways on a scale of 1-10?

7.	Access to Health Information:

- On a scale of 1-10, how significant do you think is DST's potential to provide critical health-related data and insights?
- How crucial would you rate the need for effective training to ensure accurate use and understanding of health information provided by DST on a scale of 1-10?
- On a scale of 1-10, how concerned are you about disparities in DST adoption among different groups within the NHS?

#### Key points for narrative (Barriers: Clinicians).

1.	Clinicians emphasise the DST's role in providing essential information on genetic testing and aiding in the decision-making process, enhancing understanding for patients and their families.

2.	Suggestions include implementing a patient progress dashboard within the DST, demonstrating the tool's potential to improve health outcomes.

3.	The DST should be user-friendly and accessible through various formats, like online, app-based, or leaflets, accommodating individuals with different technology familiarity levels.

4.	Clinicians recommend using the DST before engaging with healthcare professionals, reflecting the social context and influence in its utilisation.

5.	The tool needs to be universally accessible and adaptable to varying situations, taking into account differences in healthcare systems that might affect its use.

6.	While not explicit, indications of making the DST enjoyable and satisfying are hinted at by mentions of user-specific content tailoring and ease of navigation.

7.	The DST's value in facilitating genetic testing processes and improving health outcomes indirectly implies its price value.

8.	Clinicians expect DST to be used prior to genetic appointments, suggesting the tool's incorporation into standard procedures and habitual use.

9.	Clinicians see the DST as a tool that needs to be normalised and integrated into the genetic counselling process, indicating professional norms around the tool's usage.

10.	The DST should be made convenient and easy to use by offering different modes of delivery and personalised content, enhancing users' perceived control. The tool's usage is suggested to be planned before discussions with healthcare professionals, showing a strategic implementation plan.

#### Key points for narrative (Facilitators: Clinicians).

1.	Clinicians believe that adopting the Decision Support Tool (DST) could enhance patient care and provide better patient information, making consultations more efficient and job roles easier.

2.	There are concerns that learning how to use the DST could take time and require training, but it is perceived that the tool aligns well with current job roles and is straightforward for clinicians to understand.

3.	Successful DST adoption is suggested to depend on acceptance and support from department heads, senior clinicians, and hospital management.

4.	Usage of the DST is expected to be standardised across the organisation, irrespective of the treating clinician.

5.	Identified barriers to DST adoption include financial constraints, need for additional staff and training, compatibility with existing systems, and potential workload increases.

6.	Facilitating conditions for DST adoption could include post-COVID improvement in technology acceptance, more tech-savvy consultants, and potential integration with clinical records.

7.	There are concerns about the financial cost to the NHS and questions about who would fund the DST. The financial implications might be offset by savings from performing fewer emergency procedures, but this is not confirmed.

8.	Changes in NHS habits due to COVID have led to more openness towards technology use. Some departments already employ apps for patient monitoring, indicating existing habits of tech use.

9.	Clinicians have varying attitudes towards adopting new technology, influenced by leadership, peers, and the NHS culture, indicating the influence of social pressure on technology adoption.

10.	The adoption and effective use of new technologies like the DST in healthcare require clear implementation plans, adequate training, stakeholder identification, superior support, and a dedicated management team. Concerns exist regarding increased workload, procedure complexity, and potential for errors, emphasising the need for careful planning and support.

#### Questions for Clinicians

**Organisational Issues**

Management/Leadership:

1. How do you think the new technology implementation should be managed and led within the NHS?
2. What kind of leadership support do you think is necessary to facilitate successful implementation of the new technology?
3. How can leaders ensure that all staff members feel included and engaged throughout the implementation process?

Organisational Culture: 4. How do you perceive the current organisational culture within the NHS in terms of openness to new technology adoption?

5. What changes, if any, do you think need to be made to the organisational culture to successfully integrate the new technology?
6. How can the NHS foster a culture of continuous improvement and learning to support ongoing technological advancements?

Rewards and Risks: 7. What do you think are the main rewards and risks associated with implementing the new technology within the NHS?

8. How can the organisation ensure that the potential rewards outweigh the risks?
9. What risk mitigation strategies should be in place during the implementation process?

Job Fit: 10. How well do you think the new technology aligns with the current roles and responsibilities within the NHS?

11. What changes, if any, to job roles do you anticipate as a result of implementing the new technology?
12. How can the NHS manage potential job displacement or adjustments related to technology implementation?

Compatibility with the Organisation: 13. How compatible do you think the new technology is with the existing systems and processes within the NHS?

14. What modifications, if any, need to be made to ensure the seamless integration of the new technology?
15. How can the NHS prepare its infrastructure for successful technology integration?

Possibility of Adoption: 16. How likely do you think it is that the new technology will be adopted successfully within the NHS?

17. What barriers to adoption do you foresee and how can they be overcome?
18. What factors are crucial for successful technology adoption in the NHS?

Expected Results: 19. What are the expected benefits of implementing the new technology within the NHS?

20. How can the NHS measure the success of the new technology implementation?
21. What long-term impacts do you expect the new technology to have on patient care and overall organisational efficiency?

**Individual Practitioner Issues:**

Management/Leadership:

1. As an individual practitioner, what kind of support do you need from management during the technology implementation process?
2. How can leaders effectively communicate the benefits and expectations of the new technology to individual practitioners?
3. What role should individual practitioners play in the decision-making process related to new technology adoption?

Organisational Culture: 4. How does the current organisational culture within the NHS affect your willingness to adopt new technologies?

5. What changes to the organisational culture do you think would make you more open to embracing new technologies?
6. How can individual practitioners contribute to fostering a culture of continuous improvement and learning?

Rewards and Risks: 7. As an individual practitioner, what do you perceive as the main rewards and risks associated with the new technology?

8. How can you, as an individual practitioner, ensure that the potential rewards outweigh the risks when adopting new technologies?
9. What steps can you take to mitigate risks during the technology implementation process?

Job Fit: 10. How do you feel the new technology will affect your role and responsibilities as an individual practitioner?

11. What additional training or support do you think you will need to adapt to the changes brought about by the new technology?
12. How can the NHS help individual practitioners manage potential job displacement or adjustments related to technology implementation?

Compatibility with the Organisation: 13. How compatible do you think the new technology is with your current way of working within the NHS?

14. What adjustments, if any, do you think you will need to make to ensure the seamless integration of the new technology into your daily

### Supplementary Results

#### Key points for narrative: Patients, DST scope and priorities.

1. **Belief in Effectiveness**: Participants believe in the effectiveness of genetic testing for thoracic aortic dissection (TAD) as it could help protect family members by revealing genetic susceptibility and enabling preventive measures.
2. **Access and Understanding Challenges**: Participants find the genetic screening process challenging and confusing. General Practitioners (GPs) often lack knowledge about TAD and genetic testing, leading to difficulties in understanding and navigating the process.
3. **Social Influence**: Some family members decline testing due to concerns about the impact on employment and insurance, illustrating social influence in the decision to undertake genetic testing.
4. **Lack of Facilitating Conditions**: Many participants found a lack of facilitating conditions, like GP's knowledge about aortic dissection and genetic screening, which hindered access to genetic testing.
5. **Perceived Price Value**: One participant highlighted potential cost-saving to the NHS by identifying and monitoring people at risk of TAD rather than treating an emergency dissection.
6. **Attitudes towards Genetic Testing**: Attitudes towards genetic testing were generally positive, seeing it as a way to protect family members. Some societal and familial pressures against testing were also noted.
7. **Behavioural Control**: The difficulty to access genetic screening and lack of knowledge among GPs led to perceived low behavioural control.
8. **Intention and Plan to Implement**: Participants demonstrated proactive intentions for genetic screening, even identifying specific healthcare providers to discuss with, but the lack of clear information about the pathway to genetic testing indicated incomplete plans.
9. **Need for Improved Health Information**: Participants struggled to access accurate and useful information about genetic testing and TAD, indicating a need for improved resources and protocols.
10. **Informed Decision-Making and Health Promotion**: Participants were engaged in decision-making, viewing genetic testing as a preventive measure and a tool for health promotion. However, their ability to make informed decisions was compromised due to lack of support and clear information from healthcare providers.

#### Key points for narrative: Patients; Barriers and Facilitators.

**Access to Technology**: Participants generally had access to technology and could access digital platforms. However, concerns about equitable access due to factors like digital literacy, language barriers, or disabilities were raised.

1. **Understanding Health Information**: Participants appreciated the convenience of having all information in one place via the DST. However, concerns about potential misinterpretation of information were raised, suggesting clinicians could help verify understanding after using the DST.
2. **Evaluating Advantages and Disadvantages**: Participants could evaluate the pros and cons of using a DST. Trust was identified as crucial, especially regarding links to external resources and the input of personal data.
3. **Decision-Making Concerns**: While participants were open to using a DST, concerns about data privacy could influence decision-making. The need for transparent data collection and usage practices was emphasised.
4. **DST as a Health Promotion Tool**: Participants viewed the DST as a tool that could promote health by providing information about genetic screening, helping them understand their risks and potentially prevent adverse outcomes.
5. **DST as a Preventive Measure**: The DST was seen as a preventive measure that could potentially prevent aortic dissection if risks are detected early through the information about genetic screening.
6. **Potential Solution to Healthcare System Issues**: Participants saw the DST as a potential solution to issues in the healthcare system like lack of knowledge among GPs, lack of standardisation in patient care, and poor coordination across specialties.
7. **Positive Attitude towards DST**: Participants held positive attitudes towards the use of a DST with technology, appreciating its benefits and convenience. Concerns about digital literacy, language barriers, and data privacy were raised but didn't negate the overall positive attitude.
8. **Perception of Others' Attitudes**: There were varied perceptions about others' attitudes towards the DST. Most participants felt comfortable recommending the DST to family members, but some anticipated resistance from family members, especially those who declined genetic screening.
9. **Perceived Ability and Accessibility of DST**: Participants' perceived ability to use the DST varied, with some expressing difficulty due to health conditions. Preferences for DST to be available in a digital format and at any time were expressed. Concerns about accessibility for older patients and non-English speakers were raised, with suggestions for solutions like multi-language support and downloadable information.

#### Clinicians’ Focus Groups

**High level summary**

1.	Acceptance and Training: Clinicians are optimistic about the DST's potential benefits, like improving patient care and streamlining consultations, but they express concerns about learning and training needed to use it effectively.

2.	Organisational Support: Adoption of DST heavily depends on acceptance and support from department heads, senior clinicians, and hospital management. Uniformity in its usage across the organisation is also crucial.

3.	Environment and Barriers: There are concerns about financial constraints, staff and training needs, compatibility with existing systems, and potential workload increases. However, technology acceptance has improved post-COVID, more tech-savvy consultants are present, and DST's potential integration with clinical records are facilitating factors.

4.	Financial Implications: There are worries about who will fund the DST and its potential cost to the NHS. Despite potential savings from fewer emergency procedures, these are still uncertain.

5.	Attitudes Towards Adoption: Varying attitudes exist, with some clinicians being enthusiastic about the benefits of technology, while others express concerns about the effort required to learn new systems, or potential for mistakes and misinformation.

6.	Implementation Plans: Clear plans for implementation, including identifying stakeholders, gaining management support, providing training, and incorporating DST into treatment pathways, are essential.

7.	Access to Health Information: DST's potential to provide critical health-related data and insights is highlighted, but disparities in adoption among different groups within the NHS are a concern. The need for effective training is emphasised to ensure accurate use and understanding of health information.

#### GRADE Exercise: Rationale for individual consensus recommendations

1. *Should imaging tests in* first-degree relatives *(FDRs) vs. imaging tests in* second-degree relatives (*SDRs) be used for screening for aortopathy?*

**Recommendation**

We suggest the use of imaging screening in FDRs and SDRs (conditional recommendation, very low certainty of evidence).

**Evidence Summary**

The evidence evaluating the exclusive use of imaging testing is limited both by indirectness due to the cohort and screening modalities considered in the available RCTs (Lindholt 2017) (1) and by the small sample size and methodological limitations of the observational studies (Disertori 1991, Marwick 1987, McManus 1987, Warnes 1985) (2-5).

There was no data to support that screening using imaging reduced the incidence of aortic dissection. Nonetheless, according to Lindholt 2017, screening for three conditions (abdominal aortic aneurysm, peripheral arterial disease, and hypertension) resulted in a 0.006 (0.001-0.011) absolute risk reduction compared to the group without screening.

The average diagnosis rate with imaging tests among FDRs was 32% (95% CI 28% - 37%) compared to 21% (95% CI 16% - 27%) among SDRs. The uptake of the screening offer did not differ between the two groups (RR 1.39, 95% CI 0.31 – 6.15).

A recent observational study (Abbasciano 2022) (6) did not find significant differences between FDRs and SDRs undergoing screening, vs. not undergoing screening, either in terms of quality of life nor in terms of anxiety and depression (Table 2).

**Justification**

The available evidence suggests an high diagnostic yield in routinely screening SDRs compared to cascade screening, without the any differences in relatives’ screening uptake or psychological stress. Patient members of the panel described how, once aware of the potential risk of aortopathy, not knowing if one is affected likely has probably a worse impact on quality of life, and routine screening was thus identified as a priority by both patients and relatives. While the clinical value of a negative imaging test might be debatable, particularly in younger cohorts, the patients judged the reassurance offered as a desirable outcome. Given these uncertainties, and the limitations of the available evidence, the panel made a conditional recommendation for imaging screening in FDR and SDR of affected patients.

**Implementation issues**

Different modalities of imaging screening might create specific barriers to access or variations in uptake, but the available evidence did not allow us to address these aspects of the question. Moreover, there are no formal cost and cost-effectiveness studies to inform practice.

2. *Should imaging and genetic testing vs. genetic testing alone be used for screening of aortopathy?*

**Recommendation**

We suggest the use of imaging and genetic screening for aortopathy (conditional recommendation, very low certainty of evidence).

**Evidence Summary**

No randomised controlled trials were identified. Among 40 observational studies (2-41), combining genetic testing with imaging testing did not increase the diagnostic yield (OR 0.93, 95% CI 0.77 – 1.12, very low certainty of evidence for inconsistency across studies, statistical heterogeneity and low number of events). Uptake of the testing was probably higher when genetic and imaging testing were offered jointly (OR 1.22, 95% CI 1.00 – 1.50, very low certainty of evidence for inconsistency across studies, statistical heterogeneity and low number of events).

Thijssen and colleagues (Thijssen 2020) (42) recently performed a health-related quality of life (HRQOL) assessment in 147 patients with TAD, 114 screening participants and 66 partners. The study found that patients with TAD had lower HRQOL, however, patients being screened were not as affected. Young patients and female patients scored lower on the SF-36 and Hospital Anxiety and Depression Scale (HADS) compared to older male patients. Previous aortic surgery was associated with high HADS depression scores.

**Justification**

Clinicians in the panel described how the major limitation in the available evidence is the lack of follow up data which could not capture potential improvement in long term clinical outcomes that could justify the implementation of complex screening programmes. Patient members of the panel felt that the presence of an additional secondary test is a valuable form of reassurance and therefore carries significant desirable effects, despite the risk of finding variants of unclear significance during a genetic test.

**Implementation issues**

Costs deriving from the implementation of a combined approach to screening in TAD might be significant, but no direct evidence is available for a cost-effectiveness analysis.

Beil and colleagues (Beil 2021) (43) described their experience in disclosing pathogenic variants to thoracic aortic dissection biobank participants. The total expense per person was $400, which covered verification and mailing letters, for 26 participants. The total expense per person who got genetic counselling and confirmation from a certified lab was $605.

3. *Should. whole exome sequencing vs. gene panels be used for screening relatives of patients affected by thoracic aortic diseases?*

**Recommendation**

We suggest using whole exome sequencing over gene panels in screening family members of individuals with non-syndrome thoracic aortic disease (conditional recommendation, low certainty).

**Evidence Summary**

We did not find any studies directly comparing the diagnostic accuracy of whole exome sequencing to gene panels. Six observational studies studied testing with a gene panel for TAD in non-syndromic found an average diagnosis rate of 21% (95% CI 16% to 26%) on a total of 390 participants. Whole exome sequencing was used in eight observational studies; on average, the diagnosis rate was 30% (95% CI 23% to 39%) in a cohort of 278 participants with non-syndromic conditions. No studies reported clinical outcomes of either screening approach.

A single study presented data regarding psychological outcomes in 10 patients included in a thoracic aortic dissociation biobank, who were offered underwent whole exome sequencing, offer of clinical genetic counseling and confirmatory genetic testing (Beil 2021) (43). The participants reported low levels of psychological distress after the genetic counselling session (the average FACToR score was 16.0 ± 4.2, out of 100). The subscale scores were also low for negative feelings (3.7 ± 3.4; range 0–12, out of 12), uncertainty (2.0 ± 1.7; range: 0–5, out of 8), and privacy concerns (1.7 ± 2.0, range 0–5, out of 8). Participants had level of positive emotional responses to the genetic test result (8.7 ± 3.8; range 0–12, out of 16). The average decisional regret score was low, 11.5 ± 11.6 (range 0–25, out of 100) indicating low levels of regret about deciding to learn their genetic test results.

**Justification**

Non-syndromic TAD are linked to family history. Screening relatives of probands/patients may detect early disease, and thus allow follow-up and intervention to reduce morbidity and mortality; on the other hand, screening relatives may cause distress and impact mental health. These outcomes may be impacted by genetic testing strategies with different diagnosis rates (gene panel vs. whole exome sequencing). Given the catastrophic harm of acute aortic events, a higher diagnosis rate in families of patients is desirable, and whole exome sequencing results in a sizable absolute risk difference in diagnosis rate of around 10%. At present, the disease is more likely under-diagnosed rather than over-diagnosed. Whole exome sequencing can also facilitate research, or diagnosis at a later time, as new risk variants are uncovered.

Nonetheless, risks of false reassurance from negative tests may be higher with whole exome sequencing as it may be perceived as a more definitive test than a gene panel. The primary downside of whole exome sequencing is that its higher diagnosis rate may result in overdiagnosis and overmedicalisation; however, these may be mitigated through cautious clinical follow-up. While we did not find direct evidence comparing clinical outcomes between the two genetic testing strategies, the WG judged that underdiagnosis poses a more serious and catastrophic risk, thus favouring a conditional recommendation for whole exome sequencing over gene panels.

As expressed by the patient members in the panel, early diagnosis with ongoing follow-up might be preferrable to some, whereas others may prefer not to be aware of genetic risk. However, in patients interested in genetic testing, the value of a higher diagnosis rate obtained through whole exome sequencing (in the absence of a clinically actionable result) would likely correspond to a desirable effect.

Lastly, whole exome sequencing compensates the inequity in recruitment in the current studies of genes (and the resulting gene panels). The use of whole exome sequencing may become more effective at diagnosis in in diverse groups within a multicultural setting, compared to the limited number of genes tested in a single panel.

**Implementation issues**

Gene panels are easier to implement and require fewer resources than whole exome sequencing, although the costs of latter continue to diminish. The WG judged that the resource difference between these two approaches will continue to change in the long term, especially considering the rapid growth in the science and industry of genetic testing.

4. *Should angiotensin receptor blockers (ARB) vs. no angiotensin receptor blockers be used for patients at risk of aortopathy?*

**Recommendation**

We suggest using an ARB in patients with Marfan's syndrome, whether or not they are already on a B-blocker. (conditional recommendation, low certainty evidence)

**Evidence Summary**

A total of 8 RCTs were identified, (44-51), most included in a recent individual patient-data meta-analysis (Pitcher 2022) (52). All trials were conducted in patients affected by Marfan’s syndrome, including one trial which also included patients with Loeys–Dietz syndrome; the majority of trial participants (~75% in both arms were already on a B-blocker). The primary limitation of the evidence was the small number of events for critical outcomes, such as mortality and acute aortic syndromes.

When ARB compared with placebo, ARB may the risk of mortality compared with placebo (RR 0.11, 95% CI 0.01 – 0.88, n=452, 2 RCTs, low certainty). When compared to B-blockers, the effect of ARB on mortality was unclear (RR 2.23, 95% CI 0.35 – 15.58, 2 RCTs, n=748, low certainty). The number of events in both comparisons was small, 8 and 4, respectively, resulting in low certainty evidence due to very serious imprecision.

ARBs had an unclear effect on the incidence of acute aortic syndromes, both when compared with control (RR 0.49, 95% CI 0.21 – 1.14, 5 studies, n=695, low certainty of evidence) and against B-blocker (RR 1.00, 95% CI 0.23 – 4.35, 2 studies, n=748, low certainty of evidence), and on the need for surgery, versus control (RR 0.97, 95% CI 0.61 – 1.55, 5 studies, n-695, low certainty) and versus B-blocker (RR 1.36, 95% CI 0.77 – 2.41, 2 studies, n=748, low certainty). Increase in aneurysmal size (measured as body surface area-adjusted aortic root dimension Z score) was marginally lower with ARB treatment vs control (MD -0.07, 95% CI -0.11 – 0.01, 4 studies, n=626, moderate certainty), and similar when comparing ARB treatment vs B-blocker (MD -0.02, 95% CI -0.05 – 0.1, 3 studies, n=722, moderate certainty).

Adverse events rate were diverse among the different trials, but with similar incidence between intervention and control group (Forteza 2016 (45): discontinuation of medication 4/70 losartan, 6/70 atenolol; reduced dose for side effects losartan 4/70; atenolol 6/70; Groenink 2013 (46): 17/116 discontinued losartan due to 14 dizziness/low BP; 1 renal dysfunction; 1 fatigue; 1 angioedema; Lacro 2014 (47): serious adverse events 50/305 losartan vs. 40/303 atenolol; Milleron 2015 (48): adverse events possibly related to drug losartan 6/151 vs placebo 0/148; any adverse event losartan 51/151 and placebo 48/148; Mullen 2019 (50): no difference in serious adverse events. 21/104 vs 24/88).

**Justification**

Patients with known aortopathy from any cause, including Marfan syndrome, are likely to highly value interventions that can lower their risk of complications or surgery. The evidence suggests that compared to placebo, ARBs result in a minor decrease in aneurysm growth rate and possibly a decrease in mortality, although certainty of evidence is low. As aortic size directly correlates with need for surgery (itself being an indication to operate) reducing rate of growth may delay the need for operation. The primary downside of treatment with ARBs is adverse events, especially in patients without concomitant hypertension. Reducing the dosage might lower the side and adverse effects, but it is unclear if this would also mitigate the potential benefit of ARB therapy. However, the use of ARBs is clearly feasible, as shown by indirect evidence and long-term use of these drugs in general treatment of hypertension. However when used in the setting of Marfan’s disease, especially in patinets without hypertension, acceptability of preventative treatment may vary.

The WG noted that recommendation is only applicable to patients with Marfan's syndrome; this recommendation does not directly apply to patients without Marfan's (eg. patients with other syndromes or non-syndromic patients) as there was no evidence outside of this population, and there are pathophysiologic reasons to assume patients with Marfan’s may respond differently to ARB treatment compared to other patients. Until more evidence is available the choice of which medication (if any) to be used in patients with aortopathy other than Marfan’s may be guided by the anticipated mechanism of effect (eg. genetics/pathology vs. shear stress etc.) and comorbidities (eg. hypertension, kidney disease, or diabetes).

**Implementation issues**

Compared to B-blockers, ARBs are more costly and require regular monitoring of renal function, resulting in slightly higher resource usage. However, this cost is relatively small. Indirect evidence of cost-effectiveness of anti-hypertensives to treat hypertension suggest that ARBs may be more cost effective than B-blockers, and that either is more cost-effective than no treatment (Park 2017) (53). However in patients without pre-existing hypertension, these comparisons may no longer apply.

There may be variability regarding patients’ willingness to take medications for future event prevention, depending on their understanding and appreciation of prognosis, engagement in their own health, experience with illness, and stage of life. For instance, younger patients may value prevention and side effects differently than older patients. Individualized counselling based upon risk and anticipated adverse events may be required to determine if ARB treatment are consisten with a patient’s values and preferences.

5. *Should Angiotensin Converting Enzyme inhibitors (ACEi) vs. no ACEi be used for patients at risk of aortopathy?*

**Recommendation**

We made no recommendation on the use of ACEi in thoracic aortopathy due to the lack of evidence.

**Evidence Summary**

One trial (Bicknell 2016) (54) assessed the effect of an angiotensin converting enzyme inhibitor (Perindopril) on the growth rate of small abdominal aortic aneurysms (AAAs). The study randomized 224 participants (from 2011 to 2013) into three groups: placebo (n = 79), perindopril (n = 73), and amlodipine (n = 72). The number of AAAs that reached 5.5 cm in diameter or required elective surgery was similar between groups: 11 for placebo, 10 for perindopril, and 11 for amlodipine. The growth rates of small AAAs were lower than expected, but perindopril did not have a significant effect compared to placebo or the combination of placebo and amlodipine, despite lowering BP more effectively.

**Justification**

The WG judged that AAAs are physiologically distinct from TAD and thus the evidence was too indirect to make a recommendation. The WG suggested to consider the use of ARB instead given there is more evidence for this, as described above, at least in patients with Marfan’s.

6. *Should beta-blockers vs. no beta-blockers be used for patients at risk of aortopathy?*

**Recommendation**

We suggest using B-blockers in patients at risk for aortopathy, specifically those with Marfan's syndrome. (conditional recommendation, low certainty evidence)

**Evidence Summary**

Three RCTs (Baderkhan 2021; Ong 2010; Shores 1994) (55-57), alongside a meta-analysis by Gersony et al. including 5 observational studies (Gersony 2007) (58) were used to inform this recommendation. All the studies were conducted among participants with a syndromic aortopathy (eg. Marfan Syndrome, Ehlers-Danlos).

Beta-blockers may reduce mortality (RR 0.32, 95% CI 0.05 – 1.87, 2 studies, n = 123, low certainty) and incidence of acute aortic syndromes (RR 0.56, 95% CI 0.15 – 2.13, 2 studies, n=120, low certainty), but both estimates are of low certainty, limited by very serious imprecision due to a low number of events (6 and 9, respectively).

In observational studies, B-blockers had an unclear effect on the composite outcome death or cardiovascular events (RR 1.42, 95% CI 0.91 – 2.22, 5 studies, n=732, very low certainty). Beta-blockers may to reduce disease progression, measured as a dichotomous event (RR 0.45, 95% CI 0.20 – 1.02, 2 studies, n=123, low certainty).

Adverse events were common. Ong et al. (56) reported that 12% of the patients (3 out of 25) felt tired. Shores et al. (57) found that 33% of the patients (10 out of 30) who received propranolol, a non-selective B-blocker, had adverse effects such as heart block, lethargy, depression, insomnia, dream disturbance, bronchospasm, and increased sensitivity to alcohol.

**Justification**

Compared to no treatment, the use of B-blocker in patients with aortopathy may modestly slow the progression of the disease, but the impact upon patient-important outcomes such as mortality or need for surgery, is unclear due to the very small number of events in the included trials; reassuringly in the RCTs the point estimate does favour B-blockers. The adverse effects of beta-blocker were minimal and mostly tolerable, similar to the rates in other groups who use these medications. Moreover, cardioselective B-blockers, which target specific receptors in the heart, are less likely to cause side effects than non-selective B-blockers, which affect receptors throughout the body, and were implicated in the trial by Shores et al.

Considering the above factors, the panel made a conditional recommendation for B-blockers over no such treatment. There are several caveats to this recommendation. First, the main benefit of b-blockers is that they may slow down the rate of aortic root enlargement; while it is anticipated this could improve the long-term outcomes such as preventing aortic dissection, reducing the need for surgery, or prolonging survival, this is far from clear in the existing evidence. Second, most of the available evidence comes from studies involving patients with Marfan syndrome, so the effects of b-blockers on other types of aortopathy, such as those caused by Ehlers-Danlos syndrome or non-syndromic causes, are uncertain. Third, this recommendation should be considered in the context of other treatment options that are available for aortopathy, such as angiotensin receptor blockers (ARBs), which have also shown promising results in reducing aortic root growth and may have additive effects with b-blockers.

**Implementation issues**

The use of beta-blockers for patients with aortopathy has some challenges in its implementation. One challenge is the possible variability in the patients' willingness to take preventive medications, which may depend on several factors, similarly to what previously described with ARBs (understanding of their prognosis, engagement in their own health, experience with illness, stage of life).

Another challenge is the potentially different effects of beta-blockers for patients with different types of aortopathy, such as Marfan syndrome or Ehlers-Danlos syndrome. In Marfan syndrome, the rate of change in the aortic diameter is more relevant for the risk of dissection than in Ehlers-Danlos syndrome, where dissection may occur more sporadically and less related to size change progression. Therefore, we are more certain that beta-blockers are beneficial for patients with Marfan syndrome than for other patients at risk of aortopathy.

A third challenge is the cost-effectiveness of beta-blockers for aortopathy. Although the individual cost per tablet may not be high, the cumulative cost of antihypertensive drugs over a long period of time can be significant. These costs may be offset by reducing the occurrence of adverse events that require surgery, but we are uncertain about the overall balance of costs and savings. There may also be some variability in how people value taking preventive medication for a long time versus avoiding future illness and surgery.

*7. Effect on mortality, incidence of acute aortic syndromes, disease progression and quality of life of pharmacologic cardiovascular risk management (lipid lowering, glycemic control, blood pressure control, anti-platelet agents) in patients at risk for aortopathy compared with any control or placebo*

*7a. Should Antiplatelet therapy vs. placebo be used for Aortic syndromes?*

**Recommendation**

We cannot make a recommendation for the use of antiplatelet medication in this patient population. There is no evidence for the use of antiplatelet therapy in this patient population.

**Evidence Summary**

Although there is an extensive amount of literature in regard to antiplatelet agents for general cardiovascular risk factors control, there is a lack in the use of this pharmacological strategy in patients with aortopathy. The indirect evidence available is conflicting. Lindholt et al. (Lindholt 2018) (59) followed 148 patients with small aneurysms (30-48 mm in diameter) for a median of 6.6 years as part of a screening trial. The participants were referred for surgery if their aneurysms grew larger than 50 mm. The results showed that low-dose aspirin determined a slower aneurysm growth and lower risk of surgery compared with the control group, especially for those with aneurysms between 40 and 49 mm. However, this effect was not observed for those with aneurysms smaller than 40 mm. The authors concluded that low-dose aspirin may prevent aneurysm progression and surgical intervention for medium-sized aneurysms, but they acknowledged the possibility of residual confounding factors. On the other hand, in the double-blind RCT performed by Wanhainen et al. (Wanhainen 2020) (60) 144 patients with AAA (35-49 mm maximum aortic diameter) from eight Swedish centres received either ticagrelor or placebo and underwent MRI at baseline and 12 months to measure AAA volume growth rate (%) as the primary outcome. The ticagrelor group had a 9.1% increase in MRI AAA volume and the placebo group had a 7.5% increase (P = 0.205) in an intention-to-treat analysis, and 8.5% vs. 7.4% in a per-protocol analysis (P = 0.372). The changes in MRI diameter were 2.5 mm vs. 1.8 mm (P = 0.113), US diameter were 2.3 mm vs. 2.2 mm (P = 0.778), and ILT volume were 12.9% vs. 10.4% (P = 0.590).

**Justification**

Use of antiplatelet therapy should be used based on other cardiovascular risk factors and recommendations in prior guidelines for the use of antiplatelet therapy in patients with known atherosclerotic disease.

**Research Priorities**

Randomised controlled trials in this specific patient cohort are needed to elucidate its impact in disease incidence and progression

*7b. Should Glycemic Control vs. placebo be used for Aortic syndromes?*

**Recommendation**

We suggest screening for diabetes and having glycemic control in those with aortic diseases to prevent complications as per general cardiovascular risk prevention (Conditional recommendation, very low certainty of evidence).

**Evidence Summary**

Cardiovascular diseases and atherosclerosis are more likely to occur in people with diabetes, as high blood glycaemic levels can damage the blood vessels and increase the risk of complications. Moreover, hyperglycemia can also worsen the outcomes of microvascular problems, such as kidney disease and eye damage, in diabetic patients. Therefore, one might expect that diabetes would also increase the risk of aortic dissection. However, several recent studies have challenged this assumption and suggested that diabetes may actually have a protective effect against aortic dissection (61-63). One study looked at incidence of aortic disease incidence with glycemic control (Golledge, 2017), with glycemic control being protective. Most other diseases examined the relationship between diabetes and aortic disease. With aortic disease incidence favouring diabetes (RR 0.81, 95%CI 0.68,0.97), incidence of dissection (OR 0.61, 95%CI 0.42, 0.88), and growth of dissection (HR 0.43, 95%CI 0.24, 0.77).

**Justification**

The unexpected finding described above has attracted considerable attention and controversy among researchers, with inconsistent conclusions, but currently the impact of antidiabetic treatments on TAD development and progression remains uncertain. (64)

**Research Priorities**

The possible link between diabetes mellitus and reduced risk of aortic aneurysm growth or rupture is intriguing, but it is not conclusive. It is only an observation that raises a hypothesis that needs to be tested further. More research is needed to confirm or reject the causal relationship between these two conditions.

*7c. Should Lipid Control vs. placebo be used for Aortic syndromes?*

**Recommendation**

We cannot make a recommendation at this time for the use of lipid control in this population. (Very low certainty of evidence).

Lipid control could be considered in this patient population based on individual factors, but data is lacking on the effect of lipid control in this population on primary aortic dissection prevention.

**Evidence Summary**

Extensive literature is available on lipid-lowering drugs for general cardiovascular control, but there is a lack of high-quality evidence specific to TAD. Yonemura et al. (Yonemura 2005; Yonemura 2009) (65, 66) conducted a prospective, randomized, open-label trial to compare the long-term effects of 20-mg and 5-mg doses of atorvastatin on thoracic and abdominal aortic plaques in 36 hypercholesterolemic patients, using magnetic resonance imaging (MRI), to measure the vessel wall area changes at baseline and after 1 and 2 years of treatment. The 20-mg dose significantly reduced low-density lipoprotein (LDL)-cholesterol levels (-47%) and induced regression of thoracic plaques (-15%) after 2 years, while the 5-mg dose did not (-35% and +7%, respectively). However, the 20-mg dose did not cause further regression from 1 to 2 years (-1%), suggesting that maintaining low LDL-cholesterol levels is necessary to prevent plaque progression. A correlation between thoracic plaque regression and LDL-cholesterol reduction (r=0.61) and between thoracic plaque change from 1 to 2 years and on-treatment LDL-cholesterol levels (r=0.64) was also observed. There was no significant change in mortality during follow up (RR 0.20, 95% CI 0.01 – 3.89, 1 study, low certainty of evidence).

**Justification**

Statins may facilitate regression of thoracic plaques and retard abdominal plaques in hypercholesterolemic patients, but optimal LDL-cholesterol levels are required to achieve these benefits. The evidence is however of very low certainty due to the small sample size and the low number of events. From patients’ perspective, although the effect in slowing disease progression is small, this was considered a valuable outcome. Nonetheless, potential side effects were considered debilitating as well. In conclusion, given the level of evidence, and the indirectness of the patient population, these results might not be applicable to the target population.

**Implementation issues**

Statins are commonly prescribed for patients with cardiovascular diseases, but their role in aortic dissection prevention is unclear. Statins are generally well tolerated by patients, but they may have some side effects such as muscle pain, liver damage or diabetes. The cost of statins may vary depending on the type, dose and availability of the drug in different countries and regions.

**Research priorities**

Although statins may have a protective effect on AD by reducing inflammation and stabilizing the aortic wall, the evidence is limited and inconsistent, and more research is needed to determine the optimal dose, duration and timing of statin therapy for AD patients. Biological plausibility studies are required.

*8. Should echocardiogram vs. MRI be used in patients affected by- or at risk for aortopathy?*

**Recommendation**

**Echo vs. MRI**

We could not make a recommendation for the use of echo or MRI for screening aortopathy given the lack of evidence comparing the two modalities (no recommendation, very low certainty of evidence). The panel highlighted that patients with type B dissection would be missed on screening with echo compared to MRI screening.

**Evidence Summary**

One study compared echocardiograms compared to MRI with respect to incidence of aortic diseases, and showed no difference between the two modalities RR0.98 (95%CI 0.42,2.26) (Abbasciano, 2022). Given few events and wide confidence intervals the overall certainty of evidence was low. Only 1 study examined the use of ultrasound versus CT for screening with few events and wide confidence intervals (RR 0.73, 95%CI 0.43, 1.25) resulted in very low certainty of evidence. Due to the sparse evidence available no recommendation could be made.

**Justification**

The panel highlighted during review of the evidence that the are potential inequities that exist between these screening modalities. Type B dissections may be missed with echography alone, where Type A dissections will be identified. Both types of dissection could be identified using MRI screening. Data is lacking on cost of resources required and cost effectiveness of using one modality compared to another.

**Implementation issues**

Both modalities are currently being used in practice for screening of aortopathies. More research is needed, however, if MRI screening is recommended in the future as the primary screening modality this would require increased access through the UK.

*9. Should pre-test genetic counselling vs. current practice / no counselling be used in in families affected by acute aortic syndromes?*

**Recommendation**

No studies were identified to inform this question examining these interventions.

**Evidence Summary**

No studies identified examined these specific comparisons, studies identified addressed different aspects of care of this patient poptulation A systematic review by Mariscalco et al (Mariscalco 2018) examined the incidence of aortic diseases, effectiveness of screening tests of those with nonsyndromic thoracic diseases, and found that relatives could benefit from personalized screening programs. Other studies identified included reviews of the use of multidisciplinary programs (von Kodolitsch, 2016; Bradley &Bowdin, 2015),assessing quality of life in this population (de Heer, 2019) and examining the disclosure of genetic variants in aortic diseases (Beil, 2021). A research priority is the impact of pre-test genetic counselling for patients and families affected by acute aortic syndromes.

*10. Should routine aortic angio-MRI in view of pregnancy vs no routine aortic angio-MRI be used in women with a family history of aortic disease?*

**Recommendation**

We could not formulate a recommendation on the specifics of prevention during pregnancy in women with a family history of aortic disease, due to the lack of studies. This topic constitutes a research priority, both for the burden of the disease and for the extent of the evidence gap.

**Evidence Summary**

The risk of aortic dissection is four times higher during pregnancy than one year after pregnancy, according to a cohort-crossover study involving approximately 5 million women who had either given birth or had an abortive pregnancy outcome between 2005 and 2013.(67) The researchers estimated the incidence rate ratio to be 4.0 (95% confidence interval, 2.0-8.2) for pregnancy versus the control period.

Braverman et al. (68) analysed the cohort of patients from IRAD that experienced an aortic dissection related to pregnancy. Twenty-nine cases were recorded, which accounted for 0.3% of all aortic dissections in the registry and 1% of aortic dissections in women. Most women (69%) had an underlying syndromic aortopathy or a positive family history (15%), but half of them were not aware of their condition until after the aortic dissection. The aortic dissection was type A in 13 women (45%) and type B in 16 women (55%). The onset time was during pregnancy for 15 women (4 in the first trimester and 11 in the third trimester) and after delivery for 12 women (mean of 12.5 ± 14 days post-partum).

In the absence of research conducted in patients with non-syndromic aortopathy, guidance for the management of pregnancy in aortopathic patients with syndromic conditions (in particular Marfan’s syndrome) (69, 70) can be used as an indirect source of recommendations.

Echocardiography is adequate for assessment of the aortic root and proximal ascending aorta but lacks sensitivity for the distal ascending aorta, the aortic arch and the descending thoracic aorta. MRI is a non-invasive technique for cardiac imaging that has several benefits over other modalities such as echocardiography and CT. MRI can accurately and reproducibly measure cardiac volumes and function, with high spatial resolution and contrast between blood and vascular wall or myocardium. CMR can also image structures in any plane, regardless of acoustic windows. Moreover, CMR does not expose the patient to ionizing radiation, making it a safe option for pregnant women.(71)

**Research priorities**

Until further research becomes available, clinicians should carefully examine family history and clinical signs of aortopathy and maintain a low threshold to implement first-line and second-line imaging in pregnancy.

*11. Should decision support tools (DSTs) vs. usual care be used* *for survivors of acute aortic syndromes and their families?*

**Recommendation**

We suggest using DSTs for survivors of acute aortic syndromes and their families to help guide screening and treatment decisions. (conditional recommendation, low certainty evidence)

**Evidence Summary**

We did not find any direct evidence supporting DSTs, but did find indirect evidence which the WG judged to be sufficiently direct to consider, noting that while the content of the DSTs varied, they addressed cardiovascular diseases with similarly high-risk decisions. Knops et. al. (Knops 2014) (72) conducted an RCT of a decision aid in patients with an asymptomatic AAA, finding them to be better informed and have less decisional conflict regarding their treatment options compared to not using at DST. Eid et. al. (Eid 2022) (73) looked at the factors associated with preference of choice of abdominal aortic aneurysm repair (Open vs. Endovascular). Leblanc et. al. (Leblanc 2018) (74) study looked at whether patient satisfaction with the consent discussion was improved or not by (solely) showing patients their computed tomography or angiography images before they undergo vascular surgery. Overall, DSTs may not have a significant effect on patients’ satisfaction (SMD -0.06, 95% CI -0.25 – 0.13, 3 studies, low certainty evidence), decisional conflict (MD -2, 95% CI -7.38 – 3.38, 2 studies, n=440, low certainty), patient anxiety (SMD -0.07, 95% CI -0.33 – 0.2, 2 studies, n=217, low certainty), or quality of life (MD -1, 95% CI -4.06 – 2.06, 1 study, n=164, low certainty). DSTs may improve patients’ understanding, albeit marginally (SMD -0.27, 95% CI -0.54 – 0.01, 2 studies, n=215, low certainty).

**Justification**

Patients with TAD face difficult decisions about their diagnosis, monitoring, and treatment options. These decisions depend on their personal values and preferences, as well as the potential benefits and harms of different interventions. DST are designed to help patients make informed choices that align with their goals and improve their care outcomes. However, most of the existing decision aids assessed were focused on patients with abdominal aortic aneurysms. Patients who survive an acute aortic syndrome or their families may have different needs and concerns, even if they have similar clinical conditions. Moreover, there is limited evidence on how effective decision support tools are for patients with TAD. The quality and accessibility of the tools may also affect their impact. Therefore, although these would be extremely valuable, more research is needed to develop and evaluate decision support tools that are tailored to patients with thoracic aortic diseases and their specific decisions.

DSTs may also increase equity by providing more standardized information and decision-making processes for patients, including those who may not have access to such information. This depends on whether or not they the tools are provided equitably in a consistent way.

**Implementation issues**

Assuming decision tools are helpful, most patients would agree to their use, as highlighted by the patients’ representation in the panel. However, there are also moderate costs involved in designing, implementing and updating these tools, which may require more resources than simply providing usual care. For instance, in one study (Leblanc 2018), patients receiving the intervention had a longer discussion time than those who did not (8.18 ± 4.28 minutes vs 6.35 ± 2.72 minutes; P = .07). This could nonetheless be seen as a positive outcome from the patient's perspective, as they had more time to review and discuss their options.

The cost-effectiveness of the decision support tools may vary depending on their downstream effects and whether they increase or decrease the overall costs of TAD management. The opportunity costs and time required may also be affected by the type and quality of the tool.

Lastly, many patients with thoracic aortic diseases would likely accept the use of decision support tools, if they supplemented and reinforced current care, and not if they replaced current processes (e.g., meeting with the clinical team).

*12. Should imaging vs. usual care be used for screening first degree and/or second degree relatives of patients with non-syndromic thoracic aortic disease?*

**Recommendation**

We recommend extensive adoption of imaging screening in first degree relatives of patients with non-syndromic thoracic aortic disease. (strong recommendation, low certainty of evidence)

**Evidence Summary**

We identified 5 observational studies relevant to this PICO. (2-4, 6, 41). Imaging of first-degree relatives of patients affected by non-syndromic TAD may result in a high diagnosis rate of 26% (95% CI 16% to 40; 5 studies, n=645, low certainty), and a similar rate in screening of SDRs (24%, 95%CI 0.11 to 0.44; 2 studies, n=26, low certainty). Patients’ depression symptoms (assessed with: PHQ-9) and anxiety symptoms (assessed with GAD-7) were unchanged before and after screening (3 months follow up) according to a single observational study. (6) The overall level of certainty for the outcomes assessed was low, due to the methodological limitations of the studies included (observational studies) and limited sample size, especially for second degree relatives. We did not identify any studies evaluating the clinical outcomes of screening.

**Justification**

Non-syndromic thoracic aortic diseases have a high mortality when they present as emergencies; early detection can facilitate earlier intervention to reduce morbidity and mortality. Evidence suggests that relatives of patients who present with non-syndromic thoracic aortic disease are at increased risk, raising the question of whether screening with imaging is warranted.(75) The high diagnosis rate in first degree relatives -- with over 1 in 4 relatives having an aortic aneurysm-- provides an opportunity to avoid a catastrophic event, and carries minimal patient harm. By comparison, incidental rates of diagnosis of aortic aneurysm (defined as thoracic aorta diameter ≥ 4.0 cm) on CT scans performed at a single hospital on 5662 patients (Mori 2019) (76) was 2.1%, with 97.8% occurring in patients age over 50 years old (rates increased with age, with 5.7% of males over 84 years old having aortic root dilation of >=4.5 cm), demonstrating a disproportionate risk in FDRs. In GRADE, strong recommendations in the setting of low certainty evidence are only made under certain conditions, including life-threatening situations; in this instance, the panel places a very high value on the uncertain but potentially life-preserving benefit of detecting a seemingly common disorder, recognizing that the harms of screening are less clear.

**Implementation issues**

Although the advantage of an earlier diagnosis seems likely, the ratio between benefits and costs of screening relatives of patients with aortic aneurysms are unclear, as there is no direct evidence that screening leads to earlier treatment and better outcomes for the relatives. Screening also requires more resources than not screening, both in the initial tests and follow-up visits for the relatives. The cost-effectiveness of screening is uncertain and depends on whether higher diagnosis rates translate into better outcomes in a cost-effective manner. Screening outside of a research setting may be difficult or expensive, and there may also be issues with access to follow-up care, psychological support, and other services related to screening, with a lack of consistency in the service currently offered to the public across different regions.(77, 78)

### eFigure 1 – PRISMA 2020 flow diagram for the systematic review

**Identification of studies via databases and registers**

Records removed *before screening*:

Duplicate records removed (n = 280)

7115 records identified from databases and registers

**Identification**

Records excluded

(n = 3390)

Records screened

(n = 6835)

Reports excluded:

Ineligible population (n = 13)

Ineligible intervention (n = 7)

Reports sought for retrieval

(n = 41)

**Screening**

Reports included from Mariscalco 2018:

n = 53

74 studies included in the systematic review

**Included**

67 studies included in the meta-analysis

The flow diagram describes the identification, screening and inclusion process for the studies selected for the consensus exercise.

### eFigure 2 – Forest Plot describing the effect of different screening modalities on diagnosis in non-syndromic thoracic aortic diseases

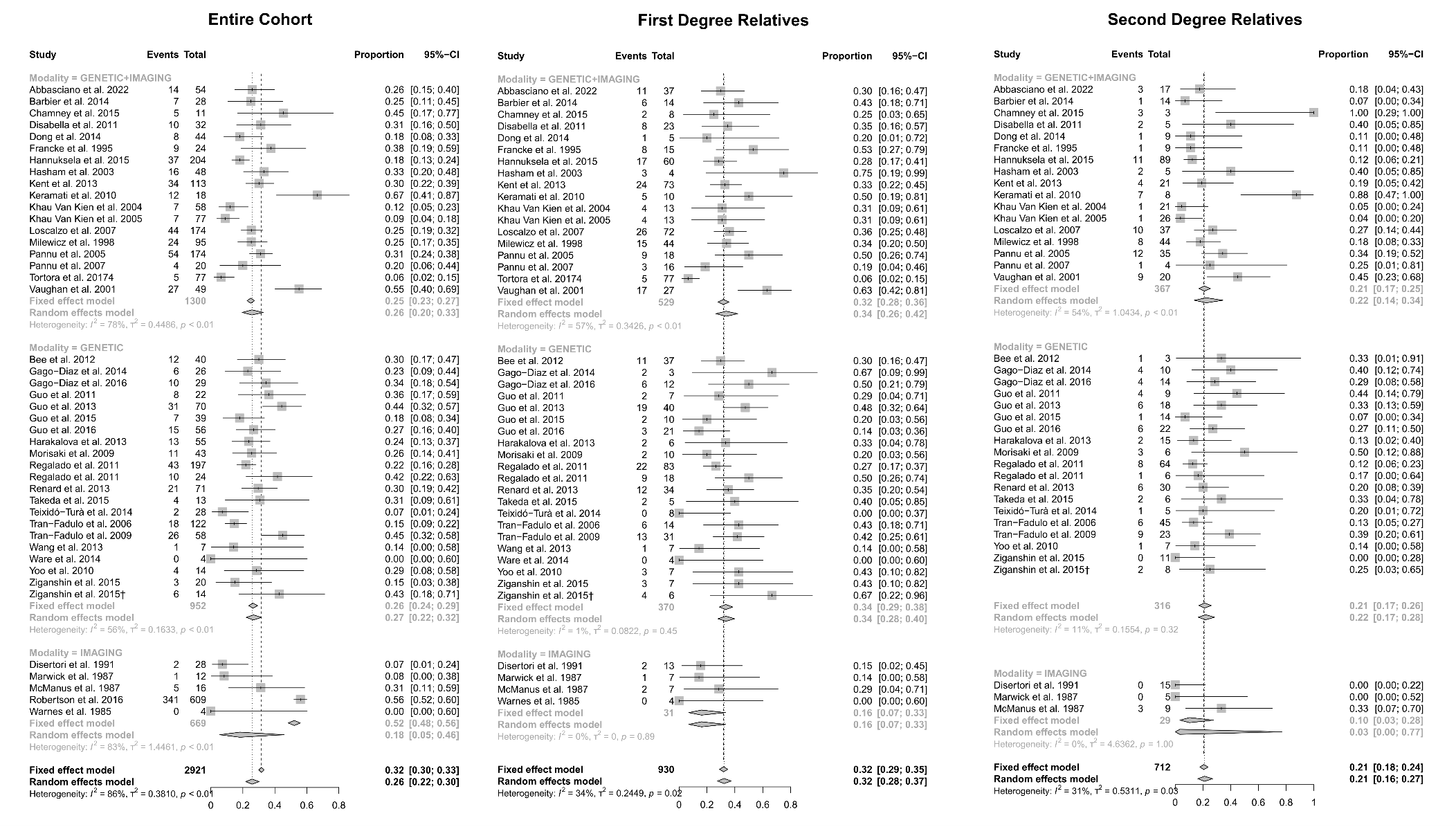

The forest plots reports the results from the proportional meta-analysis of observational studies conducted in families in which one or more relatives were diagnosed with a non-syndromic thoracic aortic disease. The different studies (grouped based on the testing modality) are compared in terms of number of diagnoses, and the results are pooled both in a fixed effect and random effect model. *CI – Confidence Interval*

### eFigure 3 - Forest Plot describing the role of different screening modalities on uptake in non-syndromic thoracic aortic diseases

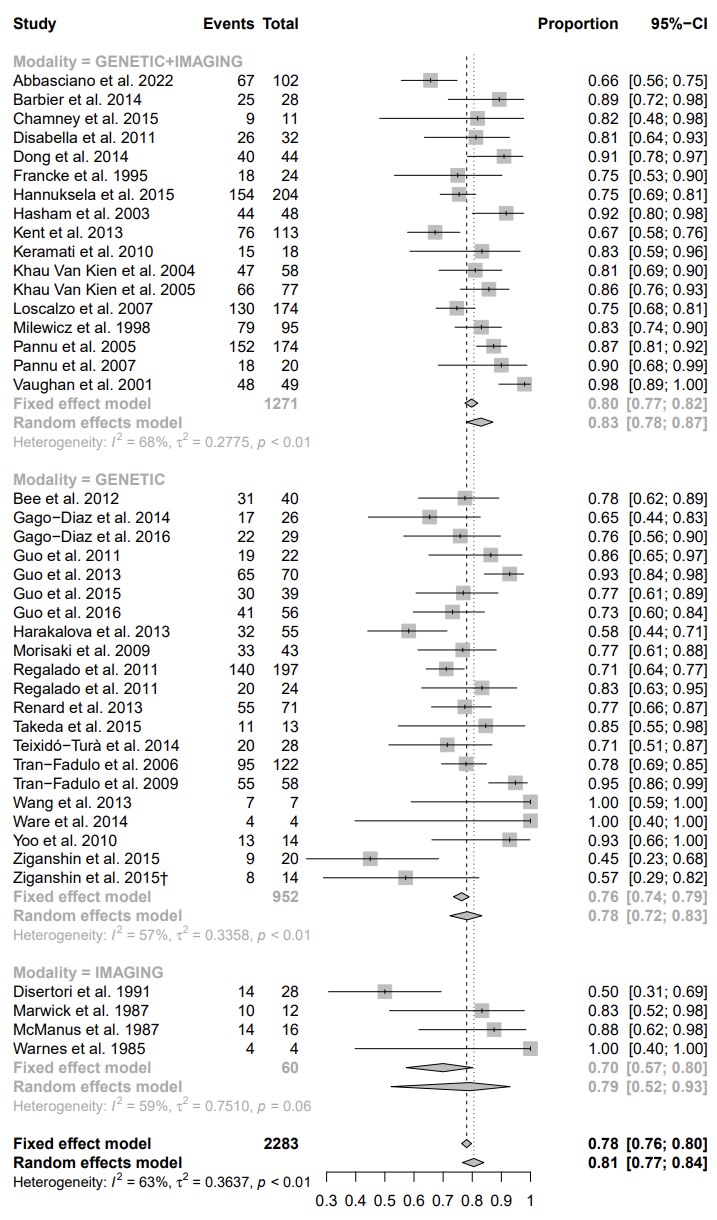

The forest plots report the results from the proportional meta-analysis of observational studies conducted in families in which one or more relatives were diagnosed with a non-syndromic thoracic aortic disease. The different studies (grouped based on the testing modality) are compared in terms of uptake of the screening offer, and the results are pooled both in a fixed effect and random effect model. *CI – Confidence Interval*

### eFigure 4 - Forest Plot describing the role of different genetic screening modalities on diagnosis in non-syndromic thoracic aortic diseases

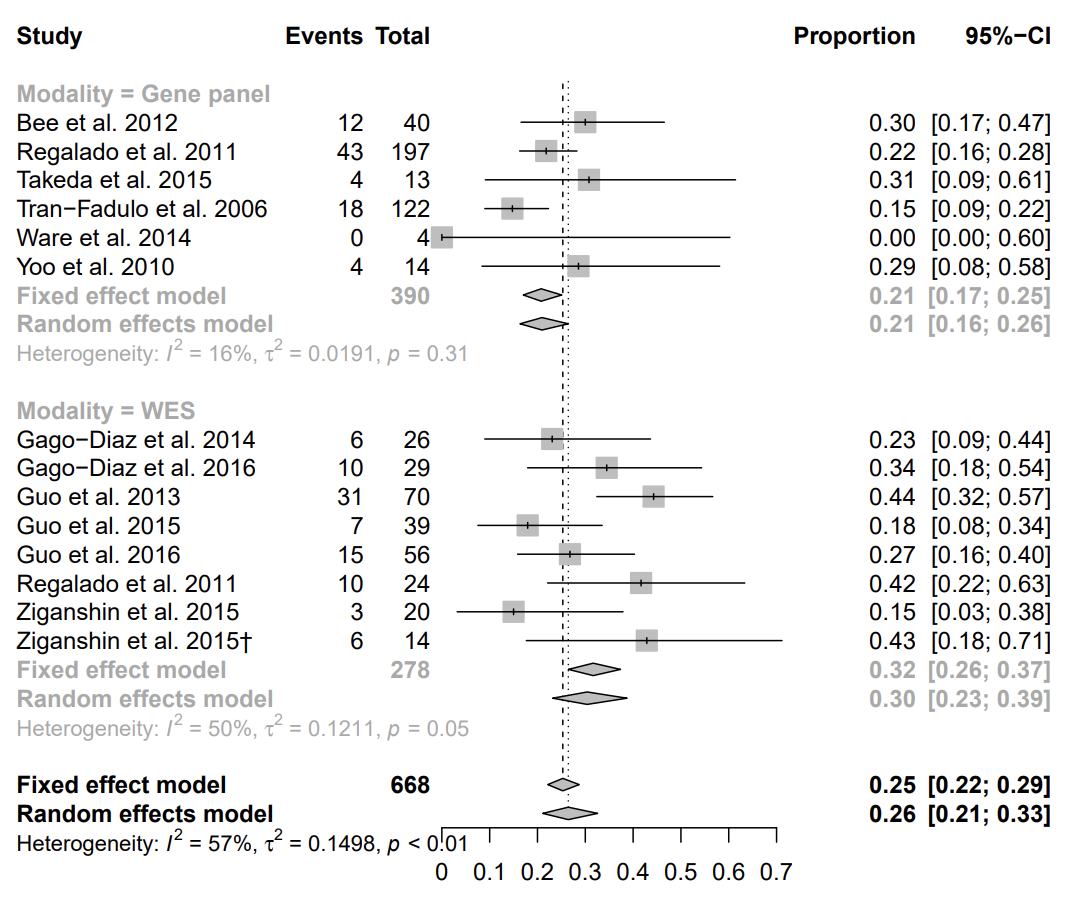

The forest plots report the results from the proportional meta-analysis of observational studies conducted in families in which one or more relatives were diagnosed with a non-syndromic thoracic aortic disease. The different studies (grouped based on the testing modality, gene panel or whole exome sequencing) are compared in terms of number of diagnoses, and the results are pooled both in a fixed effect and random effect model. *CI – Confidence Interval; WES – whole exome sequencing.*

### eFigure 5 - Forest Plot describing the role of different genetic screening modalities on uptake in non-syndromic thoracic aortic diseases

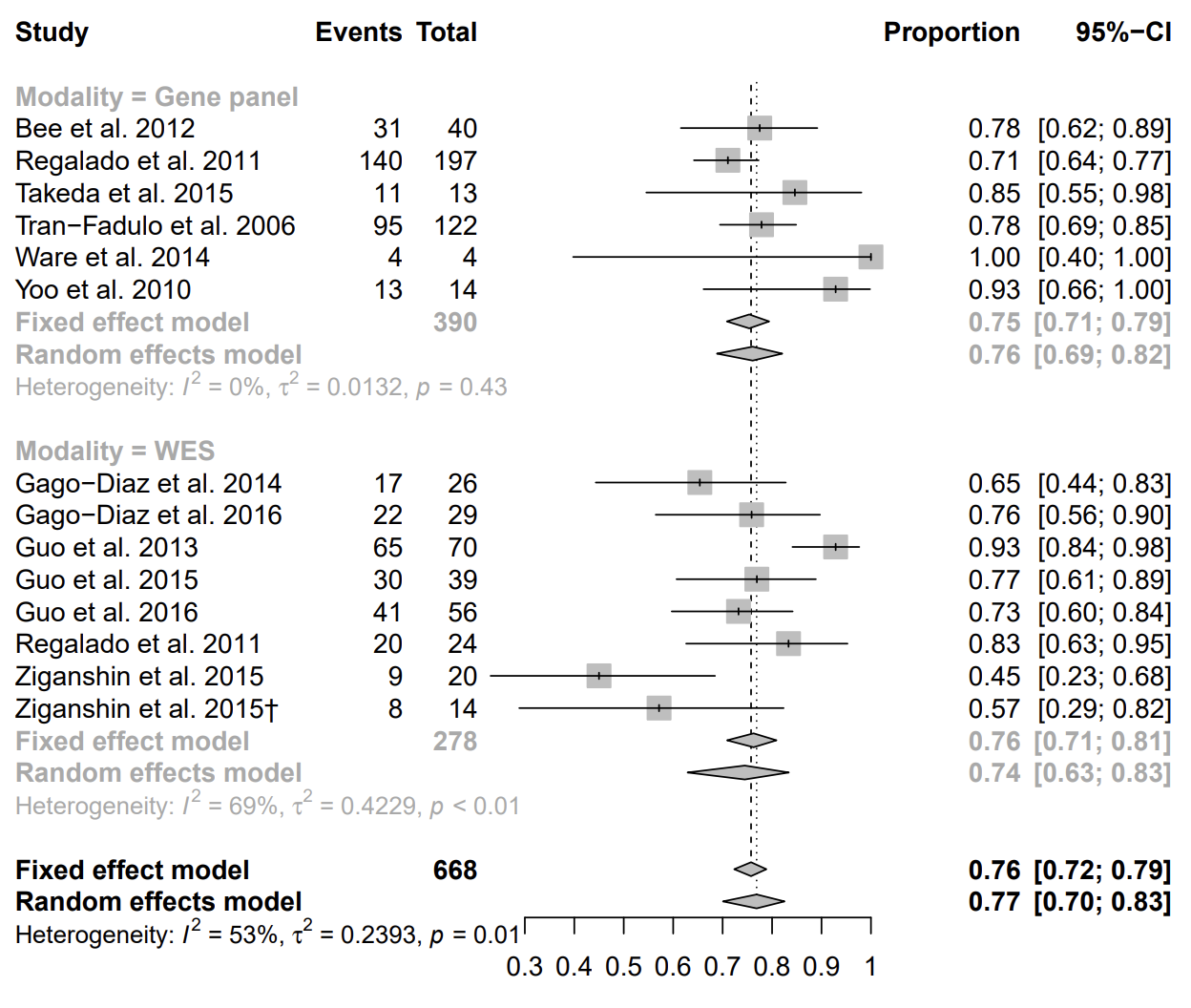

The forest plots report the results from the proportional meta-analysis of observational studies conducted in families in which one or more relatives were diagnosed with a non-syndromic thoracic aortic disease. The different studies (grouped based on the testing modality, gene panel or whole exome sequencing) are compared in terms of uptake of the screening offer, and the results are pooled both in a fixed effect and random effect model. *CI – Confidence Interval; WES – whole exome sequencing.*

### eFigure 6 - Risk of Bias plot for the individual trials

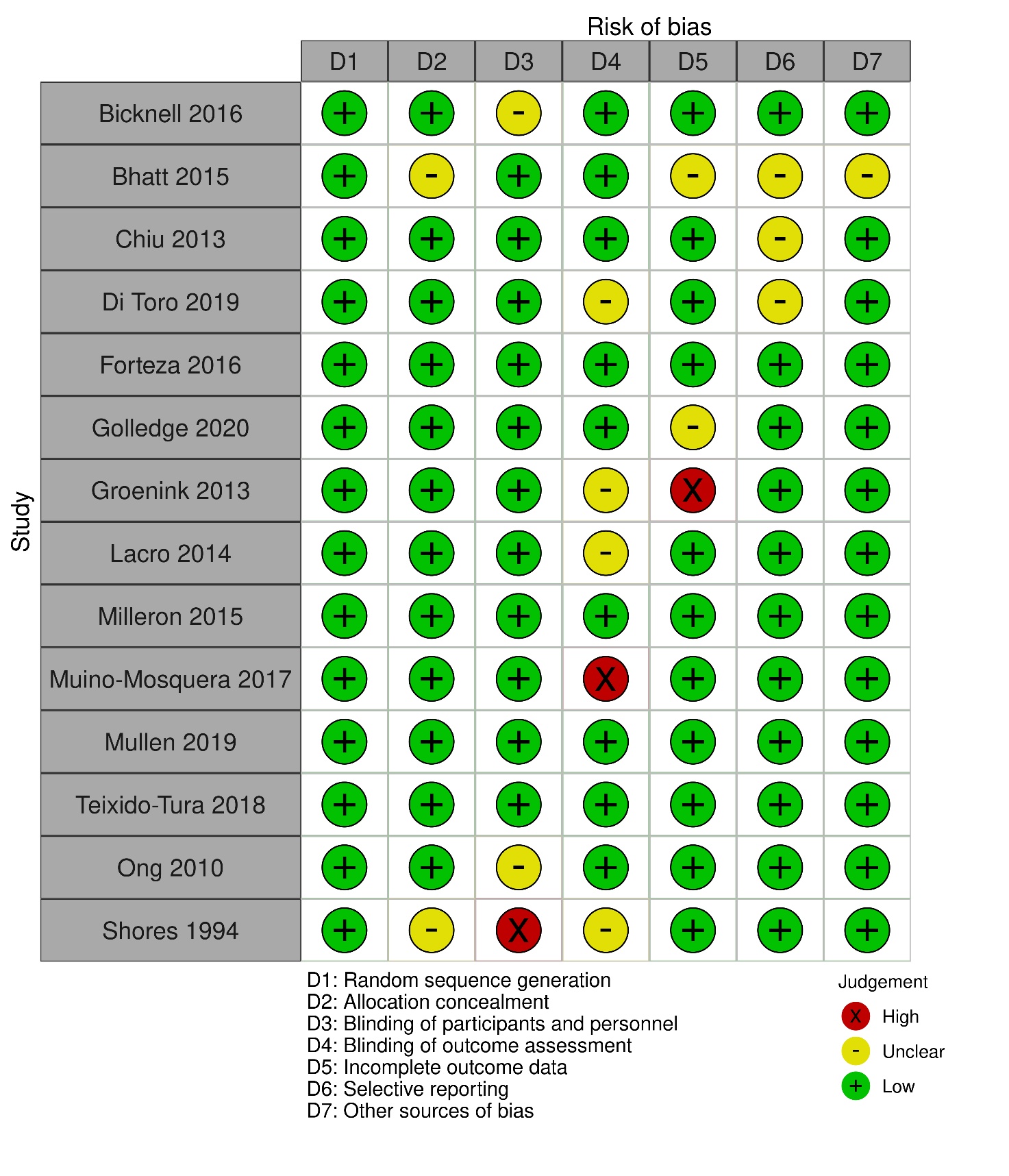

Plot reporting the risk of bias assessment for the individual trials included in the evidence synthesis exercise, according to Cochrane’s risk of bias tool, version 1 (Higgins JP, Altman DG, Gøtzsche PC, Jüni P, Moher D, Oxman AD, Savović J, Schulz KF, Weeks L, Sterne JA. The Cochrane Collaboration’s tool for assessing risk of bias in randomised trials. Bmj. 2011 Oct 18;343.).

Plot produced the robvis tool (McGuinness, LA, Higgins, JPT. Risk-of-bias VISualization (robvis): An R package and Shiny web app for visualizing risk-of-bias assessments. Res Syn Meth. 2020; 1- 7. https://doi.org/10.1002/jrsm.1411)

### eFigure 7 - Risk of Bias summary for individual trials included in the consensus exercise

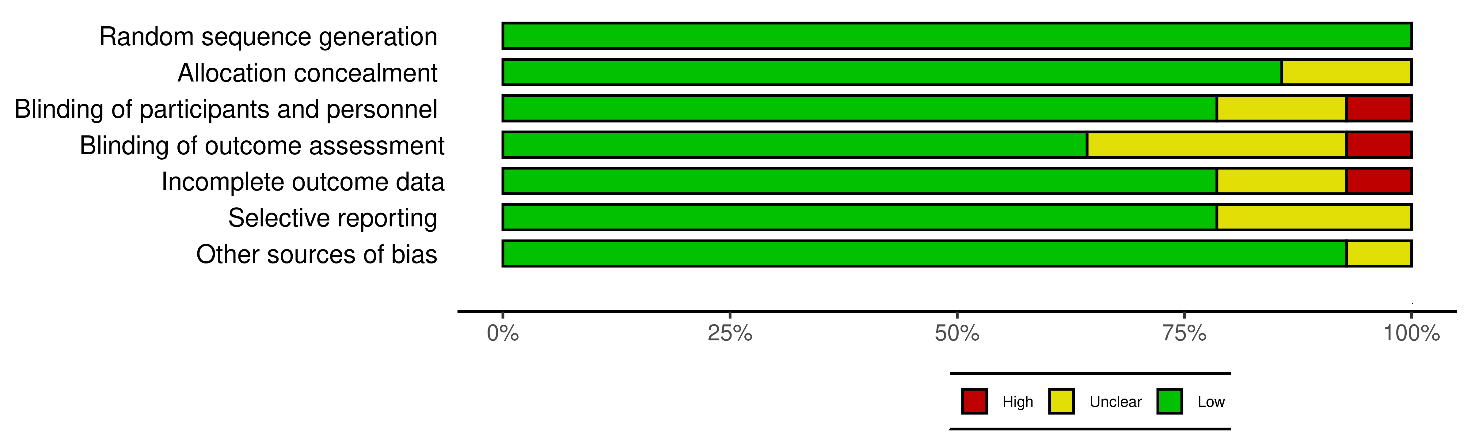

Plot summarising the overall risk of bias for the trials included in the evidence synthesis exercise, for the domains considered in Cochrane’s risk of bias tool, version 1 (Higgins JP, Altman DG, Gøtzsche PC, Jüni P, Moher D, Oxman AD, Savović J, Schulz KF, Weeks L, Sterne JA. The Cochrane Collaboration’s tool for assessing risk of bias in randomised trials. Bmj. 2011 Oct 18;343.).

Plot produced the robvis tool (McGuinness, LA, Higgins, JPT. Risk-of-bias VISualization (robvis): An R package and Shiny web app for visualizing risk-of-bias assessments. Res Syn Meth. 2020; 1- 7. https://doi.org/10.1002/jrsm.1411)

### eFigure 8 - Forest Plot describing the effect of ARB on mortality in TAD

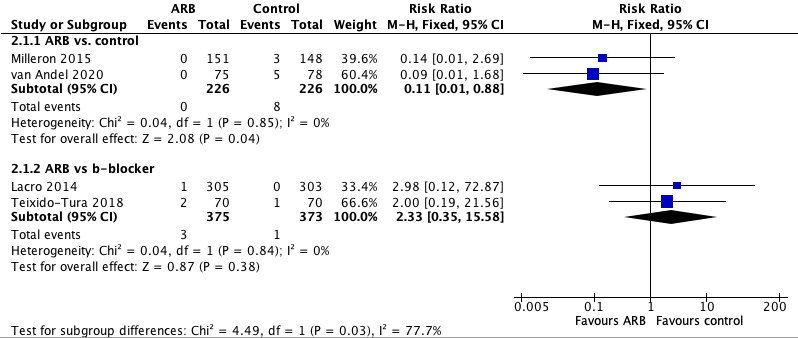

The forest plots report the results from the meta-analysis of randomised controlled trials conducted in patients affected by thoracic aortic disease. The different studies (grouped based on the comparison, control or beta-blocker) are compared in terms of mortality, and the results are pooled a fixed effect model. *ARB – Angiotensin Receptor Blocker; CI – Confidence Interval; TAD – Thoracic Aortic Disease.*

### eFigure 9 - Forest Plot describing the effect of ARB on incidence of acute aortic syndromes in TAD

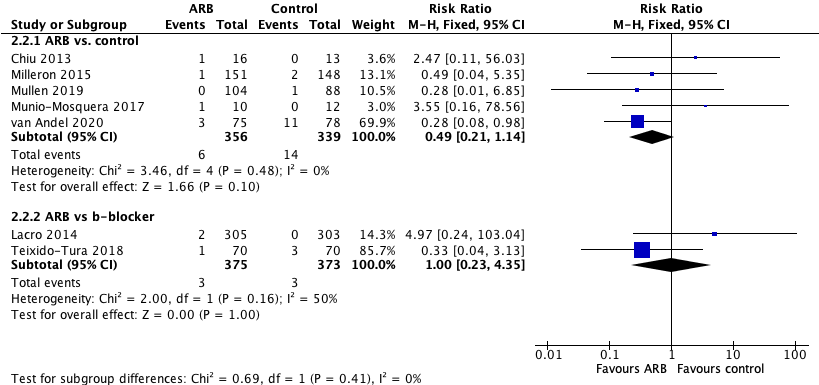

The forest plots report the results from the meta-analysis of randomised controlled trials conducted in patients affected by thoracic aortic disease. The different studies (grouped based on the comparison, control or beta-blocker) are compared in terms of incidence of acute aortic syndrome, and the results are pooled a fixed effect model. *ARB – Angiotensin Receptor Blocker; CI – Confidence Interval; TAD – Thoracic Aortic Disease.*

### eFigure 10 - Forest Plot describing the effect of ARB on need for surgery in TAD

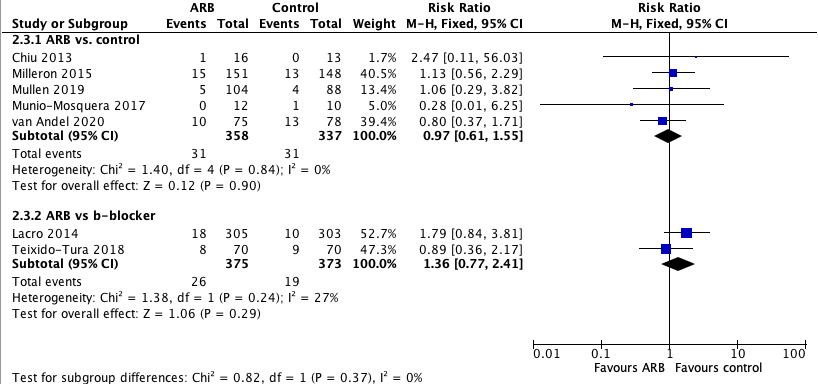

The forest plots report the results from the meta-analysis of randomised controlled trials conducted in patients affected by thoracic aortic disease. The different studies (grouped based on the comparison, control or beta-blocker) are compared in terms of incidence of acute aortic syndrome, and the results are pooled a fixed effect model. *ARB – Angiotensin Receptor Blocker; CI – Confidence Interval; TAD – Thoracic Aortic Disease.*

### eFigure 11- Forest Plot describing the effect of ARB on rate of change in TAD

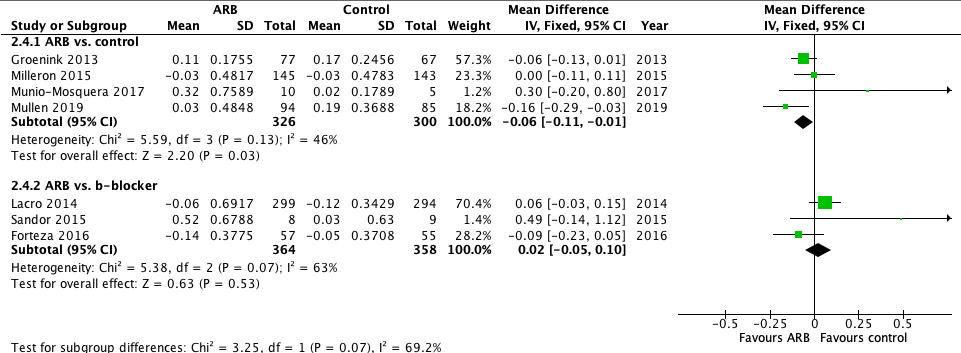

The forest plots report the results from the meta-analysis of randomised controlled trials conducted in patients affected by thoracic aortic disease. The different studies (grouped based on the comparison, control or beta-blocker) are compared in terms of rate of change (measured as body surface area-adjusted aortic root dimension Z score at the sinuses of Valsalva), and the results are pooled as mean difference in a fixed effect model. *ARB – Angiotensin Receptor Blocker; CI – Confidence Interval; TAD – Thoracic Aortic Disease.*

### eFigure 12 - Forest Plot describing the effect of B-blocker on mortality in TAD

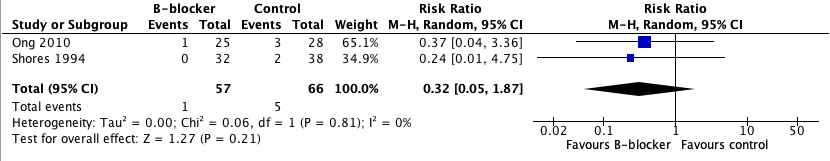

The forest plots report the results from the meta-analysis of randomised controlled trials conducted in patients affected by thoracic aortic disease. The different studies are compared in terms of rate of mortality, and the results are pooled as risk ratio in a random effect model. *CI – Confidence Interval; TAD – Thoracic Aortic Disease.*

### eFigure 13 - Forest Plot describing the effect of B-blocker on incidence of acute aortic syndromes in TAD

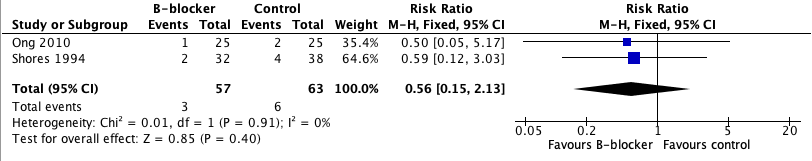

The forest plots report the results from the meta-analysis of randomised controlled trials conducted in patients affected by thoracic aortic disease. The different studies are compared in terms of rate of incidence of acute aortic syndromes, and the results are pooled as risk ratio in a random effect model. *CI – Confidence Interval; TAD – Thoracic Aortic Disease.*

### eFigure 14 - Forest Plot describing the effect of B-blocker on disease progression in TAD

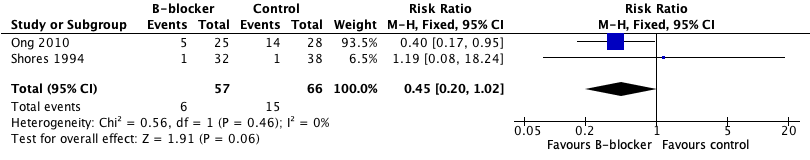

The forest plots report the results from the meta-analysis of randomised controlled trials in patients affected by thoracic aortic disease. The different studies are compared in terms of rate of disease progression (defined as composite of cardiac or arterial events; or aortic root>6cm), and the results are pooled as risk ratio in a random effect model. *CI – Confidence Interval; TAD – Thoracic Aortic Disease.*

### eTable 1. Results of Survey in Probands and Relatives.

| **Age** (Years) Mean (SD) | 58.4 (9.17) | 42.5 (31.25-57.6) |
| --- | --- | --- |
| **Age (Years) categorised** |  |  |
| Under 20 |  | 11 (6.5) |
| 20-30 |  | 28 (16.5) |
| 30-40 | 2 (2.8) | 41 (24.1) |
| 40-50 | 11 (15.5) | 29 (17.1) |
| 50-60 | 29 (40.8) | 28 (16.5) |
| 60-70 | 22 (31.0) | 22 (12.9) |
| 70+ | 7 (9.9) | 11 (6.5) |
| **Sex** |  |  |
| Male | 36 (50.7) | 68 (39.8) |
| Female | 35 (49.3) | 103 (60.2) |
| **Ethnicity** |  |  |
| White | 69 (97.2) | 165 (95.9) |
| Black | 1 (1.4) | 1 (0.6) |
| Asian | 0 (0.0) | 1 (0.6) |
| Chinese | 1 (1.4) | 0 (0.0) |
|  |  | 5 (2.9) |
| **Region** |  |  |
| Cymru/Wales | 4 (5.9) | 7 (4.1) |
| East | 5 (7.4) | 8 (4.7) |
| East Midlands | 7 (10.3) | 17 (10.1) |
| London | 7 (10.3) | 18 (10.7) |
| North East | 2 (2.9) | 6 (3.6) |
| North west | 4 (5.9) | 19 (11.2) |
| Scotland | 2 (2.9) | 4 (2.4) |
| South East | 13 (19.1) | 7 (4.1) |
| South West | 13 (19.1) | 35 (20.7) |
| West Midlands | 3 (4.4) | 24 (14.2) |
| Yorkshire and the Humber | 8 (11.8) | 11 (6.5) |
| **Probands Only** |  |  |
| Time since aortic dissection |  |  |
| One year | 17 (23.9) |  |
| Two years | 7 (9.9) |  |
| Three years | 12 (16.9) |  |
| More than three years | 35 (49.3) |  |
| **Diagnosed with syndromic condition** |  |  |
| Yes | 6 (8.5) |  |
| No | 65 (91.5) |  |
| **Had a relative diagnosed with syndromic condition** |  |  |
| Yes | 6 (8.5) |  |
| No | 58 (81.7) |  |
| Don’t know | 7 (9.9) |  |
| Received genetic testing |  |  |
| No, but was offered it | (2.9) |  |
| No, wasn’t offered it | 34 (49.3) |  |
| Yes | 33 (47.8) |  |
| **Relatives Only** |  |  |
| **Relation to person with dissection** |  |  |
| First degree |  | 154 (89.0) |
| Second degree |  | 16 (9.3) |
| Other |  | 3 (1.7) |
| **Diagnosed with syndromic condition** |  |  |
| Yes |  | 5 (3.1) |
| No |  | 159 (97.0) |
| **Received genetics appointment** |  |  |
| Yes |  | 15 (9.2) |
| No |  | 143 (87.7) |
| Not sure/ Don’t want to say |  | 5 (3.1) |

Out of 42 patients who were under 60 at the time of filling in the survey 24(57.1%) received genetics screening. Out of 55 patients who were under 65 at the time of filling in the survey 28 (50.9%) received genetic screening.

### eTable 2. Summary of discussions from Focus Groups

|  | Focus Group (WP 1) | Focus Group (WP 4) |
| --- | --- | --- |
| The Unified Theory of Acceptance and Use of Technology framework focuses on understanding users' acceptance and adoption of technology. | | |
| **Performance Expectancy.** This concept deals with the belief that using the system or technology will help improve performance. | In this case, the performance can be interpreted as the overall medical outcome of the patients and their family members. Many participants believe in the effectiveness of genetic testing in understanding why a thoracic aortic dissection (TAD) had happened to them or a family member. The main benefit identified was protecting family members, particularly children, from potential risks. They believe the process can reveal the genetic susceptibility of TAD, enabling preventive measures and early treatments. | In this context, participants expressed that they saw the benefits of using a Decision Support Tool (DST) with technology, such as having all the information in one place, linking to reliable resources, and the ability to access the tool at any time. However, there were concerns about the clarity of the information provided, and if misinterpreted, could lead to misunderstandings. |
| **Effort Expectancy:** This refers to the perceived ease of use of the technology. | In the context of this study, participants found accessing genetic screening challenging and confusing. GPs often lacked knowledge about TAD and genetic testing, leading to difficulties in understanding and navigating the process. They struggled with obtaining access to screening, understanding what was being tested, and knowing what timelines to expect. Therefore, the perceived effort of using this technology is high, indicating that improvements need to be made to simplify the process and make it more user-friendly. | While some found apps difficult to use due to health conditions, others had no issues with accessing content online. They preferred the DST in multiple formats, ideally digital. However, concerns about digital literacy and technology access, particularly for older patients and non-English speakers, were identified. Solutions such as making the tool multilingual, having leaflets, and allowing information download were suggested. |
| **Social Influence:** This term deals with the degree to which users perceive that important others (like family, friends, colleagues) believe they should use the new technology. | In this study, the participants were in favour of genetic screening, but some of their family members declined the testing due to worries about the impact of a positive test on employment and insurance, or not wanting the knowledge of their risk to affect their life. These instances illustrate the role of social influence in the decision to undertake genetic testing. | The participants were generally open to recommending the DST to family members, but there were mixed feelings about whether family members would accept genetic screening or use the DST. Participants agreed that clinician endorsement of the DST might hold more authority and credibility. |
| **Facilitating Conditions:** This concept refers to users' beliefs that an organisational and technical infrastructure exists to support the use of the system. | In this context, many participants found a lack of facilitating conditions such as GP's knowledge about aortic dissection and genetic screening, which hindered them from accessing genetic testing smoothly. A tool like the DST would only be beneficial if barriers to genetic testing, like lack of knowledge and coordination among healthcare providers, are removed. | Participants appreciated the potential convenience and accessibility of the DST but also expressed concerns about data security and the potential misuse of personal information. They emphasised the need for robust data protection measures and transparency about how data is used. |
| **Hedonic Motivation:** This usually refers to the fun or pleasure derived from using the technology. . It doesn't seem to be directly applicable to this scenario as genetic testing is a serious matter rather than a source of pleasure or fun. However, some participants may derive a sense of relief or satisfaction from taking proactive steps to protect their family's health, which could be considered a form of hedonic motivation. |  | While not explicitly mentioned in the text, the participants' enthusiasm for having all information in one place and the potential for the DST to contain a wide array of helpful resources suggests some level of hedonic motivation. |
| **Price Value:** This refers to users' cognitive trade-off between the perceived benefits and the monetary cost of using the technology. In this case, price value could be considered in terms of the potential cost savings to the healthcare system. | One participant noted the potential cost-saving to the NHS of identifying and monitoring people at risk of TAD rather than treating an emergency dissection, indicating a perception of high price value. | While the text doesn't mention the price explicitly, one participant suggested that the required technology (like an iPad) should be made available within the hospital, implying a consideration of cost. |
| **Habit:** This refers to the extent to which people tend to perform behaviours automatically because of learning. This theme does not appear to be directly relevant to this study, as genetic testing for TAD is not a regular or routine behaviour. However, participants' actions of researching into TAD and genetic screening themselves, or consistently pushing to receive the screening, could indicate the formation of a habit to actively manage their health conditions. |  | The text doesn't provide enough information to infer about the habitual use of the DST. However, participants mentioned that they were generally willing to use technology, suggesting they could easily incorporate the DST into their routines. |
| Theory of Planned Behaviour / Implementation Intentions: The Theory of Planned Behaviour (TPB) posits that human action is guided by three kinds of considerations: behavioural beliefs (attitudes), normative beliefs (subjective norms), and control beliefs (perceived behavioural control). | | |
| **Attitudes (Behavioural Beliefs):** | The attitudes towards genetic testing among the participants were generally positive, seeing it as a way to protect family members and understand why a thoracic aortic dissection (TAD) had happened to them. The main benefit identified by participants was the potential to protect family members, particularly children. Some participants didn't see any risks, indicating a favourable attitude towards genetic testing. | Participants in general held positive attitudes towards the use of a Decision Support Tool (DST) with technology. They appreciated the benefits such as having all the information in one place, easy access to additional resources, and the flexibility of accessing it at any time. Some concerns were raised, particularly around digital literacy, language barriers, and data privacy, but these didn't negate the overall positive attitude. |
| **Subjective Norms (Normative Beliefs):** The norms in this context could be defined by the perceptions of family members, GPs, and society. | Some participants experienced family members declining the testing due to potential impacts on insurance and employment, and fears about knowing they're at risk, indicating some societal and familial pressures against testing. On the other hand, some participants found themselves being the source of information for other family members, indicating a possible social role in promoting genetic testing. | The perception of others' attitudes was varied. Most participants felt comfortable recommending the DST to family members, indicating they believe their family members would approve of its use. However, some participants felt their family members would be resistant to using the DST, especially those who had declined genetic screening. |
| **Perceived Behavioural Control:** This refers to individuals' perceptions of their ability to perform a given behaviour. | In this context, participants found it difficult to access genetic screening, with many expressing confusion about the process and experiencing difficulties in navigation. The lack of knowledge about TAD and the genetic screening process among GPs often created barriers, leading to perceived low behavioural control. | Participants' perceived ability to use the DST varied, with some expressing difficulty due to health conditions and others preferring certain types of technology. There were also concerns about older patients and non-English speakers struggling to use the DST, indicating perceived barriers to its use. However, solutions such as multi-language support and downloadable information were suggested to improve this control. |
| **Implementation Intentions** refer to the individual's plan specifying the "when, where and how" to perform the intended behaviour. In this context, implementation intentions could be viewed in terms of the specific steps the participants took or planned to take to access genetic screening. | Participants were proactive in pushing for genetic screening, indicating clear intentions to implement the behaviour. Some participants even identified specific individuals (such as a specialist aortic nurse or a geneticist) they'd prefer to discuss the decision support tool (DST) with, suggesting a plan for how to carry out their intentions. However, some participants expressed a need for more clear information about the pathway to genetic testing, indicating that the plan to implement was not fully formed due to lack of information.  There was also a variation in "when" the participants believed genetic testing should be carried out - some believed it should be offered as soon as possible, while others felt that they did not feel able to consider genetic testing for several months after experiencing a TAD, reflecting different implementation intentions based on personal circumstances. | **When**: Participants expressed that the DST should be accessible at any time as patients and families may be ready to consider genetic screening at different points after dissection.  **Where**: There was a preference for the DST to be available in a digital format (like an app or online platform), but also support for the tool to be available in multiple formats for those who might struggle with technology.  **How**: There was a range of opinions on how the DST should be used. Some participants felt that personal data should be input into the DST, while others were more concerned about privacy. There was a general consensus that any data input should be secure and the purpose of the data collection should be transparent. It was also suggested that clinicians could play a role in checking a user's understanding after using the DST. |
| Health literacy, including accessing, understanding, appraising, and making decisions regarding health information. Additionally, considering the themes related to health promotion, health prevention, and healthcare. Here are the themes identified from these perspectives: | | |
| **Accessing Health Information** | Many participants struggled to access accurate and useful information about genetic testing and TAD. They often faced confusion, with GPs lacking knowledge and providing unclear directions for accessing screening. In terms of health promotion and prevention, this indicates a need for improved resources and protocols to facilitate access to information about genetic testing and TAD. | Participants generally had access to technology and could access digital platforms. However, some participants mentioned concerns about equitable access due to factors like digital literacy, language barriers, or disabilities. It was suggested to provide the information in multiple formats and languages to ensure accessibility. |
| **Understanding Health Information** | The participants demonstrated a wide range of abilities to understand the information they accessed. They often had to conduct their own research to supplement the lack of information provided by their GPs. However, this varied widely as the participants' health literacy levels were likely higher than average TAD patients due to being recruited from ADAUK and agreeing to participate in the focus group sessions. | The understanding of health information was varied among participants. They appreciated the benefits of having all information in one place and easily accessible. However, concerns were raised about potential misinterpretation of information, and it was suggested that clinicians could help in checking the user's understanding after using the DST. |
| **Appraising Health Information** | Participants often had to judge the validity and relevance of information themselves, especially due to lack of understanding from their GPs about aortic dissection and the need for genetic screening. This was also seen in the participants' discussions about the benefits and risks of genetic testing, where they had to evaluate the pros and cons themselves. | Participants showed an ability to evaluate the advantages and disadvantages of using a DST. They highlighted the importance of trust, especially when the DST provides links to external resources or requires the input of personal data. |
| **Making Decisions Regarding Health Information** | Most participants were in favour of genetic testing and made the decision to pursue it, even amidst barriers. This shows a high level of engagement in making health decisions. However, their ability to make informed decisions was compromised by difficulties in accessing, understanding, and appraising relevant information. This highlights the need for improved communication and support from healthcare providers. | Most participants were open to using a DST and making decisions about genetic screening. However, there were concerns about data privacy, which could influence decision-making. Participants emphasised the need for transparent data collection and usage practices. |
| **Health Promotion** | Health promotion was a recurring theme, as genetic testing was seen as a tool for promoting the health of family members, particularly children. Participants saw the value in understanding their risk to potentially prevent adverse outcomes. However, a lack of standardised care and lack of coordination across specialties hampered this. | Participants viewed the DST as a tool that could promote health. By providing valuable information about genetic screening, it can help them understand their risks and potentially prevent adverse outcomes. |
| **Health Prevention** | Genetic testing for TAD was largely viewed as a preventive measure, allowing those at risk to be monitored and potentially prevent the occurrence of aortic dissection. Participants also noted the potential cost savings for the NHS if people at risk could be identified and monitored rather than treated after an emergency dissection. | The DST was seen as a preventive measure, enabling users to access and understand information about genetic screening, which could potentially prevent aortic dissection if the risk is detected early. |
| **Healthcare** | Concerns about the healthcare system were raised, including a lack of standardisation in patient care for TAD, a lack of knowledge and awareness among GPs, and a lack of coordination of care across specialties. These concerns affected the participants' access to, understanding of, and ability to appraise health information, and thus their capacity to make informed health decisions. | Participants raised concerns about the healthcare system, such as the lack of knowledge among GPs, lack of standardisation in patient care for TAD, and poor coordination across specialties. The DST was seen as a potential solution to these issues by providing reliable information, promoting better understanding of TAD and genetic screening, and facilitating better communication and support from healthcare providers. There were also suggestions for integrating the DST within the existing healthcare infrastructure, like making it accessible within hospitals. |

### eTable 3 - Risk of Bias summary for observational studies included in the consensus exercise

| **Study (Author/Year)** | **Newcastle-Ottawa Scale** | | |
| --- | --- | --- | --- |
|  | **Selection** | **Comparability** | **Outcome** |
| Abbasciano et al. 2022 | ☆☆☆ | ☆☆☆ | ☆☆ |
| Barbier et al. 2014 | ☆☆ | ☆☆ | ☆ |
| Bee et al. 2012 | ☆ | ☆☆ | ☆☆☆ |
| Chamney et al. 2015 | ☆ | ☆ | ☆☆ |
| Disabella et al. 2011 | ☆☆☆ | ☆☆ | ☆☆☆ |
| Disertori et al. 1991 | - | - | ☆ |
| Dong et al. 2014 | ☆☆☆ | ☆☆ | ☆☆☆ |
| Francke et al. 1995 | ☆☆ | ☆ | ☆☆☆ |
| Gago-Diaz et al. 2014 | ☆☆☆ | ☆☆ | ☆☆☆ |
| Gago-Diaz et al. 2016 | ☆☆ | ☆☆ | ☆☆ |
| Guo et al. 2001 | ☆ | ☆ | ☆☆☆ |
| Guo et al. 2007 | ☆☆ | ☆ | ☆☆☆ |
| Guo et al. 2009 | ☆☆☆ | ☆ | ☆☆☆ |
| Guo et al. 2011 | ☆☆☆ | ☆ | ☆☆☆ |
| Guo et al. 2013 | ☆☆☆ | ☆ | ☆☆☆ |
| Guo et al. 2015 | ☆☆☆ | ☆ | ☆☆☆ |
| Guo et al. 2016 | ☆☆☆ | ☆ | ☆☆ |
| Hannuksela et al. 2015 | ☆☆☆ | ☆ | ☆☆☆ |
| Hannuksela et al. 2016 | ☆☆☆ | ☆☆ | ☆☆ |
| Harakalova et al. 2013 | ☆☆☆ | ☆☆ | ☆☆☆ |
| Hasham et al. 2003 | ☆☆☆ | ☆☆ | ☆☆☆ |
| Kakko et al. 2003 | ☆☆☆ | ☆☆ | ☆☆ |
| Kent et al. 2013 | ☆☆ | ☆☆ | ☆☆☆ |
| Keramati et al. 2010 | ☆☆ | ☆ | ☆☆ |
| Khau Van Kien et al. 2004 | ☆☆☆ | ☆☆ | ☆☆☆ |
| Khau Van Kien et al. 2005 | ☆☆☆ | ☆☆ | ☆☆☆ |
| Kuang et al. 2016 | ☆☆☆ | ☆☆ | ☆ |
| Loscalzo et al. 2007 | ☆☆☆ | ☆ | ☆☆☆ |
| Marwick et al. 1987 | - | ☆ | ☆ |
| McManus et al. 1987 | - | ☆ | ☆☆ |
| Milewicz et al. 1998 | ☆☆ | ☆ | ☆☆ |
| Morisaki et al. 2009 | ☆☆ | ☆ | ☆ |
| Pannu et al. 2005 | ☆☆☆ | ☆☆ | ☆ |
| Pannu et al. 2007 | ☆☆☆ | ☆☆ | ☆☆☆ |
| Regalado et al. 2011 | ☆☆ | ☆☆ | ☆ |
| Regalado et al. 2011 | ☆☆ | ☆☆ | ☆☆ |
| Regalado et al. 2011 | ☆☆ | ☆☆ | ☆☆ |
| Renard et al. 2013 | ☆☆ | ☆☆ | ☆☆ |
| Robertson et al. 2016 | ☆☆☆ | ☆☆ | ☆☆☆ |
| Sherrah et al. 2016 | ☆☆☆ | ☆☆ | ☆☆☆ |
| Takeda et al. 2015 | ☆☆ | ☆☆ | ☆☆ |
| Teixidó-Turà et al. 2014 | ☆ | ☆ | ☆ |
| Tortora et al. 2017 | * | * | * |
| Tran-Fadulo et al. 2006 | ☆☆ | ☆ | ☆☆ |
| Tran-Fadulo et al. 2009 | ☆☆ | ☆☆ | ☆☆ |
| Vaughan et al. 2001 | ☆☆☆ | ☆☆ | ☆☆☆ |
| Wang et al. 2010 | ☆☆ | ☆ | ☆☆ |
| Wang et al. 2013 | ☆☆ | ☆☆ | ☆☆ |
| Ware et al. 2014 | ☆ | ☆ | ☆ |
| Warnes et al. 1985 | - | ☆ | ☆ |
| Weigang et al. 2007 | ☆☆☆ | ☆☆ | ☆☆ |
| Yoo et al. 2010 | ☆☆ | ☆☆ | ☆☆ |
| Zhu et al. 2006 | ☆☆☆ | ☆☆ | ☆☆ |
| Ziganshin et al. 2015 | ☆☆ | ☆ | ☆ |

The table reports the risk of bias judgements for the observational studies assessing screening modalities included in the evidence synthesis exercise, for the domains considered in the Newcaste-Ottawa scale (Wells G, Shea B, O'Connell D, Peterson j, Welch V, Losos M, et al. The Newcastle–Ottawa Scale (NOS) for Assessing the Quality of Non-Randomized Studies in Meta-Analysis, 2000).

For observational studies conducted before 2018, the ratings expressed in the review by Mariscalco et al. (Mariscalco G, Debiec R, Elefteriades JA, Samani NJ, Murphy GJJJotAHA. Systematic review of studies that have evaluated screening tests in relatives of patients affected by nonsyndromic thoracic aortic disease. 2018;7(15):e009302.) have been adopted.

### eTable 4 – Evidence-to-decision framework for the PICO “Should imaging tests in FDR vs. imaging tests in SDR be used for screening for aortopathy?”

The table below reports the GUIDE evidence-to-decision process to discuss the PICO related to cascade screening for imaging test vs extending the imaging screening to second degree relative (SDR).

| Question | |
| --- | --- |
| **Should imaging tests in FDR vs. imaging tests in SDR be used for screening for aortopathy?** | |
| **Population:** | screening for aortopathy |
| **Intervention:** | imaging tests in FDR |
| **Comparison:** | imaging tests in SDR |
| **Main outcomes:** | Quality of life; Uptake of Screening offered (events for patients not screened); Diagnosis rate with imaging; Diagnosis rate with imaging; Psychometric Testing; |
| **Setting:** |  |
| **Perspective:** |  |
| **Background:** |  |
| **Conflict of interests:** |  |

Assessment

| Problem  Is the problem a priority? | | |
| --- | --- | --- |
| Judgement | Research evidence | Additional considerations |
| ○ No ○ Probably no ● Probably yes ○ Yes ○ Varies ○ Don't know | **Outcomes** \| **Anticipated absolute effects^*^ (95% CI)** \| \| **Relative effect (95% CI)** \| **№ of participants (studies)** \| **Certainty of the evidence (GRADE)** \| **Comments**; \| **Risk with imaging tests in SDR** \| **Risk with imaging tests in FDR** \| \| \| \|; Quality of life \| One study examined the mean scores between FDR and second degree relatives in terms of physical functioning, role limitation due to physical health, emotional problmes, as well as impact on energy/fatigue, emotional wellbeing, social function, bodily pain and general health and found no difference between FDR and SDR in terms of these domains. \| \| - \| (1 observational study) \| ⨁⨁◯◯ Low \|; Uptake of Screening offered (events for patients not screened) \| Study population \| \| **OR 0.42** (0.14 to 1.33) \| 60 (4 observational studies) \| ⨁◯◯◯ Very low^a,b^ \|; \| 364 per 1,000 \| **194 per 1,000** (74 to 432) \| \| \| \|; Diagnosis rate with imaging \| Study population \| \| **RR 1.37** (0.35 to 5.41) \| 53 (3 observational studies) \| ⨁◯◯◯ Very low^a^ \|; \| 103 per 1,000 \| **142 per 1,000** (36 to 560) \| \| \| \|; Diagnosis rate with imaging \| Study population \| \| **OR 1.42** (0.32 to 6.27) \| 53 (3 observational studies) \| ⨁◯◯◯ Very low^a,b^ \|; \| 103 per 1,000 \| **141 per 1,000** (36 to 420) \| \| \| \|; Psychometric Testing \| One study examined the psychometric testing between PHQ and GAD scores between FDR and SDR and found no difference between the two groups in this testing (PHQ p=0.088, GAD p=0.06). \| \| - \| (1 observational study) \| ⨁⨁◯◯ Low \| Rated down for inconsistent results Rated down for wide confidence intervals and low event rate | Identified during Delphi exercise    Considerations of stressors on relatives in being screened. |
| Desirable Effects  How substantial are the desirable anticipated effects? | | |
| Judgement | Research evidence | Additional considerations |
| ○ Trivial ○ Small ○ Moderate ○ Large ○ Varies ● Don't know | **Outcomes** \| **Anticipated absolute effects^*^ (95% CI)** \| \| **Relative effect (95% CI)** \| **№ of participants (studies)** \| **Certainty of the evidence (GRADE)** \| **Comments**; \| **Risk with imaging tests in SDR** \| **Risk with imaging tests in FDR** \| \| \| \|; Quality of life \| One study examined the mean scores between FDR and second degree relatives in terms of physical functioning, role limitation due to physical health, emotional problmes, as well as impact on energy/fatigue, emotional wellbeing, social function, bodily pain and general health and found no difference between FDR and SDR in terms of these domains. \| \| - \| (1 observational study) \| ⨁⨁◯◯ Low \|; Uptake of Screening offered (events for patients not screened) \| Study population \| \| **OR 0.42** (0.14 to 1.33) \| 60 (4 observational studies) \| ⨁◯◯◯ Very low^a,b^ \|; \| 364 per 1,000 \| **194 per 1,000** (74 to 432) \| \| \| \|; Diagnosis rate with imaging \| Study population \| \| **RR 1.37** (0.35 to 5.41) \| 53 (3 observational studies) \| ⨁◯◯◯ Very low^a^ \|; \| 103 per 1,000 \| **142 per 1,000** (36 to 560) \| \| \| \|; Diagnosis rate with imaging \| Study population \| \| **OR 1.42** (0.32 to 6.27) \| 53 (3 observational studies) \| ⨁◯◯◯ Very low^a,b^ \|; \| 103 per 1,000 \| **141 per 1,000** (36 to 420) \| \| \| \|; Psychometric Testing \| One study examined the psychometric testing between PHQ and GAD scores between FDR and SDR and found no difference between the two groups in this testing (PHQ p=0.088, GAD p=0.06). \| \| - \| (1 observational study) \| ⨁⨁◯◯ Low \| Rated down for inconsistent results Rated down for wide confidence intervals and low event rate | Desirable effects-  Screening uptaking  Diagnosis  Considerations- non syndromic included.  Systematic review showed increased diagnostic yield in SDR compared to cascade screening.  Would have expected a difference in screening uptake and stress in undergoing screening.  In clinical practice screening in 1st and 2nd degree relatives.  Patient partner and family partner experience: Not knowing maybe has worse impact on patients QoL. Screening is wanted by patients and relatives.  Sometimes people not wanting to screening because of modalities screening. Barries to access also a issue. |
| Undesirable Effects  How substantial are the undesirable anticipated effects? | | |
| Judgement | Research evidence | Additional considerations |
| ○ Large ○ Moderate ○ Small ○ Trivial ○ Varies ● Don't know | **Outcomes** \| **Anticipated absolute effects^*^ (95% CI)** \| \| **Relative effect (95% CI)** \| **№ of participants (studies)** \| **Certainty of the evidence (GRADE)** \| **Comments**; \| **Risk with imaging tests in SDR** \| **Risk with imaging tests in FDR** \| \| \| \|; Quality of life \| One study examined the mean scores between FDR and second degree relatives in terms of physical functioning, role limitation due to physical health, emotional problmes, as well as impact on energy/fatigue, emotional wellbeing, social function, bodily pain and general health and found no difference between FDR and SDR in terms of these domains. \| \| - \| (1 observational study) \| ⨁⨁◯◯ Low \|; Uptake of Screening offered (events for patients not screened) \| Study population \| \| **OR 0.42** (0.14 to 1.33) \| 60 (4 observational studies) \| ⨁◯◯◯ Very low^a,b^ \|; \| 364 per 1,000 \| **194 per 1,000** (74 to 432) \| \| \| \|; Diagnosis rate with imaging \| Study population \| \| **RR 1.37** (0.35 to 5.41) \| 53 (3 observational studies) \| ⨁◯◯◯ Very low^a^ \|; \| 103 per 1,000 \| **142 per 1,000** (36 to 560) \| \| \| \|; Diagnosis rate with imaging \| Study population \| \| **OR 1.42** (0.32 to 6.27) \| 53 (3 observational studies) \| ⨁◯◯◯ Very low^a,b^ \|; \| 103 per 1,000 \| **141 per 1,000** (36 to 420) \| \| \| \|; Psychometric Testing \| One study examined the psychometric testing between PHQ and GAD scores between FDR and SDR and found no difference between the two groups in this testing (PHQ p=0.088, GAD p=0.06). \| \| - \| (1 observational study) \| ⨁⨁◯◯ Low \| Rated down for inconsistent results Rated down for wide confidence intervals and low event rate | Patient partner and family partner experience: Not knowing maybe has worse impact on patients QoL. Screening is wanted by patients and relatives.  Sometimes people not wanting to screening because of modalities screening. Barries to access also a issue. |
| Certainty of evidence  What is the overall certainty of the evidence of effects? | | |
| Judgement | Research evidence | Additional considerations |
| ● Very low ○ Low ○ Moderate ○ High ○ No included studies |  |  |
| Values  Is there important uncertainty about or variability in how much people value the main outcomes? | | |
| Judgement | Research evidence | Additional considerations |
| ○ Important uncertainty or variability ○ Possibly important uncertainty or variability ● Probably no important uncertainty or variability ○ No important uncertainty or variability |  | Patient and family partners: Willing to be screened with imaging. No concerns about outcomes assessed. Quality of life value of reassurance is of high value even if clinically not as reassurance.  Extending screening- could identify those with mild dilation in early/late in life that may not be clinically relevant. Also if normal doesn’t mean no risk, but less risk. |
| Balance of effects  Does the balance between desirable and undesirable effects favor the intervention or the comparison? | | |
| Judgement | Research evidence | Additional considerations |
| ○ Favors the comparison ○ Probably favors the comparison ○ Does not favor either the intervention or the comparison ○ Probably favors the intervention ○ Favors the intervention ○ Varies ○ Don't know |  |  |
| Resources required  How large are the resource requirements (costs)? | | |
| Judgement | Research evidence | Additional considerations |
| ○ Large costs ○ Moderate costs ○ Negligible costs and savings ○ Moderate savings ○ Large savings ○ Varies ● Don't know | There are costs associated with extending the screening. Compared to cost of managing acute dissection compared to screening. No studies identified. | All imaging modalites included.  Cost benefits of different screening programs (for instance, echocardiograms compared to MRIs compared to CTs).    Formal cost and cost effectiveness studies are required to inform practice. |
| Certainty of evidence of required resources  What is the certainty of the evidence of resource requirements (costs)? | | |
| Judgement | Research evidence | Additional considerations |
| ○ Very low ○ Low ○ Moderate ○ High ● No included studies |  |  |
| Cost effectiveness  Does the cost-effectiveness of the intervention favor the intervention or the comparison? | | |
| Judgement | Research evidence | Additional considerations |
| ○ Favors the comparison ○ Probably favors the comparison ○ Does not favor either the intervention or the comparison ○ Probably favors the intervention ○ Favors the intervention ○ Varies ● No included studies |  |  |
| Equity  What would be the impact on health equity? | | |
| Judgement | Research evidence | Additional considerations |
| ○ Reduced ○ Probably reduced ○ Probably no impact ● Probably increased ○ Increased ○ Varies ○ Don't know |  | Geographical limitations are a consideration (being near centre with MRI/CT ect).  Access to referrals for screening. |
| Acceptability  Is the intervention acceptable to key stakeholders? | | |
| Judgement | Research evidence | Additional considerations |
| ○ No ○ Probably no ● Probably yes ○ Yes ○ Varies ○ Don't know |  |  |
| Feasibility  Is the intervention feasible to implement? | | |
| Judgement | Research evidence | Additional considerations |
| ○ No ○ Probably no ● Probably yes ○ Yes ○ Varies ○ Don't know |  |  |

Summary of judgements

|  | **Judgement** | | | | | | |
| --- | --- | --- | --- | --- | --- | --- | --- |
| **Problem** | No | Probably no | **Probably yes** | Yes |  | Varies | Don't know |
| **Desirable Effects** | Trivial | Small | Moderate | Large |  | Varies | **Don't know** |
| **Undesirable Effects** | Large | Moderate | Small | Trivial |  | Varies | **Don't know** |
| **Certainty of evidence** | **Very low** | Low | Moderate | High |  |  | No included studies |
| **Values** | Important uncertainty or variability | Possibly important uncertainty or variability | **Probably no important uncertainty or variability** | No important uncertainty or variability |  |  |  |
| **Balance of effects** | Favors the comparison | Probably favors the comparison | Does not favor either the intervention or the comparison | Probably favors the intervention | Favors the intervention | Varies | Don't know |
| **Resources required** | Large costs | Moderate costs | Negligible costs and savings | Moderate savings | Large savings | Varies | **Don't know** |
| **Certainty of evidence of required resources** | Very low | Low | Moderate | High |  |  | **No included studies** |
| **Cost effectiveness** | Favors the comparison | Probably favors the comparison | Does not favor either the intervention or the comparison | Probably favors the intervention | Favors the intervention | Varies | **No included studies** |
| **Equity** | Reduced | Probably reduced | Probably no impact | **Probably increased** | Increased | Varies | Don't know |
| **Acceptability** | No | Probably no | **Probably yes** | Yes |  | Varies | Don't know |
| **Feasibility** | No | Probably no | **Probably yes** | Yes |  | Varies | Don't know |

Type of recommendation

| Strong recommendation against the intervention | Conditional recommendation against the intervention | Conditional recommendation for either the intervention or the comparison | **Conditional recommendation for the intervention** | Strong recommendation for the intervention |
| --- | --- | --- | --- | --- |
| ○ | ○ | ○ | **●** | ○ |

Conclusions

Recommendation

We suggest the use of imaging screening for first and second degree relatives (conditional recommendation, very low certainty of evidence);.

Subgroup considerations

Geographical considerations to improve screening for FDR and SDR

Research priorities

Cost and cost effectiveness studies.

### eTable 5 – Evidence-to-decision framework for the PICO “Should imaging and genetic testing vs. genetic testing alone be used for screening of aortopathy?”

The table below reports the GUIDE evidence-to-decision process to discuss the PICO related to the role of imaging and genetic in screening for aortopathy.

| Question | |
| --- | --- |
| **Should imaging and genetic testing vs. genetic testing alone be used for screening of aortopathy (Non-diagnostic summary)?** | |
| **Population:** | screening of aortopathy (non-diagnostic summary) |
| **Intervention:** | imaging and genetic testing |
| **Comparison:** | genetic testing alone |
| **Main outcomes:** | Diagnosis; Uptake of Testing; Quality of life; Psychometric testing; |
| **Setting:** |  |
| **Perspective:** |  |
| **Background:** |  |
| **Conflict of interests:** |  |

Assessment

| Problem  Is the problem a priority? | | |
| --- | --- | --- |
| Judgement | Research evidence | Additional considerations |
| ○ No ○ Probably no ● Probably yes ○ Yes ○ Varies ○ Don't know |  |  |
| Desirable Effects  How substantial are the desirable anticipated effects? | | |
| Judgement | Research evidence | Additional considerations |
| ○ Trivial ○ Small ● Moderate ○ Large ○ Varies ○ Don't know | **Outcomes** \| **Anticipated absolute effects^*^ (95% CI)** \| \| **Relative effect (95% CI)** \| **№ of participants (studies)** \| **Certainty of the evidence (GRADE)** \| **Comments**; \| **Risk with genetic testing alone** \| **Risk with imaging and genetic testing** \| \| \| \|; Diagnosis \| Study population \| \| **OR 0.93** (0.77 to 1.12) \| 2252 (39 observational studies) \| ⨁◯◯◯ Very low^a,b^ \|; \| 264 per 1,000 \| **250 per 1,000** (216 to 286) \| \| \| \|; Uptake of Testing \| Study population \| \| **OR 1.22** (1.00 to 1.50) \| 2223 (38 observational studies) \| ⨁◯◯◯ Very low^a,b^ \|; \| 764 per 1,000 \| **798 per 1,000** (764 to 829) \| \| \| \|; Quality of life \| Thijssen 2020 performed a health-related quality of life (HRQOL) in patients with thoracic aortic disease (TAD), those undergoing screening, and partners. The study found that patients with TAD had lower HRQOL, however, patients being screened were not as affected. Young patients and female patients score lower on the SF-36 and HADS compared to older male patients. Previous aortic surgery was associated with high HADS depression scores. \| \| - \| (1 observational study) \| ⨁◯◯◯ Very low \|; Psychometric testing \| Thijssen 2020 compared patients with TAD to general population and those screened. The authors found that compared with the general population patients with TAD had lower HRQOL, however, those screened were not as affected. Younger and female patients had significantly lower scores on the SF-36 and HADS compared to older and male participants. Smaller Aortic diameter was associated with better RDSQ, and previous aortic surgery was associated with higher depression scores. Factors related to psychological stress included coping skills, impact on social and professional life, state of aortic diameter as well as symptoms and disease related knowledge. \| \| - \| (1 observational study) \| ⨁◯◯◯ Very low \| Rated down for inconsistency across studies and statistical heterogeneity. rated down for low number of events. | Challenges with the data- don’t have enough data on follow up on patients passed screening in terms of long-term outcomes.      Patient partners- with imaging and genetics if one is not positive, the secondary test provides reassurance and improves quality of life. Also having options in screening. Screening identifies disease risk. Given the impact on the screening, from patient perspective desirable effects are significant.    From clinical perspective-more modalities to screening |
| Undesirable Effects  How substantial are the undesirable anticipated effects? | | |
| Judgement | Research evidence | Additional considerations |
| ○ Large ○ Moderate ● Small ○ Trivial ○ Varies ○ Don't know | **Outcomes** \| **Anticipated absolute effects^*^ (95% CI)** \| \| **Relative effect (95% CI)** \| **№ of participants (studies)** \| **Certainty of the evidence (GRADE)** \| **Comments**; \| **Risk with genetic testing alone** \| **Risk with imaging and genetic testing** \| \| \| \|; Diagnosis \| Study population \| \| **OR 0.93** (0.77 to 1.12) \| 2252 (39 observational studies) \| ⨁◯◯◯ Very low^a,b^ \|; \| 264 per 1,000 \| **250 per 1,000** (216 to 286) \| \| \| \|; Uptake of Testing \| Study population \| \| **OR 1.22** (1.00 to 1.50) \| 2223 (38 observational studies) \| ⨁◯◯◯ Very low^a,b^ \|; \| 764 per 1,000 \| **798 per 1,000** (764 to 829) \| \| \| \|; Quality of life \| Thijssen 2020 performed a health realted quality of life (HRQOL) in patients with thoracic aortic disease (TAD), those undergoing screening, and partners. The study found that patients with TAD had lower HRQOL, however, patients being screened were not as affected. Young patients and female patients score lower on the SF-36 and HADS compared to older male patients. Previous aortic surgery was associated with high HADS depression scores. \| \| - \| (1 observational study) \| ⨁◯◯◯ Very low \|; Psychometric testing \| Thijssen 2020 compared patients with TAD to general population and those screened. The authors found that compared with the general population patients with TAD had lower HRQOL, however, those screened were not as affected. Younger and female patients had significantly lower scores on the SF-36 and HADS compared to older and male participants. Smaller Aortic diameter was associated with better RDSQ, and previous aortic surgery was associated with higher depression scores. Factors related to psychological stress included coping skills, impact on social and professional life, state of aortic diameter as well as symptoms and disease related knowledge. \| \| - \| (1 observational study) \| ⨁◯◯◯ Very low \| Rated down for inconsistency across studies and statistical heterogeneity. rated down for low number of events | Residual concern remains if screening is negative. But being screening and monitor does provide reassurance.    Finding a variant of unclear significance, or a negative screening. Uncertainty may have increased stress for patients and impact of QoL. |
| Certainty of evidence  What is the overall certainty of the evidence of effects? | | |
| Judgement | Research evidence | Additional considerations |
| ● Very low ○ Low ○ Moderate ○ High ○ No included studies |  |  |
| Values  Is there important uncertainty about or variability in how much people value the main outcomes? | | |
| Judgement | Research evidence | Additional considerations |
| ○ Important uncertainty or variability ○ Possibly important uncertainty or variability ○ Probably no important uncertainty or variability ● No important uncertainty or variability |  |  |
| Balance of effects  Does the balance between desirable and undesirable effects favor the intervention or the comparison? | | |
| Judgement | Research evidence | Additional considerations |
| ○ Favors the comparison ○ Probably favors the comparison ○ Does not favor either the intervention or the comparison ● Probably favors the intervention ○ Favors the intervention ○ Varies ○ Don't know |  |  |
| Resources required  How large are the resource requirements (costs)? | | |
| Judgement | Research evidence | Additional considerations |
| ○ Large costs ○ Moderate costs ○ Negligible costs and savings ○ Moderate savings ○ Large savings ○ Varies ● Don't know | In Miller 2021 the cost range was $266 to $605 per participant (confirmation of genetic test from biobank result, plus offer of genetic counselling). |  |
| Certainty of evidence of required resources  What is the certainty of the evidence of resource requirements (costs)? | | |
| Judgement | Research evidence | Additional considerations |
| ○ Very low ○ Low ○ Moderate ○ High ● No included studies |  |  |
| Cost effectiveness  Does the cost-effectiveness of the intervention favor the intervention or the comparison? | | |
| Judgement | Research evidence | Additional considerations |
| ○ Favors the comparison ○ Probably favors the comparison ○ Does not favor either the intervention or the comparison ○ Probably favors the intervention ○ Favors the intervention ○ Varies ● No included studies |  |  |
| Equity  What would be the impact on health equity? | | |
| Judgement | Research evidence | Additional considerations |
| ○ Reduced ○ Probably reduced ○ Probably no impact ● Probably increased ○ Increased ○ Varies ○ Don't know |  |  |
| Acceptability  Is the intervention acceptable to key stakeholders? | | |
| Judgement | Research evidence | Additional considerations |
| ○ No ○ Probably no ○ Probably yes ● Yes ○ Varies ○ Don't know |  |  |
| Feasibility  Is the intervention feasible to implement? | | |
| Judgement | Research evidence | Additional considerations |
| ○ No ○ Probably no ● Probably yes ○ Yes ○ Varies ○ Don't know |  |  |

Summary of judgements

|  | **Judgement** | | | | | | |
| --- | --- | --- | --- | --- | --- | --- | --- |
| **Problem** | No | Probably no | **Probably yes** | Yes |  | Varies | Don't know |
| **Desirable Effects** | Trivial | Small | **Moderate** | Large |  | Varies | Don't know |
| **Undesirable Effects** | Large | Moderate | **Small** | Trivial |  | Varies | Don't know |
| **Certainty of evidence** | **Very low** | Low | Moderate | High |  |  | No included studies |
| **Values** | Important uncertainty or variability | Possibly important uncertainty or variability | Probably no important uncertainty or variability | **No important uncertainty or variability** |  |  |  |
| **Balance of effects** | Favors the comparison | Probably favors the comparison | Does not favor either the intervention or the comparison | **Probably favors the intervention** | Favors the intervention | Varies | Don't know |
| **Resources required** | Large costs | Moderate costs | Negligible costs and savings | Moderate savings | Large savings | Varies | **Don't know** |
| **Certainty of evidence of required resources** | Very low | Low | Moderate | High |  |  | **No included studies** |
| **Cost effectiveness** | Favors the comparison | Probably favors the comparison | Does not favor either the intervention or the comparison | Probably favors the intervention | Favors the intervention | Varies | **No included studies** |
| **Equity** | Reduced | Probably reduced | Probably no impact | **Probably increased** | Increased | Varies | Don't know |
| **Acceptability** | No | Probably no | Probably yes | **Yes** |  | Varies | Don't know |
| **Feasibility** | No | Probably no | **Probably yes** | Yes |  | Varies | Don't know |

Type of recommendation

| Strong recommendation against the intervention | Conditional recommendation against the intervention | Conditional recommendation for either the intervention or the comparison | **Conditional recommendation for the intervention** | Strong recommendation for the intervention |
| --- | --- | --- | --- | --- |
| ○ | ○ | ○ | **●** | ○ |

Conclusions

Recommendation

We suggest the use of imaging and genetic screening for aortopathy (Considerational Recommendation, Very Low Certainty of Evidence).

### eTable 6 – Evidence-to-decision framework for the PICO “Should gene panels (any) vs. whole exome sequencing be used for screening relatives of patients?”

The table below reports the GUIDE evidence-to-decision process to discuss the PICO related to the modality of genetic testing.

Question

**Should gene panels (any) vs. whole exome sequencing be used for screening relatives of patients affected by thoracic aortic diseases?**

Assessment

| Problem  Is the problem a priority? | | |
| --- | --- | --- |
| Judgement | Research evidence | Additional considerations |
| ○ No ○ Probably no ○ Probably yes ● Yes ○ Varies ○ Don't know |  | In addition to lifestyle risk factors, non-syndromic thoracic aortic diseases are also linked to family history. Screening relatives of probands/patients may detect early disease, and allow follow-up and intervention to reduce morbidity and mortality; on the other hand, screening relatives may cause distress and impact mental health.    These outcomes may be impacted by genetic testing strategies with different diagnosis rates (gene panel vs. whole exome sequencing). |
| Desirable Effects  How substantial are the desirable anticipated effects? | | |
| Judgement | Research evidence | Additional considerations |
| ○ Trivial ○ Small ○ Moderate ● Large ○ Varies ○ Don't know | Using a gene panel, the diagnosis rate is 21% (95% CI 0.16 to 0.26). Using whole exome sequencing, the diagnosis rate is 30% (95%CI 0.23 to 0.39); the difference in the diagnosis rate using whole exome sequencing is 10.8% (95%CI, 4.6% to 16%).    In a small study of 20 patients who had been identified in a biobank of having aortopathy genes, 10 accepted clinical genetic screening. Psychological distress (16.0 ± 4.2, out of 100) and decisional regret (11.5 ± 11.6, out of 100) scores were low, but only measured in the patients who completed the screening. | Higher diagnosis rate may be highly desired, whole exome sequencing results in a sizable absolute risk difference in diagnosis rate. At present, the disease is more likely underdiagnosed rather than overdiagnosed.    Whole exome sequencing can also facilitate research, or diagnosis at a later time. |
| Undesirable Effects  How substantial are the undesirable anticipated effects? | | |
| Judgement | Research evidence | Additional considerations |
| ○ Large ○ Moderate ● Small ○ Trivial ○ Varies ○ Don't know |  | Risks of false reassurance from negative tests may be higher with whole exome sequencing (perceived as a more definitive test).    Risk of overdiagnosis is the biggest drawback, overmedicalization, but these are unclear at present, and ongoing screening and follow-up may mitigate these.    Current risk is underdiagnosis of a serious and catastrophic aortic dissection. |
| Certainty of evidence  What is the overall certainty of the evidence of effects? | | |
| Judgement | Research evidence | Additional considerations |
| ○ Very low ● Low ○ Moderate ○ High ○ No included studies |  | Areas of uncertainty include diagnostic test characteristics (eg. false positives and false negative test rates), and long-term follow-up of screened patients.    What is included in the panels varies: some panels may have better or worse diagnosis rates. |
| Values  Is there important uncertainty about or variability in how much people value the main outcomes? | | |
| Judgement | Research evidence | Additional considerations |
| ○ Important uncertainty or variability ● Possibly important uncertainty or variability ○ Probably no important uncertainty or variability ○ No important uncertainty or variability |  | Some individuals may prefer early diagnosis, ongoing follow-up and being reassured by this, whereas others may prefer not to know. Higher diagnosis rates with whole exome sequences may weigh in.    Vast majority of patients will not be able to tell the difference here between gene panel and whole exome sequencing. |
| Balance of effects  Does the balance between desirable and undesirable effects favor the intervention or the comparison? | | |
| Judgement | Research evidence | Additional considerations |
| ● Favors the comparison ○ Probably favors the comparison ○ Does not favor either the intervention or the comparison ○ Probably favors the intervention ○ Favors the intervention ○ Varies ○ Don't know |  | Detection rate favours whole exome sequencing. Certainly not worse than gene panel. |
| Resources required  How large are the resource requirements (costs)? | | |
| Judgement | Research evidence | Additional considerations |
| ○ Large costs ● Moderate costs ○ Negligible costs and savings ○ Moderate savings ○ Large savings ○ Varies ○ Don't know |  | Gene panels are likely less resource intensive, but whole exome sequencing is becoming cheaper.    There is a significant increase in the amount of time required to review whole exome sequencing, and these are related to human resources more than finances. Gene panels also have some delays, but less than whole exome sequencing.    In the long term these may change, especially with growth in industry, and advocacy. |
| Certainty of evidence of required resources  What is the certainty of the evidence of resource requirements (costs)? | | |
| Judgement | Research evidence | Additional considerations |
| ○ Very low ● Low ○ Moderate ○ High ○ No included studies |  |  |
| Cost effectiveness  Does the cost-effectiveness of the intervention favor the intervention or the comparison? | | |
| Judgement | Research evidence | Additional considerations |
| ○ Favors the comparison ○ Probably favors the comparison ○ Does not favor either the intervention or the comparison ○ Probably favors the intervention ○ Favors the intervention ○ Varies ● No included studies |  |  |
| Equity  What would be the impact on health equity? | | |
| Judgement | Research evidence | Additional considerations |
| ○ Reduced ○ Probably reduced ○ Probably no impact ○ Probably increased ○ Increased ● Varies ○ Don't know |  | Role of private whole exome sequencing vs. public gene panel is an issue. This includes access to genetic counselling to provide context and understanding of results, even if they are faster/more accessible.    The time it takes to get results may result in decreased equity unless WES becomes more available and faster through the NHS.    Promoting WES may improve equity across the system as well.    Higher diagnostic rate will result in more equity by facilitating follow-up for other individuals who would test negative with gene panels.    Inequity of studies of genes and so gene panels may or may not apply across diverse groups within a multicultural setting. There was concern about uncertainty as whole exome sequencing may result in more diagnoses but unclear. |
| Acceptability  Is the intervention acceptable to key stakeholders? | | |
| Judgement | Research evidence | Additional considerations |
| ○ No ○ Probably no ○ Probably yes ● Yes ○ Varies ○ Don't know | Gene panels and whole exome sequencing have similar levels of acceptability, with approximately 75% of potential participants agreeing to testing to either form. |  |
| Feasibility  Is the intervention feasible to implement? | | |
| Judgement | Research evidence | Additional considerations |
| ○ No ○ Probably no ○ Probably yes ● Yes ○ Varies ○ Don't know |  | Refer to issues of resources above are the main barriers to feasibility.  Technology is continuing to advance; WES may become even more feasible with time, but these are theoretical. |

Summary of judgements

|  | **Judgement** | | | | | | |
| --- | --- | --- | --- | --- | --- | --- | --- |
| **Problem** | No | Probably no | Probably yes | **Yes** |  | Varies | Don't know |
| **Desirable Effects** | Trivial | Small | Moderate | **Large** |  | Varies | Don't know |
| **Undesirable Effects** | Large | Moderate | **Small** | Trivial |  | Varies | Don't know |
| **Certainty of evidence** | Very low | **Low** | Moderate | High |  |  | No included studies |
| **Values** | Important uncertainty or variability | **Possibly important uncertainty or variability** | Probably no important uncertainty or variability | No important uncertainty or variability |  |  |  |
| **Balance of effects** | **Favors the comparison** | Probably favors the comparison | Does not favor either the intervention or the comparison | Probably favors the intervention | Favors the intervention | Varies | Don't know |
| **Resources required** | Large costs | **Moderate costs** | Negligible costs and savings | Moderate savings | Large savings | Varies | Don't know |
| **Certainty of evidence of required resources** | Very low | **Low** | Moderate | High |  |  | No included studies |
| **Cost effectiveness** | Favors the comparison | Probably favors the comparison | Does not favor either the intervention or the comparison | Probably favors the intervention | Favors the intervention | Varies | **No included studies** |
| **Equity** | Reduced | Probably reduced | Probably no impact | Probably increased | Increased | **Varies** | Don't know |
| **Acceptability** | No | Probably no | Probably yes | **Yes** |  | Varies | Don't know |
| **Feasibility** | No | Probably no | Probably yes | **Yes** |  | Varies | Don't know |

Type of recommendation

| Strong recommendation against the intervention | Conditional recommendation against the intervention | Conditional recommendation for either the intervention or the comparison | **Conditional recommendation for the intervention** | Strong recommendation for the intervention |
| --- | --- | --- | --- | --- |
| ○ | ○ | ○ | **●** | ○ |

Conclusions

Recommendation

We suggest whole exome sequencing over gene panels in screening family members of individuals with non-syndrome thoracic aortic disease. (conditional recommendation, low certainty)

### eTable 7 – Evidence-to-decision framework for the PICO “Should imaging vs. usual care be used for screening first degree relatives of patients with non-syndromic aortic disease?”

The table below reports the GUIDE evidence-to-decision process to discuss the PICO related to the use of imaging test for screening in relatives of patients with non-syndromic aortic disease.

| Question | |
| --- | --- |
| **Should imaging vs. usual care be used for screening first degree relatives of patients with non-syndromic thoracic aortic disease?** | |
| **Population:** | screening first degree relatives of patients with non-syndromic thoracic aortic disease |
| **Intervention:** | imaging |
| **Comparison:** | usual care |
| **Main outcomes:** | Diagnosis rate; Patient depression symptoms ; Patient anxiety symptoms ; |

Assessment

| Problem  Is the problem a priority? | | |
| --- | --- | --- |
| Judgement | Research evidence | Additional considerations |
| ○ No ○ Probably no ○ Probably yes ● Yes ○ Varies ○ Don't know |  | Non-syndromic thoracic aortic diseases have a high mortality when they present as emergencies; early detection can facilitate earlier intervention to reduce morbidity and mortality. Evidence suggests that relatives of patients who present with non-syndromic thoracic aortic disease are at increased risk, raising the question of whether screening with imaging is warranted. |
| Desirable Effects  How substantial are the desirable anticipated effects? | | |
| Judgement | Research evidence | Additional considerations |
| ○ Trivial ○ Small ● Moderate ○ Large ○ Varies ○ Don't know | Imaging of first degree relative may result in a diagnosis rate of ~26% (95%CI 0.16, to 0.4). Screening of second degree relatives may result in a similar diagnosis rate (26%, 95% CI 0.18 to 0.37).    By comparison, incidental rates of diagnosis of aortic aneurysm on CT scans at a single hospital (Mori 2019) were 2.1%, with 97.8% occurring in patients age >50; rates increased with age, with 5.7% of males over 84 having aortic root dilation of >=4.5 cm. | No direct evidence of whether or not screening of relatives results in earlier intervention and improved clinical outcomes. Studies used different cutoffs to identify affected individuals (e.g. z-scores vs. absolute size). |
| Undesirable Effects  How substantial are the undesirable anticipated effects? | | |
| Judgement | Research evidence | Additional considerations |
| ○ Large ○ Moderate ○ Small ○ Trivial ● Varies ○ Don't know | There may be little to no difference in patient symptoms of depression or anxiety after screening. | The impact of screening may vary between individuals, the outcomes of studying, and resources and support available after screening. However, the panel judges this would likely be a minority of people. |
| Certainty of evidence  What is the overall certainty of the evidence of effects? | | |
| Judgement | Research evidence | Additional considerations |
| ○ Very low ● Low ○ Moderate ○ High ○ No included studies |  | Studies are consistent, but observational in nature with some limitations. |
| Values  Is there important uncertainty about or variability in how much people value the main outcomes? | | |
| Judgement | Research evidence | Additional considerations |
| ● Important uncertainty or variability ○ Possibly important uncertainty or variability ○ Probably no important uncertainty or variability ○ No important uncertainty or variability |  | Patient values and preferences for screening may vary and influence their decisions around whether or not they would want to be screened, for both first and second degree relatives. |
| Balance of effects  Does the balance between desirable and undesirable effects favor the intervention or the comparison? | | |
| Judgement | Research evidence | Additional considerations |
| ○ Favors the comparison ○ Probably favors the comparison ○ Does not favor either the intervention or the comparison ● Probably favors the intervention ○ Favors the intervention ○ Varies ○ Don't know |  | Most patients at risk of aortic disease would want to know to receive follow-up and intervention. |
| Resources required  How large are the resource requirements (costs)? | | |
| Judgement | Research evidence | Additional considerations |
| ● Large costs ○ Moderate costs ○ Negligible costs and savings ○ Moderate savings ○ Large savings ○ Varies ○ Don't know |  | Little direct evidence of costs, but screening requires more resources than not screening, including the initial tests and follow-ups for relatives of probands.    In Abbasciano et al (2022), 13 relatives required a follow-up scan at 5 years, and genetic test re-evaluation in 3 families. |
| Certainty of evidence of required resources  What is the certainty of the evidence of resource requirements (costs)? | | |
| Judgement | Research evidence | Additional considerations |
| ○ Very low ○ Low ○ Moderate ○ High ● No included studies |  |  |
| Cost effectiveness  Does the cost-effectiveness of the intervention favor the intervention or the comparison? | | |
| Judgement | Research evidence | Additional considerations |
| ○ Favors the comparison ○ Probably favors the comparison ○ Does not favor either the intervention or the comparison ○ Probably favors the intervention ○ Favors the intervention ○ Varies ● No included studies |  | This is uncertain; it could be cost-effective or not, but need more studies to determine if higher diagnosis rates result in better outcoems in a cost-effective manner. |
| Equity  What would be the impact on health equity? | | |
| Judgement | Research evidence | Additional considerations |
| ○ Reduced ○ Probably reduced ○ Probably no impact ● Probably increased ○ Increased ○ Varies ○ Don't know |  | Improved discussions with patients, increased information about their personal health, risks, and potential treatments. |
| Acceptability  Is the intervention acceptable to key stakeholders? | | |
| Judgement | Research evidence | Additional considerations |
| ○ No ○ Probably no ● Probably yes ○ Yes ○ Varies ○ Don't know |  | Probably acceptable to many relatives. Those who are not interested in screening may require more information to make the decision; for those who choose not to be screened (e.g. they have anxiety about finding out) it is likely easy to opt-out. |
| Feasibility  Is the intervention feasible to implement? | | |
| Judgement | Research evidence | Additional considerations |
| ○ No ○ Probably no ● Probably yes ○ Yes ○ Varies ○ Don't know |  | Screening first and second-degree relatives with imaging outside of a research setting may be challenging or cost-prohibitive. Wait times for imaging are already long and screening more individuals may make this worse or even be prohibitive in some regions. Access to follow-up, psychological support, etc. with respect to screening may or may not be available. |

Summary of judgements

|  | **Judgement** | | | | | | |
| --- | --- | --- | --- | --- | --- | --- | --- |
| **Problem** | No | Probably no | Probably yes | **Yes** |  | Varies | Don't know |
| **Desirable Effects** | Trivial | Small | **Moderate** | Large |  | Varies | Don't know |
| **Undesirable Effects** | Large | Moderate | Small | Trivial |  | **Varies** | Don't know |
| **Certainty of evidence** | Very low | **Low** | Moderate | High |  |  | No included studies |
| **Values** | **Important uncertainty or variability** | Possibly important uncertainty or variability | Probably no important uncertainty or variability | No important uncertainty or variability |  |  |  |
| **Balance of effects** | Favors the comparison | Probably favors the comparison | Does not favor either the intervention or the comparison | **Probably favors the intervention** | Favors the intervention | Varies | Don't know |
| **Resources required** | **Large costs** | Moderate costs | Negligible costs and savings | Moderate savings | Large savings | Varies | Don't know |
| **Certainty of evidence of required resources** | Very low | Low | Moderate | High |  |  | **No included studies** |
| **Cost effectiveness** | Favors the comparison | Probably favors the comparison | Does not favor either the intervention or the comparison | Probably favors the intervention | Favors the intervention | Varies | **No included studies** |
| **Equity** | Reduced | Probably reduced | Probably no impact | **Probably increased** | Increased | Varies | Don't know |
| **Acceptability** | No | Probably no | **Probably yes** | Yes |  | Varies | Don't know |
| **Feasibility** | No | Probably no | **Probably yes** | Yes |  | Varies | Don't know |

Type of recommendation

| Strong recommendation against the intervention | Conditional recommendation against the intervention | Conditional recommendation for either the intervention or the comparison | Conditional recommendation for the intervention | **Strong recommendation for the intervention** |
| --- | --- | --- | --- | --- |
| ○ | ○ | ○ | ○ | **●** |

Conclusions

Recommendation

We recommend extensive adoption of imaging screening in first degree relatives of patients with non-syndromic thoracic aortic disease. (strong recommendation, low certainty of evidence)

Justification

Large benefit of screening (high diagnosis rate) in first degree relatives provides an opportunity to avoid a catastrophic event.

### eTable 8 – Evidence-to-decision framework for the PICO “Should decision support tools vs. usual care be used for survivors of acute aortic syndromes and their families?”

The table below reports the GUIDE evidence-to-decision process to discuss the PICO related to the use of decision support tool to provide care to survivors of acute aortic syndromes and their relatives.

| Question | |
| --- | --- |
| **Should decision support tools vs. usual care be used for survivors of acute aortic syndromes and their families?** | |
| **Population:** | survivors of acute aortic syndromes and their families |
| **Intervention:** | decision support tools |
| **Comparison:** | usual care |
| **Main outcomes:** | Patient satisfaction; Patient understanding; Decisional Conflict; Patient anxiety; Quality of Life; |
| **Setting:** |  |
| **Perspective:** |  |
| **Background:** |  |
| **Conflict of interests:** |  |

Assessment

| Problem  Is the problem a priority? | | |
| --- | --- | --- |
| Judgement | Research evidence | Additional considerations |
| ○ No ○ Probably no ○ Probably yes ● Yes ○ Varies ○ Don't know |  | Patients with thoracic aortic diseases have to make value-laden decisions around diagnosis, monitoring, and management. Decision support tools aim to assist patients in making choices which reflect their values and improve the process of care. |
| Desirable Effects  How substantial are the desirable anticipated effects? | | |
| Judgement | Research evidence | Additional considerations |
| ○ Trivial ○ Small ● Moderate ○ Large ○ Varies ○ Don't know | Possibly small improvement in patient knowledge and understanding.  Possibly little to no difference in patient satisfaction, decisional conflict, anxiety, or quality of life. | There is evidence in other fields supporting the use of decision support tools in other conditions. Whether these would apply to thoracic aortic diseases and the associated decisions is less clear.  How informative and accessible the tools are will impact the magnitude of effects.    There may be other clinical benefits in terms of efficiency, allowing richer and more focused discussions around key clinical issues. |
| Undesirable Effects  How substantial are the undesirable anticipated effects? | | |
| Judgement | Research evidence | Additional considerations |
| ○ Large ○ Moderate ○ Small ● Trivial ○ Varies ○ Don't know | In one study (Leblanc 2018), there was a non-significant trend toward a lengthier discussion for patients receiving the intervention (8.18 ± 4.28 minutes vs 6.35 ± 2.72 minutes; P = 0.07). This may be a positive outcome from the perspective of the patient, having more time to review and discuss. |  |
| Certainty of evidence  What is the overall certainty of the evidence of effects? | | |
| Judgement | Research evidence | Additional considerations |
| ○ Very low ● Low ○ Moderate ○ High ○ No included studies | Certainty of evidence is low for all outcomes, rated down for indirectness and imprecision. |  |
| Values  Is there important uncertainty about or variability in how much people value the main outcomes? | | |
| Judgement | Research evidence | Additional considerations |
| ○ Important uncertainty or variability ○ Possibly important uncertainty or variability ● Probably no important uncertainty or variability ○ No important uncertainty or variability |  | Assuming decision tools are helpful most patients would agree to their use. |
| Balance of effects  Does the balance between desirable and undesirable effects favor the intervention or the comparison? | | |
| Judgement | Research evidence | Additional considerations |
| ○ Favors the comparison ○ Probably favors the comparison ○ Does not favor either the intervention or the comparison ● Probably favors the intervention ○ Favors the intervention ○ Varies ○ Don't know |  | Main limitation here is the quality/certainty of evidence. |
| Resources required  How large are the resource requirements (costs)? | | |
| Judgement | Research evidence | Additional considerations |
| ○ Large costs ● Moderate costs ○ Negligible costs and savings ○ Moderate savings ○ Large savings ○ Varies ○ Don't know |  | Moderate costs to design, implement, and updated the decision support tools will require more resources compared to simply providing usual care. |
| Certainty of evidence of required resources  What is the certainty of the evidence of resource requirements (costs)? | | |
| Judgement | Research evidence | Additional considerations |
| ○ Very low ○ Low ○ Moderate ○ High ● No included studies |  |  |
| Cost effectiveness  Does the cost-effectiveness of the intervention favor the intervention or the comparison? | | |
| Judgement | Research evidence | Additional considerations |
| ○ Favors the comparison ○ Probably favors the comparison ○ Does not favor either the intervention or the comparison ○ Probably favors the intervention ○ Favors the intervention ○ Varies ● No included studies |  | Cost-effectiveness may vary; the downstream effects and whether or not costs are increased or decreased are difficult to predict.    The opportunity costs and time required may be increased or decreased, depending on the decision support tool. |
| Equity  What would be the impact on health equity? | | |
| Judgement | Research evidence | Additional considerations |
| ○ Reduced ○ Probably reduced ○ Probably no impact ○ Probably increased ○ Increased ● Varies ○ Don't know |  | It may increase equity by providing more standardized information and decision-making processes for patients, including those who may not have access to such information. This depends on whether or not they the tools are provided equitably in a consistent way, and whether the DST supplement current standard processes or replace them.    There is an opportunity to provide DST in multiple languages which are suited to the needs of individual populations.    On the other hand, digital support tools may not be as accessible to some individuals who are less 'digitally literate.'    Equity of access to services after the tool is used may vary as well. |
| Acceptability  Is the intervention acceptable to key stakeholders? | | |
| Judgement | Research evidence | Additional considerations |
| ○ No ○ Probably no ● Probably yes ○ Yes ○ Varies ○ Don't know |  | Many patients with thoracic aortic diseases would likely accept the use of DST, unless these were going to replace current processes (e.g. DST instead of meeting with the clinical team). If they supplement and reinforce current processes, they will likely be acceptable. |
| Feasibility  Is the intervention feasible to implement? | | |
| Judgement | Research evidence | Additional considerations |
| ○ No ○ Probably no ● Probably yes ○ Yes ○ Varies ○ Don't know |  | DST are used in other fields to assist patients and families in other decisions, both digital and paper versions, demonstrating the feasibility of these overall.    Technological developments eg. AI, ChatGPT etc. may make the implementation of DSTs more feasible. |

Summary of judgements

|  | **Judgement** | | | | | | |
| --- | --- | --- | --- | --- | --- | --- | --- |
| **Problem** | No | Probably no | Probably yes | **Yes** |  | Varies | Don't know |
| **Desirable Effects** | Trivial | Small | **Moderate** | Large |  | Varies | Don't know |
| **Undesirable Effects** | Large | Moderate | Small | **Trivial** |  | Varies | Don't know |
| **Certainty of evidence** | Very low | **Low** | Moderate | High |  |  | No included studies |
| **Values** | Important uncertainty or variability | Possibly important uncertainty or variability | **Probably no important uncertainty or variability** | No important uncertainty or variability |  |  |  |
| **Balance of effects** | Favors the comparison | Probably favors the comparison | Does not favor either the intervention or the comparison | **Probably favors the intervention** | Favors the intervention | Varies | Don't know |
| **Resources required** | Large costs | **Moderate costs** | Negligible costs and savings | Moderate savings | Large savings | Varies | Don't know |
| **Certainty of evidence of required resources** | Very low | Low | Moderate | High |  |  | **No included studies** |
| **Cost effectiveness** | Favors the comparison | Probably favors the comparison | Does not favor either the intervention or the comparison | Probably favors the intervention | Favors the intervention | Varies | **No included studies** |
| **Equity** | Reduced | Probably reduced | Probably no impact | Probably increased | Increased | **Varies** | Don't know |
| **Acceptability** | No | Probably no | **Probably yes** | Yes |  | Varies | Don't know |
| **Feasibility** | No | Probably no | **Probably yes** | Yes |  | Varies | Don't know |

Type of recommendation

| Strong recommendation against the intervention | Conditional recommendation against the intervention | Conditional recommendation for either the intervention or the comparison | **Conditional recommendation for the intervention** | Strong recommendation for the intervention |
| --- | --- | --- | --- | --- |
| ○ | ○ | ○ | **●** | ○ |

Conclusions

Recommendation

We suggest using DSTs for survivors of acute aortic syndromes and their families to help guide screening and treatment decisions. (conditional recommendation, low certainty evidence)

### eTable 9 – Evidence-to-decision framework for the PICO “*Should angiotensin receptor blockers (ARB) vs. no angiotensin receptor blockers be used for patients at risk of aortopathy?*”

The table below reports the GUIDE evidence-to-decision process to discuss the PICO related to the use of angiotensin receptor blockers in patients at risk of aortopathy.

| Question | |
| --- | --- |
| **Should angiotensin receptor blockers vs. no angiotensin receptor blockers be used for patients at risk of aortopathy?** | |
| **Population:** | patients at risk of aortopathy |
| **Intervention:** | angiotensin receptor blockers |
| **Comparison:** | no angiotensin receptor blockers |
| **Main outcomes:** | Mortality - ARB vs. control; Mortality - ARB vs b-blocker; Acute aortic syndromes - ARB vs. control; Acute aortic syndromes - ARB vs b-blocker; Need for surgery - ARB vs. control; Need for surgery - ARB vs b-blocker; Size change - ARB vs. control; Size change - ARB vs. b-blocker; |

Assessment

| Problem  Is the problem a priority? | | |
| --- | --- | --- |
| Judgement | Research evidence | Additional considerations |
| ○ No ○ Probably no ○ Probably yes ○ Yes ○ Varies ○ Don't know |  | Patients at risk of aortopathy are likely to value interventions that will reduce their risk of complications or surgery. |
| Desirable Effects  How substantial are the desirable anticipated effects? | | |
| Judgement | Research evidence | Additional considerations |
| ○ Trivial ● Small ○ Moderate ○ Large ○ Varies ○ Don't know | **Outcomes** \| **№ of participants (studies) Follow-up** \| **Certainty of the evidence (GRADE)** \| **Relative effect (95% CI)** \| **Anticipated absolute effects^*^ (95% CI)** \|; \| \| \| \| **Risk with no angiotensin receptor blockers** \| **Risk difference with angiotensin receptor blockers**; Mortality - ARB vs. control \| 452 (2 RCTs) \| ⨁⨁◯◯ Low^a^ \| **RR 0.11** (0.01 to 0.88) \| Study population \|; \| \| \| \| 35 per 1,000 \| **32 fewer per 1,000** (35 fewer to 4 fewer); Mortality - ARB vs b-blocker \| 748 (2 RCTs) \| ⨁⨁◯◯ Low^a^ \| **RR 2.33** (0.35 to 15.58) \| Study population \|; \| \| \| \| 3 per 1,000 \| **4 more per 1,000** (2 fewer to 39 more); Acute aortic syndromes - ARB vs. control \| 695 (5 RCTs) \| ⨁⨁◯◯ Low^a^ \| **RR 0.49** (0.21 to 1.14) \| Study population \|; \| \| \| \| 41 per 1,000 \| **21 fewer per 1,000** (33 fewer to 6 more); Acute aortic syndromes - ARB vs b-blocker \| 748 (2 RCTs) \| ⨁⨁◯◯ Low^a,b^ \| **RR 1.00** (0.23 to 4.35) \| Study population \|; \| \| \| \| 8 per 1,000 \| **0 fewer per 1,000** (6 fewer to 27 more); Need for surgery - ARB vs. control \| 695 (5 RCTs) \| ⨁⨁◯◯ Low^a^ \| **RR 0.97** (0.61 to 1.55) \| Study population \|; \| \| \| \| 92 per 1,000 \| **3 fewer per 1,000** (36 fewer to 51 more); Need for surgery - ARB vs b-blocker \| 748 (2 RCTs) \| ⨁⨁◯◯ Low^a^ \| **RR 1.36** (0.77 to 2.41) \| Study population \|; \| \| \| \| 51 per 1,000 \| **18 more per 1,000** (12 fewer to 72 more); Size change - ARB vs. control \| 626 (4 RCTs) \| ⨁⨁⨁◯ Moderate^c^ \| - \| The mean size change - ARB vs. control was **0.13** annual change in Z score \| MD **0.07 annual change in Z score lower** (0.11 lower to 0.01 lower); Size change - ARB vs. b-blocker \| 722 (3 RCTs) \| ⨁⨁◯◯ Low^d,e^ \| - \| The mean size change - ARB vs. b-blocker was **-0.11** annual change in Z score \| MD **0.02 annual change in Z score higher** (0.05 lower to 0.1 higher) Very small number of events leading to substantial uncertainty due to very serious imprecision. Although significant statistical heterogeneity (I2 = 50%) this is likely due to the very small number of events and resulting very serious imprecision in the included studies. We chose to rate down as even small differences in analysis method (eg. changing from fixed to random-effects analysis) changes whether the results are statistically significant or not. Serious statistical heterogeneity (I2 =63%). Wide 95% confidence intervals which do not exclude clinically relevant benefit or harm, resulting in uncertainty due to serious imprecision. | Notes:  75% of patients in each arm of the ARB vs. Control comparisons were on B-blockers.  "Acute aortic syndromes" includes any dissection or emergency surgery for dissection  "Need for surgery" includes emergency surgery of aorta or any vascular repair (aortic or other); time to event data was not available  Size change is body surface area-adjusted aortic root dimension Z score at the sinuses of Valsalva.  small for ARB vs. no ARB; trivial for ARB vs. b-blocker |
| Undesirable Effects  How substantial are the undesirable anticipated effects? | | |
| Judgement | Research evidence | Additional considerations |
| ○ Large ○ Moderate ● Small ○ Trivial ○ Varies ○ Don't know | *Forteza 2016:* discontinuation of medication 4/70 losartan, 6/70 atenolol; reduced dose for side effects losaratan 4/70; atenolol 6/70  *Groenink 2013:* 17/116 discontinued losartan due to 14 dizziness/low BP; 1 renal dysfunction; 1 fatigue; 1 angioedema  *Lacro 2014:* serious adverse events 50/305 losartan vs. 40/303 atenolol  *Milleron 2015:* adverse events possibly related to drug losartan 6/151 vs placebo 0/148; any adverse event losartan 51/151 and placebo 48/148  *Mullen 2019:* no difference in serious adverse events. 21/104 vs 24/88 | possibility of reducing dosing to mitigate these effects, but unclear if benefits are still present or attenuated at a lower dose.  higher risk of adverse events (mostly hypotension) in patients without underlying hypertension, autonomic dysfunction |
| Certainty of evidence  What is the overall certainty of the evidence of effects? | | |
| Judgement | Research evidence | Additional considerations |
| ○ Very low ● Low ○ Moderate ○ High ○ No included studies | Overall certainty of evidence for critical outcomes (eg. mortality, acute aortic syndromes) is low. |  |
| Values  Is there important uncertainty about or variability in how much people value the main outcomes? | | |
| Judgement | Research evidence | Additional considerations |
| ○ Important uncertainty or variability ● Possibly important uncertainty or variability ○ Probably no important uncertainty or variability ○ No important uncertainty or variability |  | Possibly variability in terms patient willingness to take medications for prevention of future events depending on an individual's understanding/appreciation of prognosis, engagement in their own health, experience with illness, and stage of their life (e.g. younger patients vs. older patients may value prevention, side effects differently). These differences may be similar, or greater with b-blockers. |
| Balance of effects  Does the balance between desirable and undesirable effects favor the intervention or the comparison? | | |
| Judgement | Research evidence | Additional considerations |
| ○ Favors the comparison ○ Probably favors the comparison ○ Does not favor either the intervention or the comparison ● Probably favors the intervention ○ Favors the intervention ○ Varies ○ Don't know | ARB vs. control: probably favours ARB, whether or not B-blocker is already used  ARB vs. B-blocker: probably not favouring one over the other |  |
| Resources required  How large are the resource requirements (costs)? | | |
| Judgement | Research evidence | Additional considerations |
| ○ Large costs ○ Moderate costs ● Negligible costs and savings ○ Moderate savings ○ Large savings ○ Varies ○ Don't know |  | ARB are more expensive than B-blockers; they require monitoring of renal function and thus overall slightly higher resource use than B-blocker but this is a comparatively small cost |
| Certainty of evidence of required resources  What is the certainty of the evidence of resource requirements (costs)? | | |
| Judgement | Research evidence | Additional considerations |
| ○ Very low ○ Low ○ Moderate ○ High ● No included studies |  |  |
| Cost effectiveness  Does the cost-effectiveness of the intervention favor the intervention or the comparison? | | |
| Judgement | Research evidence | Additional considerations |
| ○ Favors the comparison ○ Probably favors the comparison ○ Does not favor either the intervention or the comparison ○ Probably favors the intervention ○ Favors the intervention ○ Varies ● No included studies | Indirect evidence of cost-effectiveness of anti-hypertensives to treat hypertension suggest that ARBs may be more cost effective than B-blockers, and that either is more cost-effective than no treatment.    However in patients without pre-existing hypertension, these comparisons likely no longer apply. |  |
| Equity  What would be the impact on health equity? | | |
| Judgement | Research evidence | Additional considerations |
| ○ Reduced ○ Probably reduced ● Probably no impact ○ Probably increased ○ Increased ○ Varies ○ Don't know |  | No evidence of a differential effect upon equity; it is not an overly expensive or prohibitive treatment that would impact equity by any particular group. |
| Acceptability  Is the intervention acceptable to key stakeholders? | | |
| Judgement | Research evidence | Additional considerations |
| ○ No ○ Probably no ○ Probably yes ○ Yes ● Varies ○ Don't know |  | patients would have variability in acceptability of preventative treatment over the long term |
| Feasibility  Is the intervention feasible to implement? | | |
| Judgement | Research evidence | Additional considerations |
| ○ No ○ Probably no ○ Probably yes ● Yes ○ Varies ○ Don't know |  | Studies have demonstrated that the treatments are feasible, as has long-term use of these antihypertensive medications. |

Summary of judgements

|  | **Judgement** | | | | | | |
| --- | --- | --- | --- | --- | --- | --- | --- |
| **Problem** | No | Probably no | Probably yes | Yes |  | Varies | Don't know |
| **Desirable Effects** | Trivial | **Small** | Moderate | Large |  | Varies | Don't know |
| **Undesirable Effects** | Large | Moderate | **Small** | Trivial |  | Varies | Don't know |
| **Certainty of evidence** | Very low | **Low** | Moderate | High |  |  | No included studies |
| **Values** | Important uncertainty or variability | **Possibly important uncertainty or variability** | Probably no important uncertainty or variability | No important uncertainty or variability |  |  |  |
| **Balance of effects** | Favors the comparison | Probably favors the comparison | Does not favor either the intervention or the comparison | **Probably favors the intervention** | Favors the intervention | Varies | Don't know |
| **Resources required** | Large costs | Moderate costs | **Negligible costs and savings** | Moderate savings | Large savings | Varies | Don't know |
| **Certainty of evidence of required resources** | Very low | Low | Moderate | High |  |  | **No included studies** |
| **Cost effectiveness** | Favors the comparison | Probably favors the comparison | Does not favor either the intervention or the comparison | Probably favors the intervention | Favors the intervention | Varies | **No included studies** |
| **Equity** | Reduced | Probably reduced | **Probably no impact** | Probably increased | Increased | Varies | Don't know |
| **Acceptability** | No | Probably no | Probably yes | Yes |  | **Varies** | Don't know |
| **Feasibility** | No | Probably no | Probably yes | **Yes** |  | Varies | Don't know |

Type of recommendation

| Strong recommendation against the intervention | Conditional recommendation against the intervention | Conditional recommendation for either the intervention or the comparison | **Conditional recommendation for the intervention** | Strong recommendation for the intervention |
| --- | --- | --- | --- | --- |
| ○ | ○ | ○ | **●** | ○ |

Conclusions

Recommendation

We suggest using an ARB in patients with Marfan's syndrome, whether or not they are already on a B-blocker. (conditional recommendation, low certainty evidence)

Conditions for this recommendation include firstly, the evidence is only applicable to patients with Marfan's syndrome; this recommendation does not directly apply to patients without Marfan's (eg. patients with other syndromes or non-syndromic patients). The choice of which medication (if any) for these patients in the interim, can be guided by anticipated mechanism of effect (eg. genetics/pathology vs. shear stress etc.) until further evidence is available.

Justification

The evidence suggests that ARBs are similar in effects to B-blocker when compared directly and provide additional benefit when added to B-blockers. There are side-effects/adverse reactions to the medications, and patients may value differently the trade-offs between slowing progression of disease vs. these effects.

Research priorities

need for more evidence on anti-hypertensive therapy and pharmacotherapy in other syndromes and in non-syndromic patients.

need for more evidence of the impacts of these therapies on long-term and patient-important outcomes

### eTable 10 – Evidence-to-decision framework for the PICO “*Should beta-blockers vs. no beta-blockers be used for patients at risk of aortopathy?*”

The table below reports the GUIDE evidence-to-decision process to discuss the PICO related to the use of beta blockers in patients at risk of aortopathy.

| Question | |
| --- | --- |
| **Should beta-bockers vs. no beta-blockers be used for patients at risk of aortopathy?** | |
| **Population:** | patients at risk of aortopathy |
| **Intervention:** | beta-bockers |
| **Comparison:** | no beta-blockers |
| **Main outcomes:** | Mortality; Acute aortic syndromes; Deaths or CV events - Marfan's Observational; Disease progression; |

Assessment

| Problem  Is the problem a priority? | | |
| --- | --- | --- |
| Judgement | Research evidence | Additional considerations |
| ○ No ○ Probably no ○ Probably yes ● Yes ○ Varies ○ Don't know |  | Patients at risk of aortopathy would likely value interventions that will reduce their risk of complications or surgery. |
| Desirable Effects  How substantial are the desirable anticipated effects? | | |
| Judgement | Research evidence | Additional considerations |
| ○ Trivial ● Small ○ Moderate ○ Large ○ Varies ○ Don't know | **Outcomes** \| **Relative effect (95% CI)** \| **Anticipated absolute effects^*^ (95% CI)** \| \| \| **Certainty of the evidence (GRADE)** \| **What happens**; \| \| **Without beta-bockers** \| **With beta-bockers** \| **Difference** \| \|; Mortality № of participants: 123 (2 RCTs) \| **RR 0.32** (0.05 to 1.87) \| Study population \| \| \| ⨁⨁◯◯ Low^a^ \|; \| \| 7.6% \| **2.4%** (0.4 to 14.2) \| **5.2% fewer** (7.2 fewer to 6.6 more) \| \|; Acute aortic syndromes № of participants: 120 (2 RCTs) \| **RR 0.56** (0.15 to 2.13) \| Study population \| \| \| ⨁⨁◯◯ Low^a^ \|; \| \| 9.5% \| **5.3%** (1.4 to 20.3) \| **4.2% fewer** (8.1 fewer to 10.8 more) \| \|; Deaths or CV events - Marfan's Observational № of participants: 732 (5 observational studies) \| **RR 1.47** (1.11 to 1.96) \| Study population \| \| \| ⨁◯◯◯ Very low^b^ \|; \| \| 19.6% \| **28.9%** (21.8 to 38.5) \| **9.2% more** (2.2 more to 18.9 more) \| \|; Disease progression № of participants: 123 (2 RCTs) \| **RR 0.45** (0.20 to 1.02) \| Study population \| \| \| ⨁⨁◯◯ Low^a^ \|; \| \| 22.7% \| **10.2%** (4.5 to 23.2) \| **12.5% fewer** (18.2 fewer to 0.5 more) \| \| Small trials with a limited number of events leading to very serious uncertainty of effects due to imprecision. Statistically significant but a small number of events and optimal information size not met, resulting in serious imprecision. | Notes:  Shores 1994 included patients with Marfan's; Ong 2010 included patients with Ehlers-Danlos  Acute aortic syndromes includes either type A or B dissections, or emergency surgery for dissections.  "Disease progression" as a dichotomous outcome was defined in the studies, typically using a threshold cutoff.  Rate of change is clinically important in Marfan's as it is prognostically relevant, but we expect event rates are very low unless studies have a very long follow-up  Length of follow-up in studies is relatively short and may not result in adequate power for events. |
| Undesirable Effects  How substantial are the undesirable anticipated effects? | | |
| Judgement | Research evidence | Additional considerations |
| ○ Large ○ Moderate ● Small ○ Trivial ○ Varies ○ Don't know | *Ong 2010:* 3/25 patients reported fatigue.  *Shores 1994:* 10/30 patients with propranolol had adverse effects (heart block, lethargy, depression, insomnia, dream disturbance, bronchospasm, accentuated effects of EtOH) | B-blockers are not likely to have greater risk of adverse events than other populations on these medications  Modern cardio selective B-blockers are less likely to have effects than non-selective B-blockers. |
| Certainty of evidence  What is the overall certainty of the evidence of effects? | | |
| Judgement | Research evidence | Additional considerations |
| ○ Very low ● Low ○ Moderate ○ High ○ No included studies | Overall certainty of evidence for critical outcomes (eg. mortality, acute aortic syndromes) is low. |  |
| Values  Is there important uncertainty about or variability in how much people value the main outcomes? | | |
| Judgement | Research evidence | Additional considerations |
| ○ Important uncertainty or variability ● Possibly important uncertainty or variability ○ Probably no important uncertainty or variability ○ No important uncertainty or variability |  | Possibly variability in terms patient willingness to take medications for prevention of future events depending on an individual's understanding and appreciation of prognosis; engagement in their own health; experience with illness; and stage of their life (e.g. younger patients vs. older patients may value prevention vs. side effects differently) |
| Balance of effects  Does the balance between desirable and undesirable effects favor the intervention or the comparison? | | |
| Judgement | Research evidence | Additional considerations |
| ○ Favors the comparison ○ Probably favors the comparison ○ Does not favor either the intervention or the comparison ● Probably favors the intervention ○ Favors the intervention ○ Varies ○ Don't know |  | Potentially different effects between Marfans patients vs. Ehlers-Danlos patients as rate of change Marfan is more prognostically relevant in terms of dissection in Ehlers-Danlos, in which events may be more sporadic and less associated with size change progression. We are more certain that the balance of effects favours b-blockers in Marfans vs. other patients at risk of aortopathy. |
| Resources required  How large are the resource requirements (costs)? | | |
| Judgement | Research evidence | Additional considerations |
| ○ Large costs ● Moderate costs ○ Negligible costs and savings ○ Moderate savings ○ Large savings ○ Varies ○ Don't know |  | Consider indirect evidence: While the individual cost per tablet may not be high, costs of antihypertensives over a very long time are not negligible could be thousands of pounds over many years. These costs are potentially offset by reducing events, but we are uncertain overall of the costs vs. savings. |
| Certainty of evidence of required resources  What is the certainty of the evidence of resource requirements (costs)? | | |
| Judgement | Research evidence | Additional considerations |
| ○ Very low ○ Low ○ Moderate ○ High ● No included studies |  | No studies included but could extrapolate from other long-term antihypertensive prescriptions. |
| Cost effectiveness  Does the cost-effectiveness of the intervention favor the intervention or the comparison? | | |
| Judgement | Research evidence | Additional considerations |
| ○ Favors the comparison ○ Probably favors the comparison ○ Does not favor either the intervention or the comparison ○ Probably favors the intervention ○ Favors the intervention ○ Varies ● No included studies |  | No included studies. |
| Equity  What would be the impact on health equity? | | |
| Judgement | Research evidence | Additional considerations |
| ○ Reduced ○ Probably reduced ● Probably no impact ○ Probably increased ○ Increased ○ Varies ○ Don't know |  | No evidence of a differential effect upon equity; it is not an overly expensive or prohibitive treatment that would impact equity by any particular group. |
| Acceptability  Is the intervention acceptable to key stakeholders? | | |
| Judgement | Research evidence | Additional considerations |
| ○ No ○ Probably no ● Probably yes ○ Yes ○ Varies ○ Don't know |  | There is probably some variability in people's attitudes toward taking preventative medication for a prolonged period of time vs. preventing future illness and need for surgery. |
| Feasibility  Is the intervention feasible to implement? | | |
| Judgement | Research evidence | Additional considerations |
| ○ No ○ Probably no ● Probably yes ○ Yes ○ Varies ○ Don't know |  | The studies demonstrate the feasibility of providing b-blockers, as does indirect evidence from antihypertensive studies. |

Summary of judgements

|  | **Judgement** | | | | | | |
| --- | --- | --- | --- | --- | --- | --- | --- |
| **Problem** | No | Probably no | Probably yes | **Yes** |  | Varies | Don't know |
| **Desirable Effects** | Trivial | **Small** | Moderate | Large |  | Varies | Don't know |
| **Undesirable Effects** | Large | Moderate | **Small** | Trivial |  | Varies | Don't know |
| **Certainty of evidence** | Very low | **Low** | Moderate | High |  |  | No included studies |
| **Values** | Important uncertainty or variability | **Possibly important uncertainty or variability** | Probably no important uncertainty or variability | No important uncertainty or variability |  |  |  |
| **Balance of effects** | Favors the comparison | Probably favors the comparison | Does not favor either the intervention or the comparison | **Probably favors the intervention** | Favors the intervention | Varies | Don't know |
| **Resources required** | Large costs | **Moderate costs** | Negligible costs and savings | Moderate savings | Large savings | Varies | Don't know |
| **Certainty of evidence of required resources** | Very low | Low | Moderate | High |  |  | **No included studies** |
| **Cost effectiveness** | Favors the comparison | Probably favors the comparison | Does not favor either the intervention or the comparison | Probably favors the intervention | Favors the intervention | Varies | **No included studies** |
| **Equity** | Reduced | Probably reduced | **Probably no impact** | Probably increased | Increased | Varies | Don't know |
| **Acceptability** | No | Probably no | **Probably yes** | Yes |  | Varies | Don't know |
| **Feasibility** | No | Probably no | **Probably yes** | Yes |  | Varies | Don't know |

Type of recommendation

| Strong recommendation against the intervention | Conditional recommendation against the intervention | Conditional recommendation for either the intervention or the comparison | **Conditional recommendation for the intervention** | Strong recommendation for the intervention |
| --- | --- | --- | --- | --- |
| ○ | ○ | ○ | **●** | ○ |

Conclusions

Recommendation

We suggest using b-blockers in patients at risk for aortopathy, specifically those with Marfan's syndrome. (conditional recommendation, low certainty evidence)

Conditions to this recommendation include, certainty of evidence is low/very low, and the only clear benefit is reduction in growth rate. The evidence is less clear about long-term, patient-important benefits such as dissection, need for surgery. Secondly, the evidence for syndromic non-Marfan's (Ehlers-Danlos) and non-syndromic patients is limited, as the available evidence focused on Marfan's syndrome patients. Lastly, this recommendation needs to be considered within the context of other treatment options that are available (including ARBs).

Justification

Small effect on disease progression, but uncertain of its effect upon the most important clinical outcomes (surgery); downsides are also small; patients may have varied perspectives on the values and trade-offs.

### eTable 12 – Evidence-to-decision framework for the PICO “*Should Antiplatelet therapy vs. placebo be used for patients with aortopathy?*”

The table below reports the GUIDE evidence-to-decision process to discuss the PICO related to the use of antiplatelet agents in patients at risk of aortopathy.

| Question | |
| --- | --- |
| **Should Antiplatelet therapy vs. placebo be used for patients with aortopathy?** | |
| **Population:** | Aortic syndromes |
| **Intervention:** | Antiplatelet therapy |
| **Comparison:** | placebo |
| **Main outcomes:** | Disease Progression; |
| **Setting:** |  |
| **Perspective:** |  |
| **Background:** |  |
| **Conflict of interests:** |  |

Assessment

| Problem  Is the problem a priority? | | |
| --- | --- | --- |
| Judgement | Research evidence | Additional considerations |
| ○ No ○ Probably no ● Probably yes ○ Yes ○ Varies ○ Don't know |  |  |
| Desirable Effects  How substantial are the desirable anticipated effects? | | |
| Judgement | Research evidence | Additional considerations |
| ○ Trivial ○ Small ○ Moderate ○ Large ○ Varies ● Don't know | **Outcomes** \| **Relative effect (95% CI)** \| **Anticipated absolute effects^*^ (95% CI)** \| \| \| **Certainty of the evidence (GRADE)** \| **What happens**; \| \| **Without Antiplatelet therapy** \| **With Antiplatelet therapy** \| **Difference** \| \|; Disease Progression № of participants: 136 (1 RCT) \| - \| The mean disease Progression without antiplatelet therapy was **0** \| - \| MD **0.7 higher** (0.01 lower to 1.41 higher) \| ⨁⨁◯◯ Low^a,b^ \| Rated down 2 levels for wide confidence intervals that include the line of no effect and low event rate. Only one study identified therefore publication bias could not be formally assessed. |  |
| Undesirable Effects  How substantial are the undesirable anticipated effects? | | |
| Judgement | Research evidence | Additional considerations |
| ○ Large ○ Moderate ○ Small ○ Trivial ○ Varies ● Don't know | **Outcomes** \| **№ of participants (studies) Follow-up** \| **Certainty of the evidence (GRADE)** \| **Relative effect (95% CI)** \| **Anticipated absolute effects^*^ (95% CI)** \|; \| \| \| \| **Risk with placebo** \| **Risk difference with Antiplatelet therapy**; Disease Progression \| 136 (1 RCT) \| ⨁⨁◯◯ Low^a,b^ \| - \| The mean disease Progression was **0** \| MD **0.7 higher** (0.01 lower to 1.41 higher) Rated down 2 levels for wide confidence intervals that include the line of no effect and low event rate. Only one study identified therefore publication bias could not be formally assessed. |  |
| Certainty of evidence  What is the overall certainty of the evidence of effects? | | |
| Judgement | Research evidence | Additional considerations |
| ● Very low ○ Low ○ Moderate ○ High ○ No included studies | Only one RCT identified. Very low certainty of evidence. |  |
| Values  Is there important uncertainty about or variability in how much people value the main outcomes? | | |
| Judgement | Research evidence | Additional considerations |
| ○ Important uncertainty or variability ○ Possibly important uncertainty or variability ● Probably no important uncertainty or variability ○ No important uncertainty or variability |  |  |
| Balance of effects  Does the balance between desirable and undesirable effects favor the intervention or the comparison? | | |
| Judgement | Research evidence | Additional considerations |
| ○ Favors the comparison ○ Probably favors the comparison ○ Does not favor either the intervention or the comparison ○ Probably favors the intervention ○ Favors the intervention ○ Varies ● Don't know | Only one RCT identified and therefore unclear if there is a benefit in this specific population is unclear. | Possible stroke increase (reference RCT). |
| Resources required  How large are the resource requirements (costs)? | | |
| Judgement | Research evidence | Additional considerations |
| ○ Large costs ● Moderate costs ○ Negligible costs and savings ○ Moderate savings ○ Large savings ○ Varies ○ Don't know | Patient perspective- concerns regarding costs of medication and balance of benefits. |  |
| Certainty of evidence of required resources  What is the certainty of the evidence of resource requirements (costs)? | | |
| Judgement | Research evidence | Additional considerations |
| ○ Very low ○ Low ○ Moderate ○ High ● No included studies |  |  |
| Cost effectiveness  Does the cost-effectiveness of the intervention favor the intervention or the comparison? | | |
| Judgement | Research evidence | Additional considerations |
| ○ Favors the comparison ○ Probably favors the comparison ○ Does not favor either the intervention or the comparison ○ Probably favors the intervention ○ Favors the intervention ○ Varies ● No included studies |  |  |
| Equity  What would be the impact on health equity? | | |
| Judgement | Research evidence | Additional considerations |
| ○ Reduced ○ Probably reduced ● Probably no impact ○ Probably increased ○ Increased ○ Varies ○ Don't know |  |  |
| Acceptability  Is the intervention acceptable to key stakeholders? | | |
| Judgement | Research evidence | Additional considerations |
| ○ No ○ Probably no ● Probably yes ○ Yes ○ Varies ○ Don't know |  |  |
| Feasibility  Is the intervention feasible to implement? | | |
| Judgement | Research evidence | Additional considerations |
| ○ No ○ Probably no ● Probably yes ○ Yes ○ Varies ○ Don't know |  |  |

Summary of judgements

|  | **Judgement** | | | | | | |
| --- | --- | --- | --- | --- | --- | --- | --- |
| **Problem** | No | Probably no | **Probably yes** | Yes |  | Varies | Don't know |
| **Desirable Effects** | Trivial | Small | Moderate | Large |  | Varies | **Don't know** |
| **Undesirable Effects** | Large | Moderate | Small | Trivial |  | Varies | **Don't know** |
| **Certainty of evidence** | **Very low** | Low | Moderate | High |  |  | No included studies |
| **Values** | Important uncertainty or variability | Possibly important uncertainty or variability | **Probably no important uncertainty or variability** | No important uncertainty or variability |  |  |  |
| **Balance of effects** | Favors the comparison | Probably favors the comparison | Does not favor either the intervention or the comparison | Probably favors the intervention | Favors the intervention | Varies | **Don't know** |
| **Resources required** | Large costs | **Moderate costs** | Negligible costs and savings | Moderate savings | Large savings | Varies | Don't know |
| **Certainty of evidence of required resources** | Very low | Low | Moderate | High |  |  | **No included studies** |
| **Cost effectiveness** | Favors the comparison | Probably favors the comparison | Does not favor either the intervention or the comparison | Probably favors the intervention | Favors the intervention | Varies | **No included studies** |
| **Equity** | Reduced | Probably reduced | **Probably no impact** | Probably increased | Increased | Varies | Don't know |
| **Acceptability** | No | Probably no | **Probably yes** | Yes |  | Varies | Don't know |
| **Feasibility** | No | Probably no | **Probably yes** | Yes |  | Varies | Don't know |

Type of recommendation

| Strong recommendation against the intervention | Conditional recommendation against the intervention | **Conditional recommendation for either the intervention or the comparison** | Conditional recommendation for the intervention | Strong recommendation for the intervention |
| --- | --- | --- | --- | --- |
| ○ | ○ | **●** | ○ | ○ |

Conclusions

Recommendation

We cannot make a recommendation for the use of antiplatelet medication in this patient population. There is no evidence for the use of antiplatelet therapy in this patient population.

Remarks:

Use of antiplatelet therapy should be used based on other cardiovascular risk factors. Recommendation in prior guidelines for the use of antiplatelet therapy in patients with known anthrosclerotic disease.

Research priorities

Randomized control trials in the use of antiplatelet therapy in this patient populations.

### eTable 12 – Evidence-to-decision framework for the PICO “*Should Anti-diabetic therapy vs. placebo be used for patients with aortopathy?*”

The table below reports the GUIDE evidence-to-decision process to discuss the PICO related to the role of glycaemic control in patients with aortopathy.

| Question | |
| --- | --- |
| **Should Anti-diabetic therapy vs. placebo be used for patients with aortopathy?** | |
| **Population:** | Aortic syndromes |
| **Intervention:** | Glycemic Control |
| **Comparison:** | placebo |
| **Main outcomes:** | Mortality; AAA Growth >5mm; Incidence of Dissection-diabetes versus no diabetes; Incidence of aortic diseases -diabetes versus no diabetes; |
| **Setting:** |  |
| **Perspective:** |  |
| **Background:** |  |
| **Conflict of interests:** |  |

Assessment

| Problem  Is the problem a priority? | | |
| --- | --- | --- |
| Judgement | Research evidence | Additional considerations |
| ○ No ○ Probably no ○ Probably yes ○ Yes ○ Varies ○ Don't know |  |  |
| Desirable Effects  How substantial are the desirable anticipated effects? | | |
| Judgement | Research evidence | Additional considerations |
| ○ Trivial ○ Small ○ Moderate ○ Large ○ Varies ○ Don't know | **Outcomes** \| **№ of participants (studies) Follow-up** \| **Certainty of the evidence (GRADE)** \| **Relative effect (95% CI)** \| **Anticipated absolute effects^*^ (95% CI)** \|; \| \| \| \| **Risk with placebo** \| **Risk difference with Glycemic Control**; Mortality \| 0 (1 RCT) \| ⨁◯◯◯ Very low^a,b^ \| **HR 7.39** (1.55 to 35.23) \| Study population \|; \| \| \| \| 0 per 1,000 \| **-- per 1,000** (-- to --); AAA Growth >5mm \| 0 (1 RCT) \| ⨁◯◯◯ Very low^b,c^ \| **HR 0.43** (0.24 to 0.77) \| Study population \|; \| \| \| \| 0 per 1,000 \| **-- per 1,000** (-- to --); Incidence of Dissection-diabetes versus no diabetes \| 0 (14 observational studies) \| ⨁◯◯◯ Very low^c,d^ \| **OR 0.61** (0.42 to 0.88) \| Study population \|; \| \| \| \| 0 per 1,000 \| **0 fewer per 1,000** (0 fewer to 0 fewer); Incidence of aortic diseases -diabetes versus no diabetes \| 843536 (10 observational studies) \| ⨁◯◯◯ Very low^c^ \| **RR 0.81** (0.68 to 0.97) \| Study population \|; \| \| \| \| 193 per 1,000 \| **37 fewer per 1,000** (62 fewer to 6 fewer) wide confidence intervals and low number of effects due to low number of studies publication bias could not be assessed. examined the difference in aortopathy with diabetes versus no diabetes. inconsistency amongst study results, I^2^82% |  |
| Undesirable Effects  How substantial are the undesirable anticipated effects? | | |
| Judgement | Research evidence | Additional considerations |
| ○ Large ○ Moderate ○ Small ○ Trivial ○ Varies ○ Don't know | **Outcomes** \| **Anticipated absolute effects^*^ (95% CI)** \| \| **Relative effect (95% CI)** \| **№ of participants (studies)** \| **Certainty of the evidence (GRADE)** \| **Comments**; \| **Risk with placebo** \| **Risk with Glycemic Control** \| \| \| \|; Mortality \| Study population \| \| **HR 7.39** (1.55 to 35.23) \| (1 RCT) \| ⨁◯◯◯ Very low^a,b^ \|; \| 0 per 1,000 \| **NaN per 1,000** (-- to --) \| \| \| \|; AAA Growth >5mm \| Study population \| \| **HR 0.43** (0.24 to 0.77) \| (1 RCT) \| ⨁◯◯◯ Very low^b,c^ \|; \| 0 per 1,000 \| **NaN per 1,000** (-- to --) \| \| \| \|; Incidence of Dissection-diabetes versus no diabetes \| Study population \| \| **OR 0.61** (0.42 to 0.88) \| (14 observational studies) \| ⨁◯◯◯ Very low^c,d^ \|; \| 0 per 1,000 \| **0 per 1,000** (0 to 0) \| \| \| \|; Incidence of aortic diseases -diabetes versus no diabetes \| Study population \| \| **RR 0.81** (0.68 to 0.97) \| 843536 (10 observational studies) \| ⨁◯◯◯ Very low^c^ \|; \| 193 per 1,000 \| **156 per 1,000** (131 to 187) \| \| \| \| Wide confidence intervals and low number of effects due to low number of studies publication bias could not be assessed. examined the difference in aortopathy with diabetes versus no diabetes. inconsistency amongst study results, I282% |  |
| Certainty of evidence  What is the overall certainty of the evidence of effects? | | |
| Judgement | Research evidence | Additional considerations |
| ○ Very low ○ Low ○ Moderate ○ High ○ No included studies |  |  |
| Values  Is there important uncertainty about or variability in how much people value the main outcomes? | | |
| Judgement | Research evidence | Additional considerations |
| ○ Important uncertainty or variability ○ Possibly important uncertainty or variability ○ Probably no important uncertainty or variability ○ No important uncertainty or variability |  |  |
| Balance of effects  Does the balance between desirable and undesirable effects favor the intervention or the comparison? | | |
| Judgement | Research evidence | Additional considerations |
| ○ Favors the comparison ○ Probably favors the comparison ○ Does not favor either the intervention or the comparison ○ Probably favors the intervention ○ Favors the intervention ○ Varies ○ Don't know |  |  |
| Resources required  How large are the resource requirements (costs)? | | |
| Judgement | Research evidence | Additional considerations |
| ○ Large costs ○ Moderate costs ○ Negligible costs and savings ○ Moderate savings ○ Large savings ○ Varies ○ Don't know |  |  |
| Certainty of evidence of required resources  What is the certainty of the evidence of resource requirements (costs)? | | |
| Judgement | Research evidence | Additional considerations |
| ○ Very low ○ Low ○ Moderate ○ High ○ No included studies |  |  |
| Cost effectiveness  Does the cost-effectiveness of the intervention favor the intervention or the comparison? | | |
| Judgement | Research evidence | Additional considerations |
| ○ Favors the comparison ○ Probably favors the comparison ○ Does not favor either the intervention or the comparison ○ Probably favors the intervention ○ Favors the intervention ○ Varies ○ No included studies |  |  |
| Equity  What would be the impact on health equity? | | |
| Judgement | Research evidence | Additional considerations |
| ○ Reduced ○ Probably reduced ○ Probably no impact ○ Probably increased ○ Increased ○ Varies ○ Don't know |  |  |
| Acceptability  Is the intervention acceptable to key stakeholders? | | |
| Judgement | Research evidence | Additional considerations |
| ○ No ○ Probably no ○ Probably yes ○ Yes ○ Varies ○ Don't know |  |  |
| Feasibility  Is the intervention feasible to implement? | | |
| Judgement | Research evidence | Additional considerations |
| ○ No ○ Probably no ○ Probably yes ○ Yes ○ Varies ○ Don't know |  |  |

Summary of judgements

|  | **Judgement** | | | | | | |
| --- | --- | --- | --- | --- | --- | --- | --- |
| **Problem** | No | Probably no | Probably yes | Yes |  | Varies | Don't know |
| **Desirable Effects** | Trivial | Small | Moderate | Large |  | Varies | Don't know |
| **Undesirable Effects** | Large | Moderate | Small | Trivial |  | Varies | Don't know |
| **Certainty of evidence** | Very low | Low | Moderate | High |  |  | No included studies |
| **Values** | Important uncertainty or variability | Possibly important uncertainty or variability | Probably no important uncertainty or variability | No important uncertainty or variability |  |  |  |
| **Balance of effects** | Favors the comparison | Probably favors the comparison | Does not favor either the intervention or the comparison | Probably favors the intervention | Favors the intervention | Varies | Don't know |
| **Resources required** | Large costs | Moderate costs | Negligible costs and savings | Moderate savings | Large savings | Varies | Don't know |
| **Certainty of evidence of required resources** | Very low | Low | Moderate | High |  |  | No included studies |
| **Cost effectiveness** | Favors the comparison | Probably favors the comparison | Does not favor either the intervention or the comparison | Probably favors the intervention | Favors the intervention | Varies | No included studies |
| **Equity** | Reduced | Probably reduced | Probably no impact | Probably increased | Increased | Varies | Don't know |
| **Acceptability** | No | Probably no | Probably yes | Yes |  | Varies | Don't know |
| **Feasibility** | No | Probably no | Probably yes | Yes |  | Varies | Don't know |

Type of recommendation

| Strong recommendation against the intervention | Conditional recommendation against the intervention | Conditional recommendation for either the intervention or the comparison | Conditional recommendation for the intervention | Strong recommendation for the intervention |
| --- | --- | --- | --- | --- |
| ○ | ○ | ○ | ○ | ○ |

Conclusions

Recommendation

We suggest screening for diabetes and having glycemic control in those with aortic diseases to prevent complications as per general cardiovascular risk prevention (Conditional recommendation, very low certainty of evidence).

### eTable 13 – Evidence-to-decision framework for the PICO “*Should lipid control vs. placebo be used for patients with aortopathy?*”

The table below reports the GUIDE evidence-to-decision process to discuss the PICO related to the role of lipid control in patients with aortopathy.

| Question | |
| --- | --- |
| **Should Lipid Control vs. placebo be used for patients with aortopathy?** | |
| **Population:** | Aortic syndromes |
| **Intervention:** | Lipid Control |
| **Comparison:** | placebo |
| **Main outcomes:** | Mortality; Rate of change (progression); |
| **Setting:** |  |
| **Perspective:** |  |
| **Background:** |  |
| **Conflict of interests:** |  |

Assessment

| Problem  Is the problem a priority? | | |
| --- | --- | --- |
| Judgement | Research evidence | Additional considerations |
| ○ No ○ Probably no ● Probably yes ○ Yes ○ Varies ○ Don't know | Current standard of care is screening for and pharmocological treatment in this population.    Cardiovascular risk factors:  Few studies have examined lipid control in this population. |  |
| Desirable Effects  How substantial are the desirable anticipated effects? | | |
| Judgement | Research evidence | Additional considerations |
| ○ Trivial ● Small ○ Moderate ○ Large ○ Varies ○ Don't know | **Outcomes** \| **Anticipated absolute effects^*^ (95% CI)** \| \| **Relative effect (95% CI)** \| **№ of participants (studies)** \| **Certainty of the evidence (GRADE)** \| **Comments**; \| **Risk with placebo** \| **Risk with Lipid Control** \| \| \| \|; Mortality \| Study population \| \| **RR 0.20** (0.01 to 3.89) \| 36 (1 RCT) \| ⨁⨁◯◯ Low^a,b^ \|; \| 111 per 1,000 \| **22 per 1,000** (1 to 432) \| \| \| \|; Rate of change (progression) \| The mean rate of change (progression) was **0** \| MD **8.3 lower** (10.19 lower to 6.41 lower) \| - \| 36 (1 RCT) \| ⨁⨁◯◯ Low^b,c,d^ \| Rated down two levels for large confidence intervals that include the line of of no effect, and few number of events. Only one study identified therefore publication bias could not be assessed. Rated down for low number of events Wide confidence intervals | From a patient perspective- progression of the disease does matter to patients, although the effect is small. |
| Undesirable Effects  How substantial are the undesirable anticipated effects? | | |
| Judgement | Research evidence | Additional considerations |
| ○ Large ○ Moderate ● Small ○ Trivial ○ Varies ○ Don't know | **Outcomes** \| **Relative effect (95% CI)** \| **Anticipated absolute effects^*^ (95% CI)** \| \| \| **Certainty of the evidence (GRADE)** \| **What happens**; \| \| **Without Lipid Control** \| **With Lipid Control** \| **Difference** \| \|; Mortality № of participants: 36 (1 RCT) \| **RR 0.20** (0.01 to 3.89) \| Study population \| \| \| ⨁⨁◯◯ Low^a,b^ \|; \| \| 11.1% \| **2.2%** (0.1 to 43.2) \| **8.9% fewer** (11 fewer to 32.1 more) \| \|; Rate of change (progression) № of participants: 36 (1 RCT) \| - \| The mean rate of change (progression) without lipid Control was **0** \| - \| MD **8.3 lower** (10.19 lower to 6.41 lower) \| ⨁⨁◯◯ Low^b,c,d^ \| Rated down two levels for large confidence intervals that include the line of of no effect, and few number of events. Only one study identified therefore publication bias could not be assessed. Rated down for low number of events wide confidence intervals | Patient perception- side effects can be debilitating to patients. Who would benefit versus who will not.  In the trial one with CK increase. /Erythema noted in patients. |
| Certainty of evidence  What is the overall certainty of the evidence of effects? | | |
| Judgement | Research evidence | Additional considerations |
| ● Very low ○ Low ○ Moderate ○ High ○ No included studies | One small RCT. Evidence rated down for wide confidence intervals and low number of events. Other considerations include heterogeneity of patient population- 36 patients with type B dissection. |  |
| Values  Is there important uncertainty about or variability in how much people value the main outcomes? | | |
| Judgement | Research evidence | Additional considerations |
| ○ Important uncertainty or variability ○ Possibly important uncertainty or variability ○ Probably no important uncertainty or variability ● No important uncertainty or variability | Patient partners- we have identified the right outcomes that are highly valued by patients. |  |
| Balance of effects  Does the balance between desirable and undesirable effects favor the intervention or the comparison? | | |
| Judgement | Research evidence | Additional considerations |
| ○ Favors the comparison ○ Probably favors the comparison ● Does not favor either the intervention or the comparison ○ Probably favors the intervention ○ Favors the intervention ○ Varies ○ Don't know | Given the level of evidence, and the indirectness of the patient population, might not be applicable to the target population.  Indirect evidence supports management of cardiovascular risk factors. |  |
| Resources required  How large are the resource requirements (costs)? | | |
| Judgement | Research evidence | Additional considerations |
| ○ Large costs ○ Moderate costs ○ Negligible costs and savings ○ Moderate savings ○ Large savings ○ Varies ● Don't know | Patient perspective- if on lipid lowering drugs, and need to be on lifelong, this could be moderate costs.  Difference in cost can be significant different across countries and regions. |  |
| Certainty of evidence of required resources  What is the certainty of the evidence of resource requirements (costs)? | | |
| Judgement | Research evidence | Additional considerations |
| ○ Very low ○ Low ○ Moderate ○ High ● No included studies |  |  |
| Cost effectiveness  Does the cost-effectiveness of the intervention favor the intervention or the comparison? | | |
| Judgement | Research evidence | Additional considerations |
| ○ Favors the comparison ○ Probably favors the comparison ○ Does not favor either the intervention or the comparison ○ Probably favors the intervention ○ Favors the intervention ○ Varies ● No included studies |  |  |
| Equity  What would be the impact on health equity? | | |
| Judgement | Research evidence | Additional considerations |
| ○ Reduced ○ Probably reduced ○ Probably no impact ○ Probably increased ○ Increased ○ Varies ● Don't know |  |  |
| Acceptability  Is the intervention acceptable to key stakeholders? | | |
| Judgement | Research evidence | Additional considerations |
| ○ No ○ Probably no ● Probably yes ○ Yes ○ Varies ○ Don't know | Lack of evidence in effect in aortic dissection. But is used in practice already.  They are easy to use and not a large pill burden. | Widely used in practice by clinicians. Acceptability amongst patients depends on tolerance of the drug side effects. However, unclear benefit in this patient population after reviewing data. |
| Feasibility  Is the intervention feasible to implement? | | |
| Judgement | Research evidence | Additional considerations |
| ○ No ○ Probably no ● Probably yes ○ Yes ○ Varies ○ Don't know |  |  |

Summary of judgements

|  | **Judgement** | | | | | | |
| --- | --- | --- | --- | --- | --- | --- | --- |
| **Problem** | No | Probably no | **Probably yes** | Yes |  | Varies | Don't know |
| **Desirable Effects** | Trivial | **Small** | Moderate | Large |  | Varies | Don't know |
| **Undesirable Effects** | Large | Moderate | **Small** | Trivial |  | Varies | Don't know |
| **Certainty of evidence** | **Very low** | Low | Moderate | High |  |  | No included studies |
| **Values** | Important uncertainty or variability | Possibly important uncertainty or variability | Probably no important uncertainty or variability | **No important uncertainty or variability** |  |  |  |
| **Balance of effects** | Favors the comparison | Probably favors the comparison | **Does not favor either the intervention or the comparison** | Probably favors the intervention | Favors the intervention | Varies | Don't know |
| **Resources required** | Large costs | Moderate costs | Negligible costs and savings | Moderate savings | Large savings | Varies | **Don't know** |
| **Certainty of evidence of required resources** | Very low | Low | Moderate | High |  |  | **No included studies** |
| **Cost effectiveness** | Favors the comparison | Probably favors the comparison | Does not favor either the intervention or the comparison | Probably favors the intervention | Favors the intervention | Varies | **No included studies** |
| **Equity** | Reduced | Probably reduced | Probably no impact | Probably increased | Increased | Varies | **Don't know** |
| **Acceptability** | No | Probably no | **Probably yes** | Yes |  | Varies | Don't know |
| **Feasibility** | No | Probably no | **Probably yes** | Yes |  | Varies | Don't know |

Type of recommendation

| Strong recommendation against the intervention | Conditional recommendation against the intervention | **Conditional recommendation for either the intervention or the comparison** | Conditional recommendation for the intervention | Strong recommendation for the intervention |
| --- | --- | --- | --- | --- |
| ○ | ○ | **●** | ○ | ○ |

Conclusions

Recommendation

We cannot make a recommendation at this time for the use of lipid control in this population. (Very low certainty of evidence).

Remarks:

Lipid control could be considered in this patient population based on individual factors, but data is lacking on the effect of lipid control in this population on primary aortic dissection prevention.

Research priorities

Further research should focus on the benefit of lipid control in patients with or at risk for aortic dissection.

Biological plausibility studies are required

### Appendices

#### Appendix 1: Reporting standards

| **Standards for Reporting Qualitative Research (SRQR)*** |  |
| --- | --- |
| <http://www.equator-network.org/reporting-guidelines/srqr/> |  |
|  | **Page/line no(s).** |
| **Title and abstract** | |
| **Title** - Concise description of the nature and topic of the study Identifying the study as qualitative or indicating the approach (e.g., ethnography, grounded theory) or data collection methods (e.g., interview, focus group) is recommended | 1 (*mixed methods*) |
| **Abstract** - Summary of key elements of the study using the abstract format of the intended publication; typically includes background, purpose, methods, results, and conclusions | 2 (structured abstract) |
| **Introduction** | |
| **Problem formulation** - Description and significance of the problem/phenomenon studied; review of relevant theory and empirical work; problem statement | 3 (Introduction) |
| **Purpose or research questio**n - Purpose of the study and specific objectives or questions | 4 (Methods - *Qualitative Research and Stakeholder Surveys)* |
| **Methods** | |
| **Qualitative approach and research paradigm** - Qualitative approach (e.g., ethnography, grounded theory, case study, phenomenology, narrative research) and guiding theory if appropriate; identifying the research paradigm (e.g., postpositivist, constructivist/ interpretivist) is also recommended; rationale** | Supplementary Material (Technology Adoption Model, Theory of Planned Behaviour, and Health Literacy) |
| **Researcher characteristics and reflexivity** - Researchers’ characteristics that may influence the research, including personal attributes, qualifications/experience, relationship with participants, assumptions, and/or presuppositions; potential or actual interaction between researchers’ characteristics and the research questions, approach, methods, results, and/or transferability | 4 (Methods - *Qualitative Research and Stakeholder Surveys)* |
| **Context** - Setting/site and salient contextual factors; rationale** | 4 (Methods - *Qualitative Research and Stakeholder Surveys)* |
| **Sampling strategy** - How and why research participants, documents, or events were selected; criteria for deciding when no further sampling was necessary (e.g., sampling saturation); rationale** | 4 (Methods - *Qualitative Research and Stakeholder Surveys)* |
| **Ethical issues pertaining to human subjects** - Documentation of approval by an appropriate ethics review board and participant consent, or explanation for lack thereof; other confidentiality and data security issues | 3 (E*thics)* |
| **Data collection methods** - Types of data collected; details of data collection procedures including (as appropriate) start and stop dates of data collection and analysis, iterative process, triangulation of sources/methods, and modification of procedures in response to evolving study findings; rationale** | 4 (Methods - *Qualitative Research and Stakeholder Surveys)* |
| **Data collection instruments and technologies** - Description of instruments (e.g., interview guides, questionnaires) and devices (e.g., audio recorders) used for data collection; if/how the instrument(s) changed over the course of the study | 3 (Ethics*)* |
| **Units of study** - Number and relevant characteristics of participants, documents, or events included in the study; level of participation (could be reported in results) | 4 (Methods - *Qualitative Research and Stakeholder Surveys)* |
| **Data processing** - Methods for processing data prior to and during analysis, including transcription, data entry, data management and security, verification of data integrity, data coding, and anonymization/de-identification of excerpts | 4 (Methods - *Qualitative Research and Stakeholder Surveys)* |
| **Data analysis** - Process by which inferences, themes, etc., were identified and developed, including the researchers involved in data analysis; usually references a specific paradigm or approach; rationale** | 4 (Methods - *Qualitative Research and Stakeholder Surveys)* |
| **Techniques to enhance trustworthiness** - Techniques to enhance trustworthiness and credibility of data analysis (e.g., member checking, audit trail, triangulation); rationale** | 4 (Methods - *Qualitative Research and Stakeholder Surveys)* |
| **Results/findings** | |
| **Synthesis and interpretation** - Main findings (e.g., interpretations, inferences, and themes); might include development of a theory or model, or integration with prior research or theory | 6 (Results - *Stakeholder engagement exercise and Focus Group)* |
| **Links to empirical data** - Evidence (e.g., quotes, field notes, text excerpts, photographs) to substantiate analytic findings | Supplementary Material |
| **Discussion** | |
| **Integration with prior work, implications, transferability, and contribution(s) to the field -** Short summary of main findings; explanation of how findings and conclusions connect to, support, elaborate on, or challenge conclusions of earlier scholarship; discussion of scope of application/generalizability; identification of unique contribution(s) to scholarship in a discipline or field | 7 (Discussion - *Stakeholder engagement exercise and Focus Group)* |
| **Limitations** - Trustworthiness and limitations of findings | 7 (Discussion - *Stakeholder engagement exercise and Focus Group)* |
| **Other** | |
| **Conflicts of interest** - Potential sources of influence or perceived influence on study conduct and conclusions; how these were managed | 7 (Discussion - *Stakeholder engagement exercise and Focus Group)* |
| **Funding** - Sources of funding and other support; role of funders in data collection, interpretation, and reporting | 3 (E*thics)* |
| *The authors created the SRQR by searching the literature to identify guidelines, reporting standards, and critical appraisal criteria for qualitative research; reviewing the reference lists of retrieved sources; and contacting experts to gain feedback. The SRQR aims to improve the transparency of all aspects of qualitative research by providing clear standards for reporting qualitative research. |  |
| **The rationale should briefly discuss the justification for choosing that theory, approach, method, or technique rather than other options available, the assumptions and limitations implicit in those choices, and how those choices influence study conclusions and transferability. As appropriate, the rationale for several items might be discussed together. |  |
| **Reference:** |  |
| O'Brien BC, Harris IB, Beckman TJ, Reed DA, Cook DA. **Standards for reporting qualitative research: a synthesis of recommendations.** *Academic Medicine*, Vol. 89, No. 9 / Sept 2014  DOI: 10.1097/ACM.0000000000000388 |  |

**STROBE Statement—Checklist of items that should be included in reports of *cohort studies***

|  | Item No | Recommendation | Page No |
| --- | --- | --- | --- |
| **Title and abstract** | 1 | (*a*) Indicate the study’s design with a commonly used term in the title or the abstract | Page 1 |
|  |  | (*b*) Provide in the abstract an informative and balanced summary of what was done and what was found | Page 2 |
| Introduction | | | |
| Background/rationale | 2 | Explain the scientific background and rationale for the investigation being reported | Supplementary Material – Study Protocol |
| Objectives | 3 | State specific objectives, including any prespecified hypotheses | Supplementary Material – Study Protocol |
| Methods | | | |
| Study design | 4 | Present key elements of study design early in the paper | Page 3 |
| Setting | 5 | Describe the setting, locations, and relevant dates, including periods of recruitment, exposure, follow-up, and data collection | Page 3 |
| Participants | 6 | Give the eligibility criteria, and the sources and methods of selection of participants. Describe methods of follow-up | Page 3 |
| Variables | 7 | Clearly define all outcomes, exposures, predictors, potential confounders, and effect modifiers. Give diagnostic criteria, if applicable | Page 3 |
| Data sources/ measurement | 8* | For each variable of interest, give sources of data and details of methods of assessment (measurement). Describe comparability of assessment methods if there is more than one group | Page 3 |
| Bias | 9 | Describe any efforts to address potential sources of bias | Page 3 |
| Study size | 10 | Explain how the study size was arrived at | Not applicable |
| Quantitative variables | 11 | Explain how quantitative variables were handled in the analyses. If applicable, describe which groupings were chosen and why | Not applicable |
| Statistical methods | 12 | (*a*) Describe all statistical methods, including those used to control for confounding |  |
|  |  | (*b*) Describe any methods used to examine subgroups and interactions | Page 3 |
|  |  | (*c*) Explain how missing data were addressed |  |
|  |  | (*d*) If applicable, explain how loss to follow-up was addressed |  |
|  |  | (*e*) Describe any sensitivity analyses |  |
| Results | | |  |
| Participants | 13* | (a) Report numbers of individuals at each stage of study—eg numbers potentially eligible, examined for eligibility, confirmed eligible, included in the study, completing follow-up, and analysed | Page 6 |
|  |  | (b) Give reasons for non-participation at each stage |  |
|  |  | (c) Consider use of a flow diagram |  |
| Descriptive data | 14* | (a) Give characteristics of study participants (eg demographic, clinical, social) and information on exposures and potential confounders | Page 6 |
|  |  | (b) Indicate number of participants with missing data for each variable of interest |  |
|  |  | (c) Summarise follow-up time (eg, average and total amount) |  |
| Outcome data | 15* | Report numbers of outcome events or summary measures over time | Page 6 |

| Main results | 16 | (*a*) Give unadjusted estimates and, if applicable, confounder-adjusted estimates and their precision (eg, 95% confidence interval). Make clear which confounders were adjusted for and why they were included | Page 6 |
| --- | --- | --- | --- |
|  |  | (*b*) Report category boundaries when continuous variables were categorized |  |
|  |  | (*c*) If relevant, consider translating estimates of relative risk into absolute risk for a meaningful time period |  |
| Other analyses | 17 | Report other analyses done—eg analyses of subgroups and interactions, and sensitivity analyses | Page 6 |
| Discussion | | | |
| Key results | 18 | Summarise key results with reference to study objectives | Page 8 |
| Limitations | 19 | Discuss limitations of the study, taking into account sources of potential bias or imprecision. Discuss both direction and magnitude of any potential bias | Page 8 |
| Interpretation | 20 | Give a cautious overall interpretation of results considering objectives, limitations, multiplicity of analyses, results from similar studies, and other relevant evidence | Page 8 |
| Generalisability | 21 | Discuss the generalisability (external validity) of the study results | Page 8 |
| Other information | | | |
| Funding | 22 | Give the source of funding and the role of the funders for the present study and, if applicable, for the original study on which the present article is based | Page 3 |

*Give information separately for exposed and unexposed groups.

**Note:** An Explanation and Elaboration article discusses each checklist item and gives methodological background and published examples of transparent reporting. The STROBE checklist is best used in conjunction with this article (freely available on the Web sites of PLoS Medicine at http://www.plosmedicine.org/, Annals of Internal Medicine at http://www.annals.org/, and Epidemiology at http://www.epidem.com/). Information on the STROBE Initiative is available at http://www.strobe-statement.org.

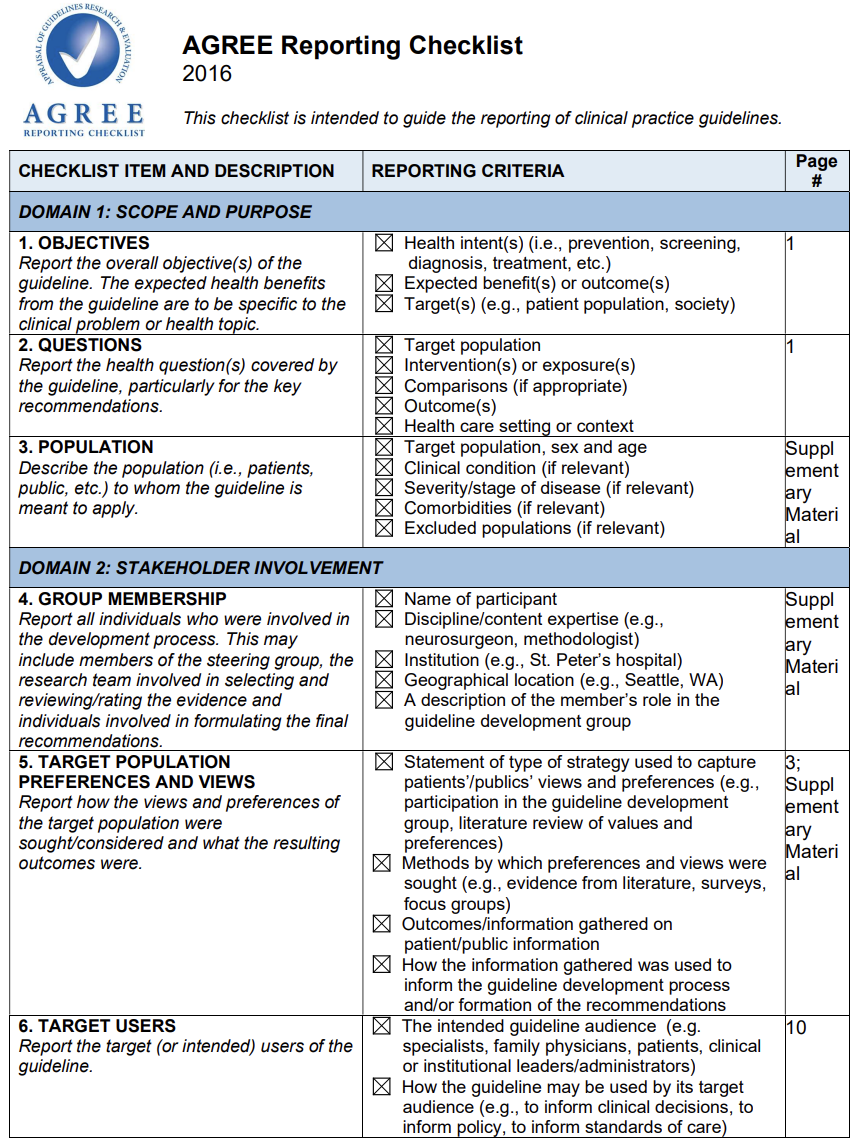

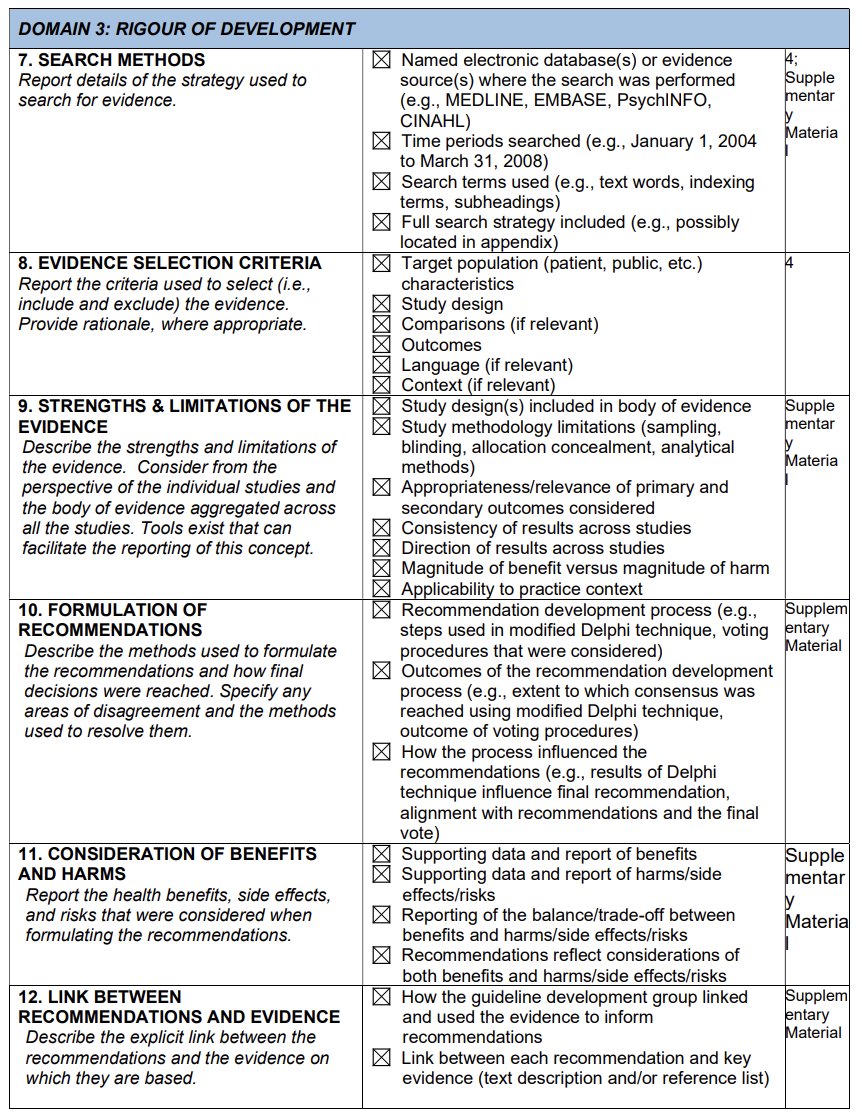

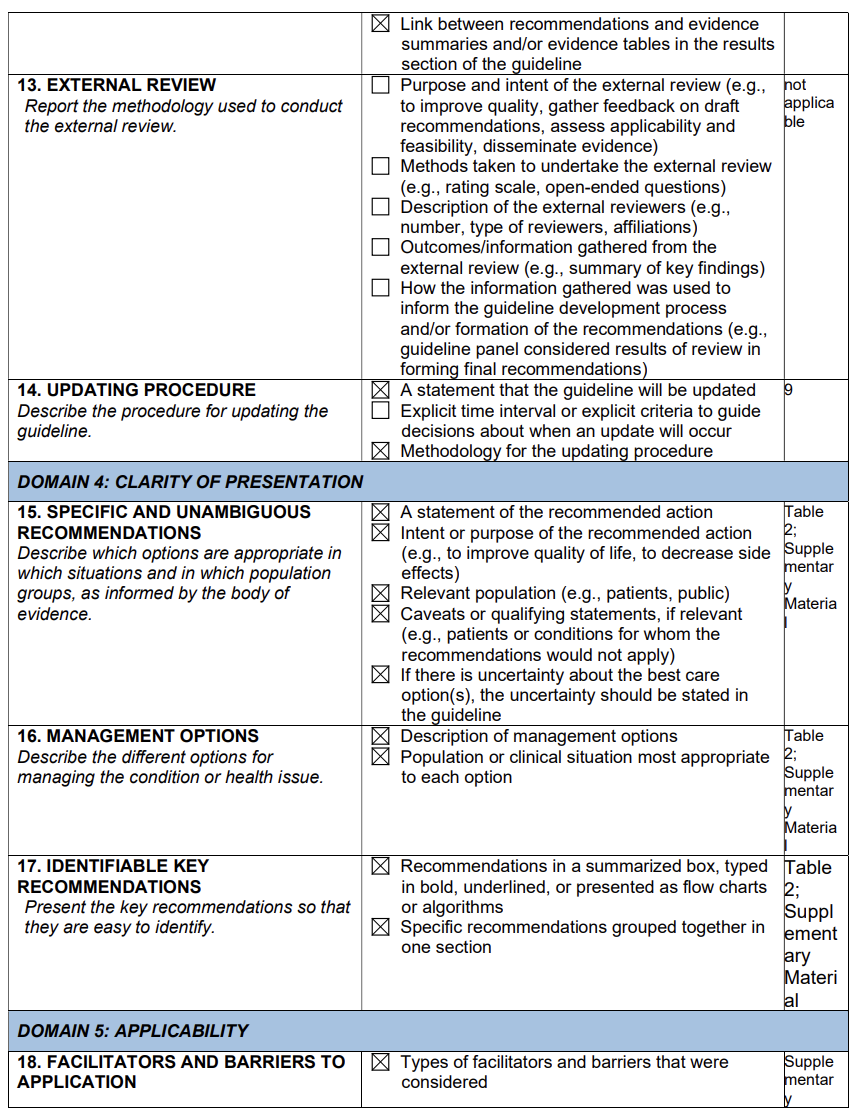

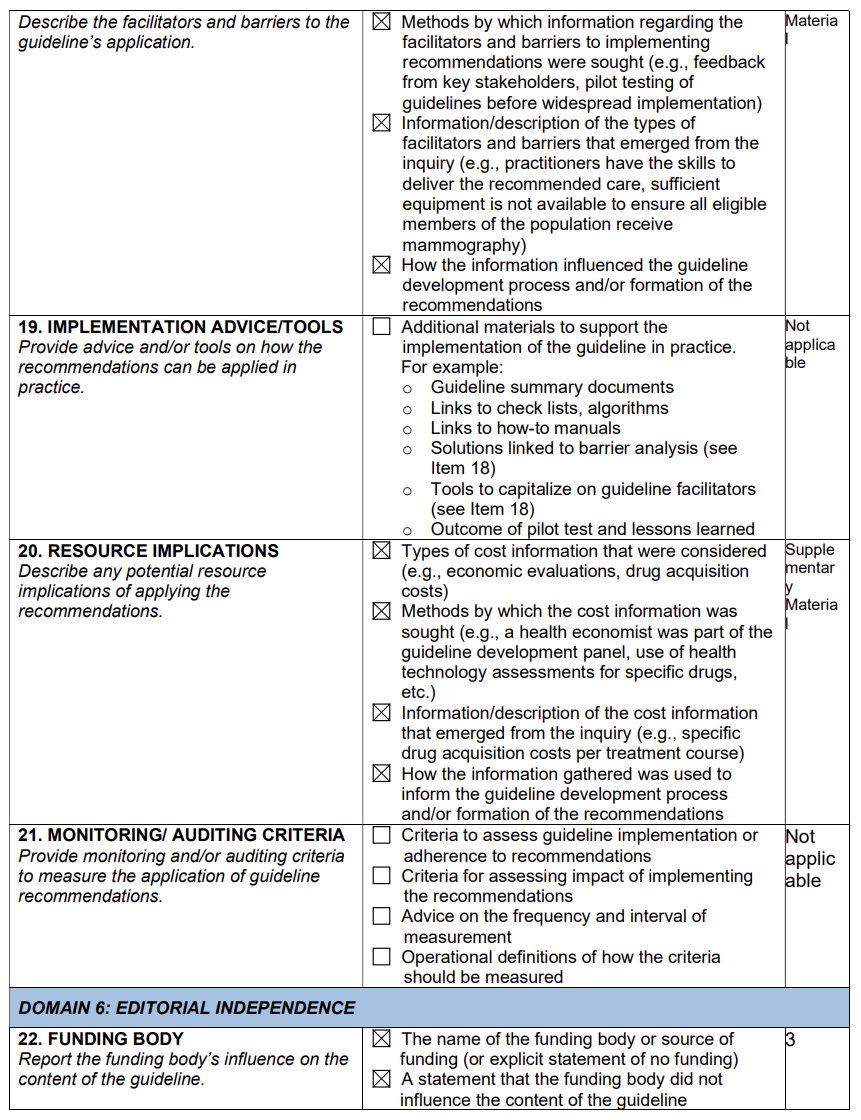

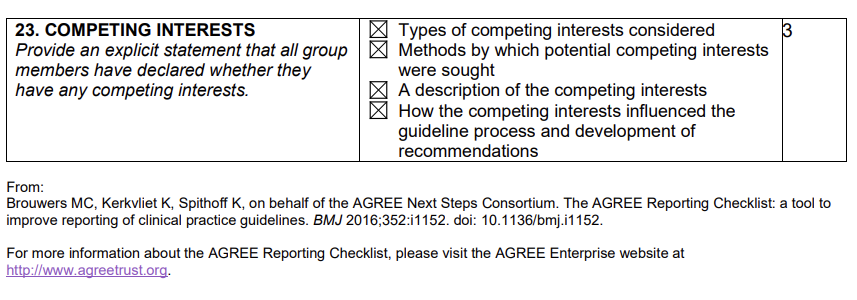

| **Section and Topic** | **Item #** | **Checklist item** | **Location where item is reported** |
| --- | --- | --- | --- |
| **TITLE** | | |  |
| Title | 1 | Identify the report as a literature review. | n/a |
| **ABSTRACT** | | |  |
| Abstract | 2 | Provide a structured summary including, as applicable: background; objectives; data sources; study eligibility criteria, participants, and interventions; study appraisal and synthesis methods; results; limitations; conclusions and implications of key findings.  See the [PRISMA 2020 for Abstracts checklist](http://www.prisma-statement.org/Extensions/Abstracts.aspx) for the complete list. | n/a |
| **INTRODUCTION** | | |  |
| Rationale | 3 | Describe the rationale for the review in the context of existing knowledge, i.e., what is already known about your topic. | *Introduction* |
| Objectives | 4 | Provide an explicit statement of the objective(s) or question(s) the review addresses with reference to participants, interventions, comparisons, outcomes, and study design (PICOS). | *Supplementary Material* |
| **METHODS** | | |  |
| Eligibility criteria | 5 | Specify the inclusion and exclusion criteria for the review and how studies were grouped for the syntheses with study characteristics (e.g., PICOS, length of follow-up) and report characteristics (e.g., years considered, language, publication status) used as criteria for eligibility, giving rationale. | *Supplementary Material – Evidence to decision framework* |
| Information sources | 6 | Specify all databases, registers, websites, organisations, reference lists and other sources searched or consulted to identify studies. Specify the date when each source was last searched or consulted. | *Methods* |
| Search strategy | 7 | Present the full search strategies for all databases, registers and websites, including any filters and limits used. | *Methods* |
| Selection process | 8 | State the process for selecting studies (i.e., screening, eligibility).  Specify the methods used to decide whether a study met the inclusion criteria of the review, including how many reviewers screened each record and each report retrieved, whether they worked independently, and if applicable, details of automation tools used in the process. | *Methods* |
| Study risk of bias assessment | 11 | Specify the methods used to assess risk of bias in the included studies, including details of the tool(s) used, how many reviewers assessed each study and whether they worked independently, and if applicable, details of automation tools used in the process. | *Methods* |
| **RESULTS** | | |  |
| Study selection | 16a | Describe the results of the search and selection process, from the number of records identified in the search to the number of studies included in the review, ideally using a flow diagram. | *Results / Supplementary Material (ETD frameworks)* |
|  | 16b | Cite studies that might appear to meet the inclusion criteria, but which were excluded, and explain why they were excluded. | *Results / Supplementary Material (ETD frameworks)* |
| Study characteristics | 17 | Cite each included study and present its characteristics (e.g., study size, PICOS, follow-up period). | *Results / Supplementary Material (ETD frameworks)* |
| Risk of bias in studies | 18 | Present assessments of risk of bias for each included study. | *Results / Supplementary Material (ETD frameworks)* |
| Results of individual studies | 19 | For all outcomes, present, for each study: (a) summary statistics for each group (where appropriate) and (b) an effect estimate and its precision (e.g. confidence/credible interval), ideally using structured tables or plots. | *Results / Supplementary Material (ETD frameworks)* |
| **DISCUSSION** | | |  |
| Discussion | 23a | Provide a general interpretation of the results in the context of other evidence. | *Discussion / Supplementary Material* |
|  | 23b | Discuss any limitations of the evidence included in the review. | *Discussion / Supplementary Material* |
|  | 23c | Discuss any limitations of the review processes used. | *Discussion / Supplementary Material* |
|  | 23d | Discuss implications of the results for practice, policy, and future research. | *Discussion / Supplementary Material* |
| **OTHER INFORMATION** | | |  |
| Registration and protocol | 24a | Provide registration information for the review, including register name and registration number, or state that the review was not registered. | n/a |
|  | 24b | Indicate where the review protocol can be accessed, or state that a protocol was not prepared. | n/a |
|  | 24c | Describe and explain any amendments to information provided at registration or in the protocol. | n/a |
| Support | 25 | Describe sources of financial or non-financial support for the review, and the role of the funders or sponsors in the review. | *Methods* |
| Competing interests | 26 | Declare any competing interests of review authors. | *Methods* |
| Availability of data, code, and other materials | 27 | Report which of the following are publicly available and where they can be found: template data collection forms; data extracted from included studies; data used for all analyses; analytic code; any other materials used in the review. | *Methods* |

#### Appendix 2: List of ICD-10 and OPCS-4 codes used to define thoracic aorta diseases (TAD) and comorbidities in the HES analysis

Thoracic aortic disease (TAD) – ICD-10

I71.0 Dissection of aorta [any part]

I71.1 Thoracic aortic aneurysm, ruptured

I71.2 Thoracic aortic aneurysm, without mention of rupture

I71.5 Thoracoabdominal aortic aneurysm, ruptured

I71.6 Thoracoabdominal aortic aneurysm, without mention of rupture

TAD procedure – OPCS-4

L18.1 Emergency replacement of aneurysmal segment of ascending aorta by anastomosis of aorta to aorta

L18.2 Emergency replacement of aneurysmal segment of thoracic aorta by anastomosis of aorta to aorta NEC

L19.1 Replacement of aneurysmal segment of ascending aorta by anastomosis of aorta to aorta NEC

L19.2 Replacement of aneurysmal segment of thoracic aorta by anastomosis of aorta to aorta NEC

L20.1 Emergency bypass of segment of ascending aorta by anastomosis of aorta to aorta NEC

L20.2 Emergency bypass of segment of thoracic aorta by anastomosis of aorta to aorta NEC

L20.8 Other specified other emergency bypass of segment of aorta

L20.9 Unspecified other emergency bypass of segment of aorta

L21.1 Bypass of segment of ascending aorta by anastomosis of aorta to aorta NEC

L21.2 Bypass of segment of thoracic aorta by anastomosis of aorta to aorta NEC

L27.3 Endovascular insertion of stent graft for thoracic aortic aneurysm

L28.3 Endovascular insertion of stent for thoracic aortic aneurysm

L22.1 Revision of prosthesis of thoracic aorta

#### Appendix 3: Search Strategy for the systematic reviews

*Prevention*

MEDLINE through OVID (Run on 4th August 2021, adapted for other databases: Embase, CENTRAL)

1	(Aortic or Aorta).mp.

2	exp aorta/

3	1 or 2

4	exp Preventive Health Services/

5	(prevent* or prophyla*).mp.

6	4 or 5

7	3 and 6

8	limit 7 to (human and randomized controlled trial)

**2069** records retrieved

*Surveillance and Screening*

MEDLINE through OVID (Run on 6th August 2022, adapted for other databases: Embase, CENTRAL)

1	(Aortic or Aorta).mp.

2	exp aorta/

3	1 or 2

4	exp Secondary Care/

5	exp Diagnostic Screening Programs/

6	(follow-up or "follow up").ab.

7	surveillance.ti,ab.

8	screening.ti,ab.

9	4 or 5 or 6 or 7 or 8

10	3 and 9

11	limit 10 to (humans and randomized controlled trial and last 10 years)

**2149** records retrieved

*Guidelines and Consensus*

Searches performed on Medline (476 references) and Embase (2696 references) on the 21^st^ October 2020

1. aorta/ or thoracic aorta/ or ascending aorta surgery/ or ascending aorta/ or aorta.mp. or thoracic aorta aneurysm/

2. gene*.mp. or genetics/ or echocardiography.mp or computed tomography.mp or magnetic resonance.mp or US.mp or CT.mp or MRI.mp

3. (consensus or recommendation* or guideline* or expert).ti,ab.

4. (surveillance or management or therapy or test or indication or threshold).mp.

5. 1 and 2 and 3 and 4

**2897** records retrieved
